# Design characteristics of Sequential Multiple Assignment Randomized Trials (SMARTs) for human health: a scoping review of studies between 2009-2024

**DOI:** 10.1101/2025.06.06.25329149

**Authors:** Nikki L. B. Freeman, Sydney E. Browder, Bryce T. Rowland, Emily P. Jones, Margaret Hoch, Alice H. Kim, Christina W. Zhou, Anna R. Kahkoska, Katharine L. McGinigle, Anastasia Ivanova, Michael R. Kosorok, Kevin J. Anstrom

## Abstract

**Objective:** To characterize the reporting practices of sequential multiple assignment randomized trials (SMARTs) in human health research.

**Design:** Scoping review of protocol and primary analysis papers describing SMARTs published between January 2009 and February 2024.

**Background:** SMARTs are innovative trial designs that allow for multiple stages of randomization to treatment based on a patient’s responses to previous treatments. They are uniquely designed to develop sequential adaptive interventions (dynamic treatment regimes) to support clinical decision-making over time. Previous reviews have identified inconsistencies in how the design, implementation, and results have been reported in published studies. A comprehensive assessment of SMART reporting practices is lacking and is necessary for developing standardized SMART-specific reporting guidelines.

**Methods:** We systematically searched multiple databases for SMART-related protocol and primary analysis papers published between January 2009 and February 2024. Title, abstract, and full-text screenings were performed by pairs of reviewers, with disagreements resolved by consensus. Data extraction included study characteristics, design elements, and analytic approaches for embedded or tailored dynamic treatment regimes (DTRs). Results were synthesized qualitatively and presented descriptively.

**Results:** From 5486 screened studies, 103 (59 protocol papers, 16 primary analysis papers, 14 protocol papers with corresponding primary analysis papers) met the inclusion criteria. Most studies targeted adults (62.7% protocols, 62.5% primary analyses, 42.9% protocol + primary analyses) and were primarily conducted in the United States. Behavioral and mental health constituted the most frequent therapeutic domain. While intervention descriptions and re-randomization criteria were consistently reported, operational characteristics such as blinding (protocols: 64.4%, primary analyses: 62.5%, protocols + primary analyses: 71.4%) and randomization details (protocols: 55.9%, primary analyses: 37.5%, protocols + primary analyses: 50.0%) were inconsistently documented. Only 46.7% of primary analyses evaluated embedded DTRs, and none explored deeply tailored DTRs.

**Conclusions:** Despite the increased adoption of SMART designs, substantial reporting variability persists. Most primary analyses underutilize the capability of SMARTs to generate data for developing dynamic treatment regimes. SMART-specific standardized reporting guidelines can help accelerate the scientific and clinical impact of SMARTs.

**Strengths and limitations of this study:** *Strengths:*

- This is the most comprehensive review of SMARTs in human health to date, spanning studies published since SMARTS were introduced and over a 15-year period (2009-2024).
- Rigorous methodology, including dual independent screening and extraction, enhances the reliability of the findings.
- The granularity of our assessment of design elements provides specific targets for developing SMART reporting guidelines.

*Limitations:*

- This study only included protocol and primary analysis papers, missing SMARTs described in trial registries alone as well as analyses of SMARTs in secondary analysis papers.
- This review included only English-language publications, potentially missing relevant studies published in other languages
- This review did not assess the quality of the included SMARTs themselves, focusing instead on reporting practices.

## INTRODUCTION

Sequential multiple assignment randomized trials (SMARTs), designed to mirror the dynamic of real-world clinical decision-making where medical providers and their patients often make decisions over time and in response to evolving health status and the effects of prior treatments, are a significant advancement in clinical research(1). Formalized by Murphy and Bingham(2), "a SMART is a type of multistage, factorial randomized trial, in which some or all participants are randomized at 2 or more decision points. Whether a patient is randomized at the second or a later decision point, and the available treatment options, may depend on the patient’s response to prior treatment”(3). **Figure 1** illustrates an example of how a SMART may be designed.

**Figure 1.**
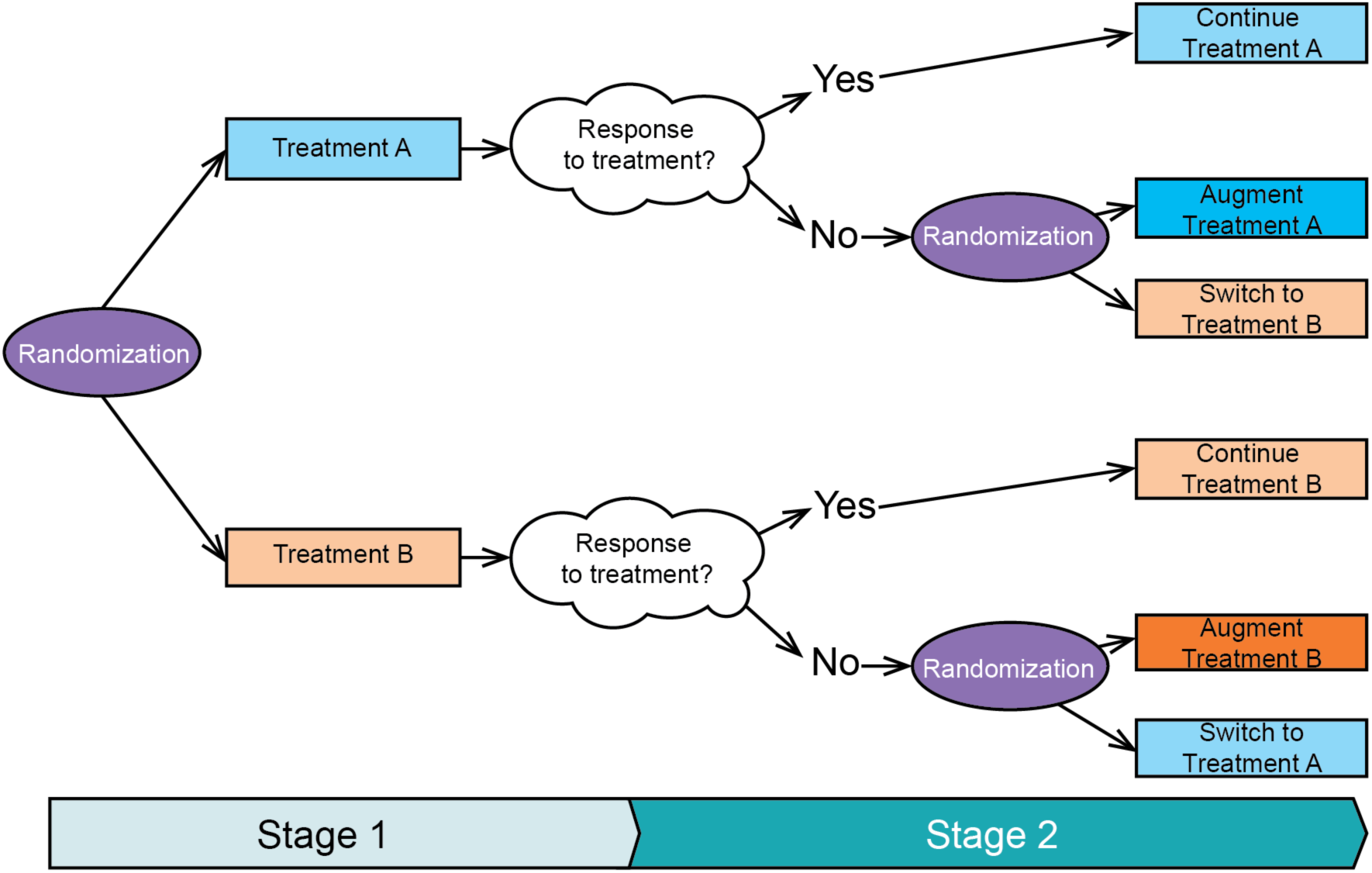
Generic SMART schematic. In this example of a particular SMART design, a trial participant may initially be randomized to one of two treatments, A or B. After 3 months, they are assessed for response to the initial treatment. If they have a positive treatment response, they will be assigned to remain on their initial treatment. Otherwise, they are re-randomized to either switch treatment or receive an augmented version of their initially assigned treatment.

SMARTs can be used to answer questions about the main effects of first-line treatments as well as the effects of second-line treatments (1). By formalizing the treatment decision process through multiple randomizations, SMARTs can also generate evidence for treatment strategies that adapt to individuals’ unique, possibly evolving characteristics across multiple treatment decisions. These treatment strategies are often evaluated in the analysis of a SMART by studying dynamic treatment regimes (DTRs), defined as sequences of decision rules that map patient characteristics to a recommended treatment decision (4,5). Embedded DTRs are predefined treatment sequences within a SMART inherent to the study design. In contrast, optimal tailored DTRs or optimal deeply tailored DTRs are DTRs that can be learned from SMART data using techniques from machine learning and reinforcement learning, but are generally not embedded in the trial itself(1). They are optimal in the sense that they attempt to generate rules that, if followed, would optimize outcomes on average in the population.

Given SMARTs’ flexibility to address multiple, often interrelated clinical questions, they can complicate the traditional hierarchy of primary, secondary, and exploratory analyses. In standard randomized controlled trials (RCTs), study objectives align with a primary hypothesis, guiding the selection of outcomes, sample size considerations, and how trial results are presented and reported. In contrast, the multipurpose nature of SMARTs makes it difficult to specify a single primary analysis, as the trial is not limited to answering one core question but rather is designed to inform a broader decision-making framework. Consequently, different researchers and trialists may prioritize different aspects of the SMART design or emphasize different analyses in published reports.

This analytic flexibility, while yielding methodological value and design efficiencies, may create substantial challenges for transparent and consistent reporting. SMARTs have been successfully conducted in multiple clinical domains, yet there is little consistency in how they are described in clinical literature. Lorenzoni et al.(6) conducted a systematic review examining SMART designs in oncology. They identified 33 records, including 15 trial reports, 4 protocols, and 14 trial registrations. They found that most studies analyzed each treatment stage separately rather than considering embedded treatment regimes. Moreover, they found that power calculations and study analyses relied primarily on statistical methods commonly used in single-stage parallel designs. The authors concluded that formal reporting guidelines for SMART designs are needed.

Bigirumurame, Uwimpuhwe, and Wason(7) conducted a broader systematic review examining the reporting quality of SMART studies across various fields. They reviewed 157 records, including 12 trial reports, 24 protocols, 91 methodological papers, and 30 review papers. Their review highlighted that key components of SMART designs were inconsistently reported, with deficiencies in reporting parameters required for sample size calculations. They also found that most trials were powered using stage-specific aims rather than leveraging the full potential of SMART designs to evaluate embedded adaptive interventions. Additionally, they noted that few studies (16.67%) focused on determining the best embedded adaptive interventions, and most trials did not report information on multiple testing adjustment.

These findings underscore a persistent gap in the reporting of the design, implementation, and results of SMARTs, hindering the interpretation, replication, and synthesis of evidence from these studies and limiting their scientific and clinical impact. The objective of this review was to systematically characterize SMART reporting practices in human health research, with a focus on design features and methodological transparency in peer-reviewed publications. We examined only protocol and primary analysis papers to provide a focused assessment of how SMARTs are initially designed and subsequently reported. We have also conducted a more granular analysis of how embedded and deeply tailored regimes are reported and analyzed, an aspect only briefly addressed in previous reviews. Finally, our review specifically aims to inform the development of reporting guidelines for SMARTs, addressing the gap identified by both previous reviews. This review’s findings can directly inform the development of SMART-specific reporting guidelines, similar to CONSORT for traditional RCTs, and enhance methodological transparency and rigor.

## METHODS

We used the PRISMA extension for scoping reviews (PRISMA-ScR)(8) to guide our process, and the full checklist is included (**Supplementary document 4**). A detailed protocol, including methods and objectives, was prospectively developed and made publicly available through the Open Science Framework(9). Our inclusion and exclusion criteria were developed to ensure a comprehensive and coherent examination of SMARTs in human health contexts. We limited inclusion to studies published from January 2009 through February 9, 2024. Although the SMART design was formally introduced in the statistical literature in 2005(10), SMARTs did not begin to appear in published human health research until around 2009. Early work between 2005-2008 focused primary on methodological development and feasibility demonstrations in simulated settings(11–13). Because our review focuses on SMARTs in practice, we chose 2009 as the anchor point for this review. Protocol/design, primary analysis, and secondary analysis papers, as well as registered trials that the authors identified as SMARTs conducted in a healthcare setting, were included in the search. Articles not in English, reviews, commentaries, letters, editorials, theses, and methodological papers were excluded. A detailed description of the full inclusion and exclusion criteria is provided in the protocol **(Supplementary document 1).**

A medical librarian (EJ) developed a comprehensive search strategy to identify potentially relevant publications in PubMed (U.S. National Library of Medicine, National Institutes of Health), Scopus (Elsevier), Embase (Elsevier), and Cochrane Central Register of Controlled Trials (CENTRAL) (Cochrane Library). The search strategy used a combination of controlled vocabulary, where applicable, and various keywords for SMART (**Supplementary document 2**). Databases were searched on February 9, 2024. Search strategies for all databases are available in the supplemental appendix. Results were imported into Endnote and deduplicated; all unique citations were then imported into Covidence for screening.

Although we included secondary analysis papers and trial registry entries in our search per our protocol, for this review, we ultimately included only protocol/design papers and primary analysis papers. This decision was motivated by prioritizing the feasibility of completing this review, identifying the publication types that could be extracted with high fidelity, and selecting publication types we believed aligned with this review’s focus. Before commencing the title and abstract screening, benchmark screenings were conducted on 100 titles and abstracts each, until inter-rater reliability, measured by Cohen’s kappa, exceeded 70% between screeners. Each title and abstract was screened independently by two reviewers (SB, MH, CZ), and conflicts were adjudicated by a third author (NF or BR) through consensus. Studies that clearly did not meet the eligibility criteria were excluded. Full-text screening for each paper was independently performed by two reviewers (SB, MH, CZ), and reasons for articles to be excluded at this stage were noted. Conflicts were adjudicated by a third author (NF) through consensus.

A standardized data extraction form developed in Microsoft Excel was piloted by the review team. Data items extracted included study characteristics (e.g., authors, publication year, country), design characteristics of the SMART designs (e.g., number of randomizations, treatment options), and operational characteristics (e.g., blinding, randomization strategies). A comprehensive list of the data items extracted is provided in **Supplementary document 3**. Data items from full texts were extracted into the extraction form by two independent extractors for each paper (SB, MH, AK, CZ, NF). Since our goal was to examine how SMARTs are reported, no additional data were sought from investigators. Two authors (NF, SB) synthesized the results from the duplicate data extraction. Patterns across the protocol and primary analysis papers were identified and described. In cases where a study had both a protocol and primary analysis paper, we analyzed them together (denoted throughout as “protocol + primary analysis”). Thus, the unit of analysis for this review is study-level (trial-level) rather than strictly paper-level.

### Patient and Public Involvement

Patients and members of the public were not involved in the design, conduct, reporting, or dissemination plans of this scoping review.

## RESULTS

We identified 10,066 citations through database searches, and 4,580 were duplicates. The remaining 5,486 citations were imported into Covidence for title/abstract screening, and 4,655 were deemed irrelevant. Full-text publications were retrieved for all remaining records (n = 831) and screened for inclusion. Of these, 532 reports were excluded, primarily for not being a SMART. The remaining 299 articles were assessed for inclusion during data extraction, with 196 excluded, mainly because they were registry entries, secondary analyses, or non-original research. In total, 103 papers were included in our analysis, including 59 protocol-only, 16 primary analysis-only, and 14 protocols with 14 corresponding primary analysis papers. The PRISMA Flow Diagram depicting the full study selection process is shown in **Figure 2**.

**Figure 2.**
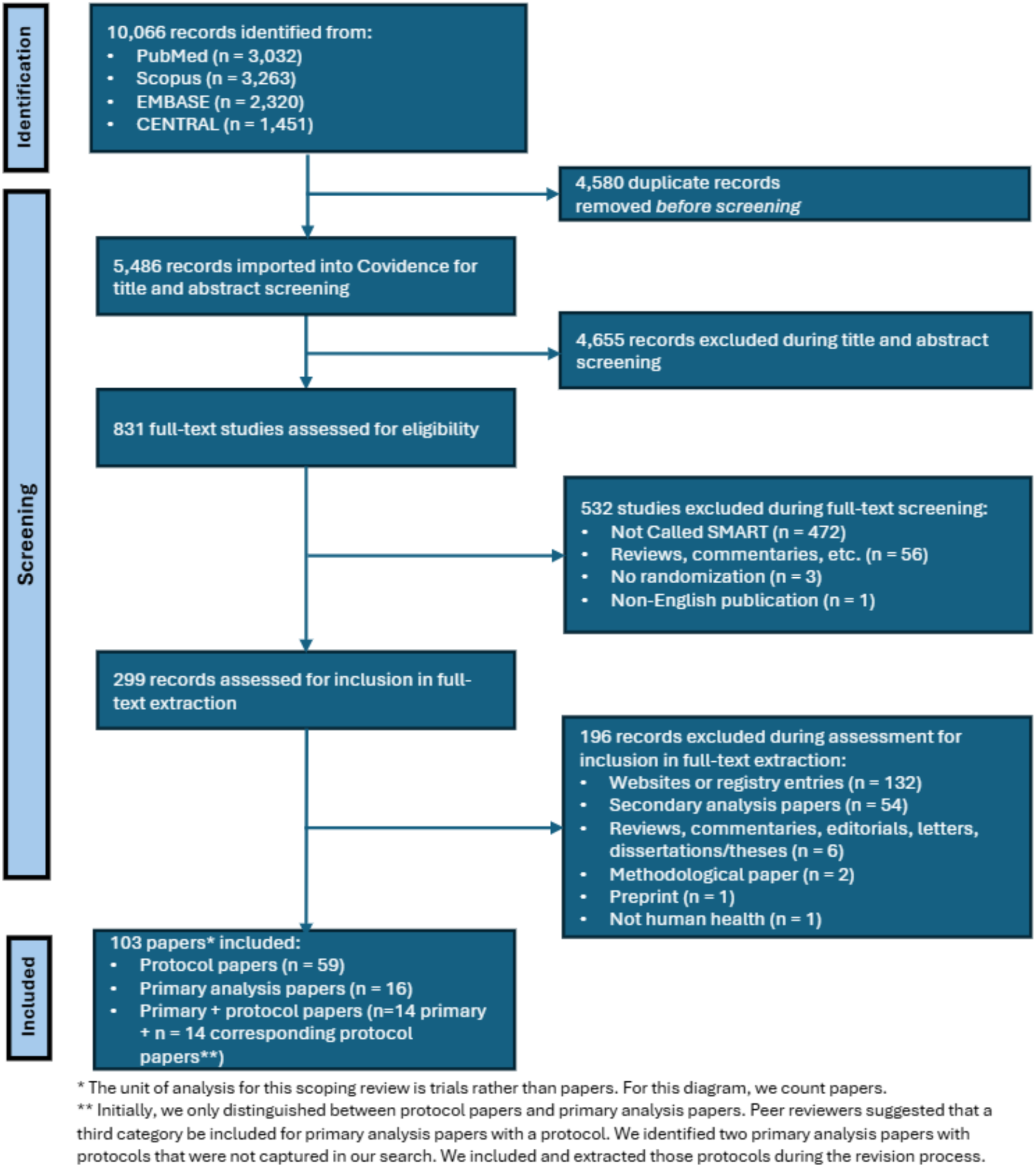
PRISMA flow diagram.

Characteristics of the included studies are summarized in **Table 1**. Seven (11.9%) protocol papers, 6 (37.5%) primary analysis papers, and 4 (28.6%) protocol + primary analysis papers were about pilot interventions. Most papers about SMARTs were published in 2019 or later [n=49 (83.1%) protocol papers; n = 11 (68.8%) primary analysis papers; 14 (100.0%) protocol + primary analysis papers]. Studies were predominantly conducted in the United States [n=44 (74.6%) protocol papers; n=13 (81.3%) primary analysis papers; 12 (85.7%) protocol + primary analysis papers]. Other countries represented included Brazil, China, Kenya, and South Africa. Two studies were conducted in multiple countries. Protocol papers were more likely to target a SMART with more than 500 participants (n=21, 35.6%) than primary analysis or protocol + primary analysis papers [n=2 (12.5%) and n=3 (21.4%), respectively]. The vast majority of SMARTs had 2 potential randomizations [n=55 protocol papers (93.2%); n=16 primary analysis papers (100.0%); n=13 protocol + primary analysis papers (92.9%)], although a few had up to 3 randomizations [n=3 protocol papers (5.1%); n=1 protocol + primary analysis paper (7.1%)] and one had 6 randomizations [n=1 protocol paper (1.7%)]. Full details are available in **Tables S1 and S2.**

**Table 1.** Study characteristics.

| **Characteristics** | **Protocol papers**<br>**(n=59)**<br>**N (%)** | **Primary analysis**<br>**papers (n=16)**<br>**N (%)** | **Protocol + primary**<br>**analysis papers (n =**<br>**14)**<br>**N (%)** |
| --- | --- | --- | --- |
| Pilot study | 7 (11.9%) | 6 (37.5%) | 4 (28.6%) |
| Publication year* |  |  |  |
| 2009 | 1 (1.7%) | 0 (0.0%) | 0 (0.0%) |
| 2014 | 1 (1.7%) | 0 (0.0%) | 0 (0.0%) |
| 2015 | 0 (0.0%) | 1 (6.3%) | 0 (0.0%) |
| 2016 | 1 (1.7%) | 2 (12.5%) | 0 (0.0%) |
| 2017 | 2 (3.4%) | 1 (6.3%) | 0 (0.0%) |
| 2018 | 5 (8.5%) | 1 (6.3%) | 0 (0.0%) |
| 2019 | 6 (10.2%) | 0 (0.0%) | 3 (21.4%) |
| 2020 | 9 (15.3%) | 2 (12.5%) | 2 (14.3%) |
| 2021 | 8 (13.6%) | 2 (12.5%) | 3 (21.4%) |
| 2022 | 6 (10.2%) | 2 (12.5%) | 2 (14.3%) |
| 2023 | 18 (30.5%) | 4 (25.0%) | 3 (21.4%) |
| 2024 | 2 (3.4%) | 1 (6.3%) | 1 (7.1%) |
| Country |  |  |  |
| Australia | 1 (1.7%) | 0 (0.0%) | 0 (0.0%) |
| Brazil | 0 (0.0%) | 1 (6.3%) | 1 (7.1%) |
| Canada | 0 (0.0%) | 1 (6.3%) | 0 (0.0%) |
| China | 3 (5.1%) | 0 (0.0%) | 0 (0.0%) |
| Hong Kong | 1 (1.7%) | 0 (0.0%) | 0 (0.0%) |
| Ireland | 1 (1.7%) | 0 (0.0%) | 0 (0.0%) |
| Kenya | 4 (6.8%) | 1 (6.3%) | 0 (0.0%) |
| Multiple countries | 1 (1.7%) | 0 (0.0%) | 1 (7.1%) |
| South Africa | 3 (5.1%) | 0 (0.0%) | 0 (0.0%) |
| United Kingdom | 1 (1.7%) | 0 (0.0%) | 0 (0.0%) |
| USA | 44 (75.6%) | 13 (81.3%) | 12 (85.7%) |
| Target sample size (protocol)/Sample size (primary)** |  |  |  |
| 1-100 | 8 (13.6%) | 9 (56.3%) | 6 (42.9%) |
| 101-200 | 9 (15.3%) | 3 (18.8%) | 1 (7.1%) |
| 201-300 | 7 (11.9%) | 0 (0.0%) | 1 (7.1%) |
| 301-400 | 10 (16.9%) | 0 (0.0%) | 2 (14.3%) |
| 401-500 | 4 (6.8%) | 2 (12.5%) | 1 (7.1%) |
| 501-600 | 2 (3.4%) | 0 (0.0%) | 0 (0.0%) |
| 601-700 | 3 (5.1%) | 0 (0.0%) | 0 (0.0%) |
| 701-800 | 3 (5.1%) | 0 (0.0%) | 0 (0.0%) |
| 801-900 | 3 (5.1%) | 0 (0.0%) | 1 (7.1%) |
| 901-1000 | 4 (6.8%) | 0 (0.0%) | 1 (7.1%) |
| 1000+ | 6 (10.2%) | 2 (12.5%) | 1 (7.1%) |
| Number of randomizations |  |  |  |
| 2 | 55 (93.2%) | 16 (100.0%) | 13 (92.9%) |
| 3 | 3 (5.1%) | 0 (0.0%) | 1 (7.1%) |
| 6 | 1 (1.7%) | 0 (0.0%) | 0 (0.0%) |
| *For studies with both a protocol and primary analysis paper, the publication year is the publication year for the primary analysis paper |  |  |  |
| **For studies with both a protocol and primary analysis paper, the sample size described is the number randomized as reported in the primary analysis paper |  |  |  |

**Table 2** summarizes the populations, interventions, and objectives in the included studies. Most papers focused on adult populations [n=37 (62.7%) protocol papers; n=10 (62.5%) primary analysis papers; n=6 (42.9%) protocol + primary analysis papers]. Additionally, some studies targeted special populations such as Black adolescents, veterans, and females (see **Table S2**). Behavioral and mental health, including substance use, was the most frequently studied therapeutic area [n=23 protocol papers (39.0%); n=12 primary analysis papers (75.0%); n=7 protocol + primary analysis papers (50.0%)], followed by infectious disease [n=8 protocol papers (13.6%); n=1 primary analysis paper (6.3%); n=1 protocol + primary analysis papers (7.1%)], overweight and obesity [n=4 protocol papers (6.8%); n=1 primary analysis papers (6.3%); n=2 protocol + primary analysis papers (14.3%)], and pain [n=4 protocol papers (6.8%); n=0 primary analysis papers (0.0%); n=1 protocol + primary analysis papers (7.1%)]. Other disease areas included cancer, cardiometabolic diseases (including diabetes and cardiovascular disease), inflammatory disorders, and neurological disease or injury.

**Table 2.**
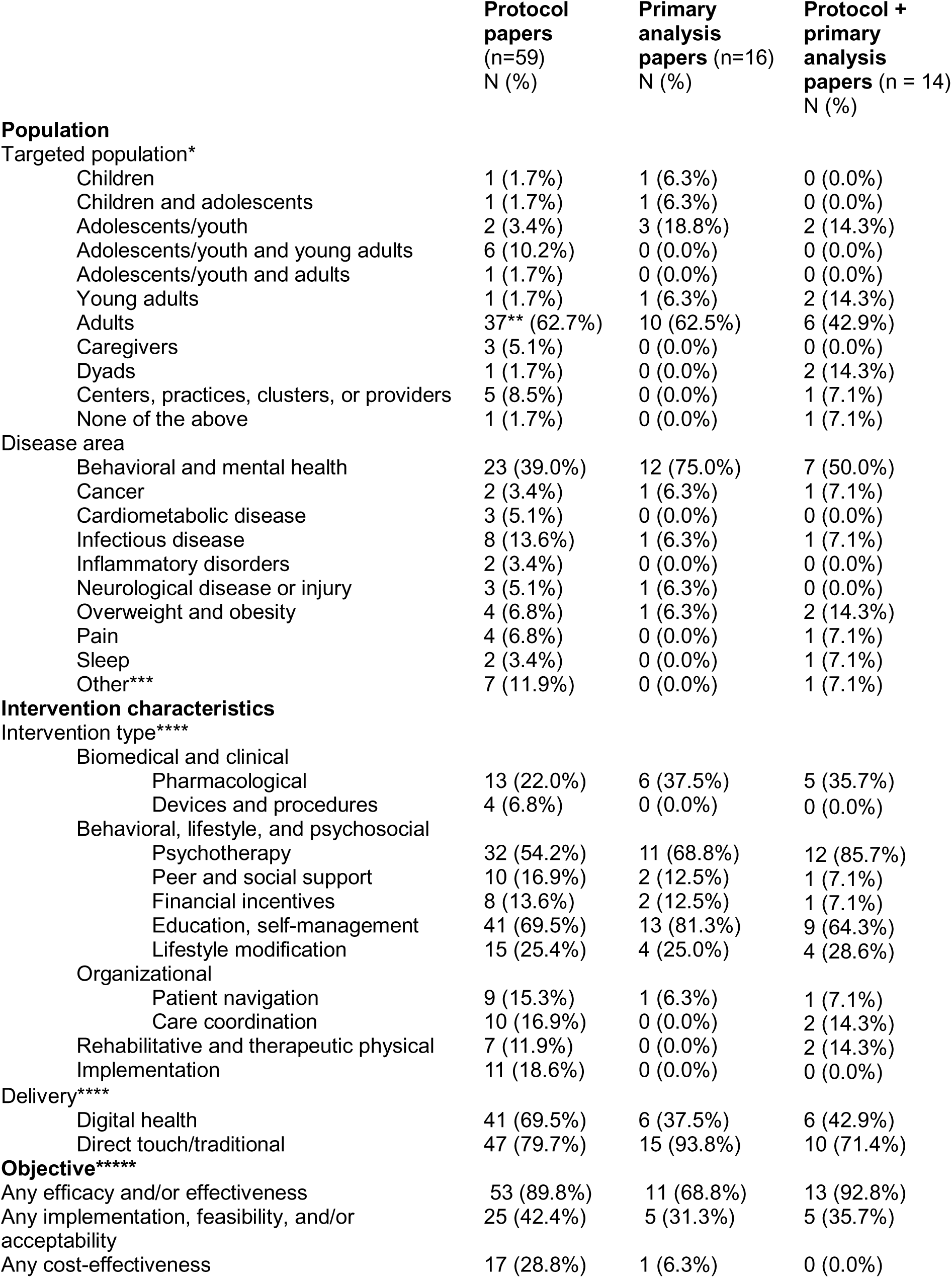
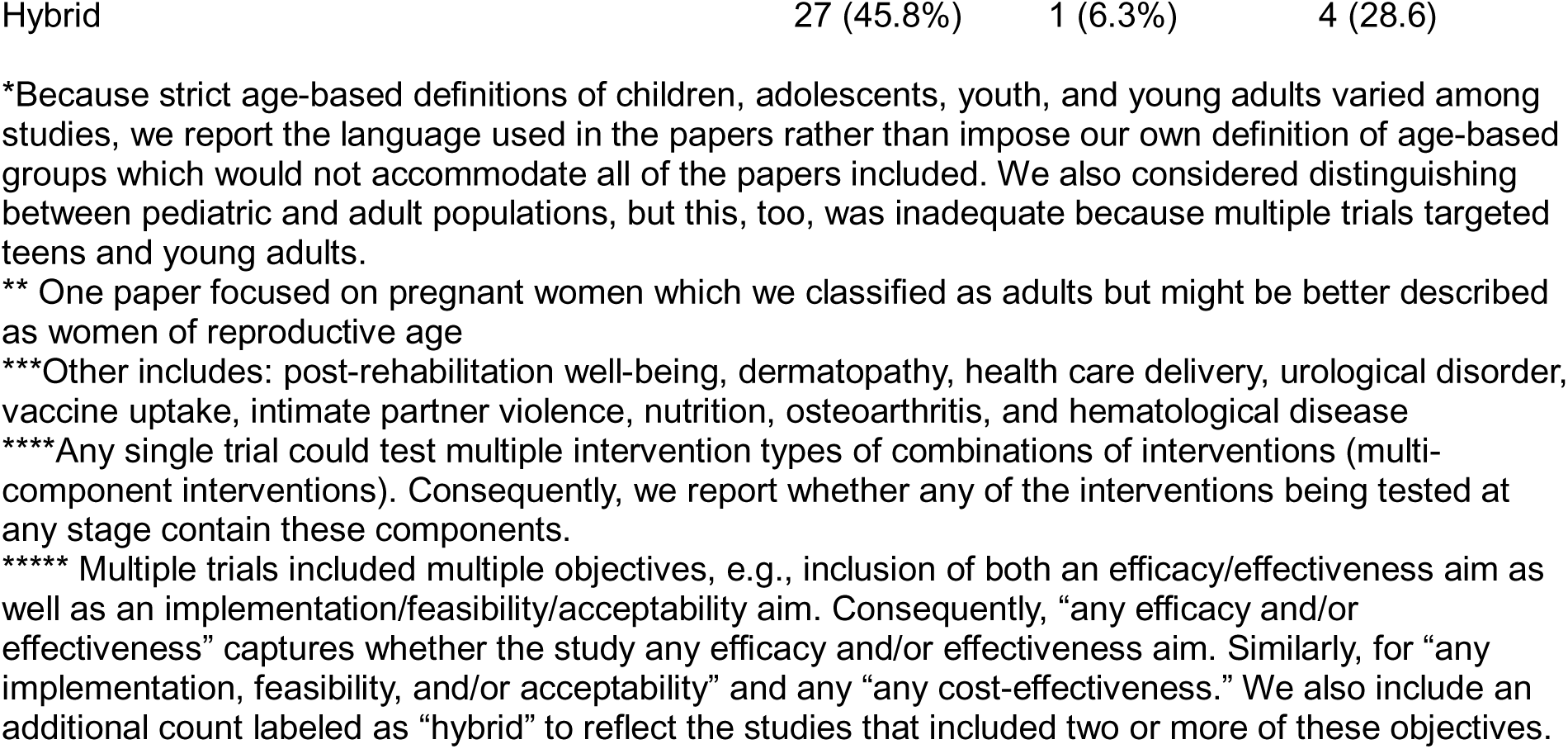
Summary of populations, interventions, and objectives.

Across paper types, the most common types of intervention components were education/self-management, psychotherapy, or pharmacological. Direct touch/traditional delivery of these interventions was used more frequently than digital delivery. Most protocol, primary analysis, and protocol + primary analysis papers evaluated the efficacy and/or effectiveness of the proposed interventions [n=53 (89.8%), n=11 (68.8%), and n=13 (92.8%), respectively]. Primary efficacy outcomes varied widely, reflecting the diverse clinical areas studied. Examples include depression severity (measured by PHQ-9), glycemic control, pain impact scores, viral suppression in HIV studies, adherence rates, and behavioral outcomes such as smoking abstinence or reduction in substance use. Secondary outcomes often addressed complementary clinical measures, patient-reported outcomes, or psychosocial variables, such as anxiety, quality of life, sleep quality, physical activity levels, and patient satisfaction. Twenty-five (42.4%) protocol papers, 5 (31.3%) primary analysis papers, and 5 (35.7%) protocol + primary analysis papers assessed implementation, feasibility, and/or acceptability. Additionally, 18 papers (17 protocol-only and 1 primary-only) evaluated the cost-effectiveness of interventions. Full details are available in **Tables S1 and S2**; our intervention classification strategy is described in **Document S5 and Table S3**.

Re-randomization criteria and outcomes are described at the study-level in **Table S2**. Common criteria for progressing to second-stage interventions included predefined thresholds for clinical or behavioral non-response, such as insufficient symptom improvement, low engagement, or unmet adherence targets. Second-stage interventions typically involved either augmentation of the first-stage intervention with additional components or switching to alternative interventions.

**Table 3** describes reporting on sample size, blinding, and randomization procedures. Most protocol papers (n=55, 93.2%) reported sample size calculations, and among those that did, 80.0% (n=44) accounted for multiple randomizations. Whether or not blinding was discussed was described in approximately two-thirds of protocol (n=38, 64.4%), primary analysis (n=10, 62.5%), and protocol + primary analysis papers (n=11, 78.6%). Randomization details varied; about half of the protocol and protocol + primary analysis papers reported first-stage allocation ratios [n=33 (55.9%) and n=9 (64.3%), respectively] compared to roughly a quarter of primary analysis only papers [n=4 (25.0%)] and fewer reported later-stage allocations for all paper types [n=31 (52.5%) protocol papers; n=3 (18.8%) primary analysis papers; n=6 (42.9%) protocol + primary analysis papers]. Stratification (protocol papers: 45.8%; primary analysis: 18.8%; protocol + primary analysis: 42.9%) and blocking (protocol: 55.9%; primary analysis: 12.5%; protocol + primary analysis: 50.0%) were moderately common, whereas minimization or other balancing approaches were infrequent (protocol: 3.4%; primary analysis: 25.0%; protocol + primary analysis: 14.3%). Some papers (protocol: 11.9%; primary analysis: 37.5%) provided no details on randomization. Reporting rates of these features varied between pilot studies and non-pilot studies. Full details are included in **Table S4**.

**Table 3.** Summary of operational characteristics described.

| Characteristics | Protocol papers<br>(n = 59)<br>N (%) | Primary analysis<br>papers (n = 16)<br>N (%) | Protocol + Primary<br>analysis papers (n = 14)<br>N (%) |
| --- | --- | --- | --- |
| **Sample size** |  |  |  |
| Was a sample size calculation reported? | 55 (93.2%) | 8 (50.0%) | 11 (78.6%) |
| • Pilot studies | 5 (71.4%) | 3 (50.0%) | 1 (25.0%) |
| • Non-pilot studies | 50 (96.2%) | 5 (50.0%) | 10 (100.0%) |
| If reported, did the sample size calculation account for multiple randomizations? | 44 (80.0%*) | 5 (62.5%*) | 7 (63.6%*) |
| • Pilot studies | 4 (80.0%*) | 2 (66.7%*) | 0 (0.0%*) |
| • Non-pilot studies | 40 (80.0%*) | 3 (60.0%*) | 7 (70.0%*) |
| **Blinding/masking** |  |  |  |
| Was blinding or lack of blinding described? | 38 (64.4%) | 10 (62.5%) | 11 (78.6%) |
| • Pilot studies | 4 (57.1%) | 5 (83.3%) | 4 (100.0%) |
| • Non-pilot studies | 34 (73.1%) | 5 (50.0%) | 7 (70.0%) |
| **Randomization** |  |  |  |
| Reported 1<sup>st</sup> stage allocation ratio | 33 (55.9%) | 4 (25.0%) | 9 (64.3%) |
| • Pilot studies | 5 (71.4%) | 3 (50.0%) | 3 (75.0%) |
| • Non-pilot studies | 28 (53.8%) | 1 (10.0%) | 6 (60.0%) |
| Reported 2<sup>nd</sup> stage and/or tertiary stage allocation ratio | 31 (52.5%) | 3 (18.8%) | 6 (42.9%) |
| • Pilot studies | 4 (57.1%) | 2 (33.3%) | 3 (75.0%) |
| • Non-pilot studies | 27 (51.9%) | 1 (10.0%) | 3 (30.0%) |
| Any stratification used? | 27 (45.8%) | 3 (18.8%) | 6 (42.9%) |
| • Pilot studies | 2 (28.6%) | 2 (33.3%) | 1 (25.0%) |
| • Non-pilot studies | 25 (48.1%) | 1 (10.0%) | 5 (50.0%) |
| Any minimization/balancing used? | 2 (3.4%) | 4 (25.0%) | 2 (14.3%) |
| • Pilot studies | 0 (0.0%) | 1 (16.7%) | 0 (0.0%) |
| • Non-pilot studies | 2 (3.8%) | 3 (30.0%) | 2 (20.0%) |
| Any blocking used? | 33 (55.9%) | 2 (12.5%) | 7 (50.0%) |
| • Pilot studies | 5 (71.4%) | 1 (16.7%) | 2 (50.0%) |
| • Non-pilot studies | 28 (53.8%) | 1 (10.0%) | 5 (50.0%) |
| No details reported | 7 (11.9%) | 6 (37.5%) | 0 (0.0%) |
| • Pilot studies | 1 (14.3%) | 2 (33.3%) | 0 (0.0%) |
| • Non-pilot studies | 6 (11.5%) | 4 (40.0%) | 0 (0.0%) |
| * Among those with a first stage sample size calculation |  |  |  |

**Table 4** describes the analysis of embedded DTRs and optimal DTRs. Of the 30 primary analyses reviewed (16 primary analysis-only and 14 primary papers with a protocol paper), 14 (46.7%) analyzed embedded regimes, while none analyzed deeply tailored DTRs. Among the papers that analyzed embedded regimes, the most common statistical method was weighting and replication(14), used in 9 papers (30.0%). See **Table S5** for study-level characterization of DTR analysis approaches.

**Table 4.** Summary of embedded and deeply tailored regimes in primary analysis papers.

| **Characteristic** | **Number of primary analysis papers (n=30)**<br>**N (%)** |
| --- | --- |
| Analyzed embedded regimes | 14 (46.7%) |
| • Pilot studies<br>• Non-pilot studies | 3 (30.0%)<br>11 (55.0%) |
| Statistical methods for embedded regimes |  |
| • Weighting and replication<br>• G-computation<br>• Marginal, weighted least squares<br>• Longitudinal TMLE<br>• GEE-based methods<br>• Cubic polynomial modeling | 9 (30.0%)<br>1 (3.3%)<br>1 (3.3%)<br>1 (3.3%)<br>1 (3.3%)<br>1 (3.3%) |
| Analyzed deeply tailored regimes | 0 (0.0%) |

## DISCUSSION

This comprehensive scoping review of eighty-nine studies, either planned, in process, or completed, reveals both the remarkable versatility of SMARTs across therapeutic domains and significant variability in reporting practices that may limit their full scientific impact.

Our work builds upon and extends previous efforts to characterize SMART reporting. As of April 2025, three reviews of SMARTs have been published, and two additional review protocols have been registered on PROSPERO. The two protocols describe studies that address highly specific questions, such as the methods used to assess SMART designs for psychosocial interventions targeting mental health disorders (15) and SMART designs among individuals with chronic diseases (16). Similarly, the published review by Lorenzoni et al.(6) focused on SMART designs in oncology, and the review by Whiston et al. (17) focused on physical activity interventions. The review published by Bigirumurame et al.(7) primarily discussed deficient reporting of sample size calculation parameters and methods. Our review provides a timely, comprehensive examination of SMARTs across all health domains with a specific focus on an expanded set of design features. For example, while most papers described some aspects of blinding and randomization procedures, critical information, such as the first-stage allocation ratio, was reported only about half the time, and even less frequently for later-stage allocation ratios.

In terms of study design and context, several notable patterns emerge. Most SMARTs have been conducted in the United States (U.S.). This is likely because the SMART methodology has largely been developed within U.S.-based research institutions and due to the considerable clinical research infrastructure required to execute a SMART study fully. Most SMARTs are small or moderately sized with 500 or fewer participants, and most have two randomizations. This may reflect feasibility constraints, as additional randomizations often increase trial duration, complexity, and cost. The majority of studies we analyzed focused on mental and behavioral health. Mental and behavioral health intervention domains may be well-suited for study using a SMART design, given that these interventions can be iterated upon over time in terms of content, intensity, and delivery setting. This approach may be especially promising for the heterogeneous patient populations targeted by trials in this therapeutic domain.

Similar to the previous finding of insufficient sample size reporting (7), our review confirms and extends this observation to additional key trial operational characteristics. In the papers we reviewed, intervention descriptions and re-randomization criteria were consistently well-documented, yet critical design elements, including blinding procedures, randomization allocation ratios, and randomization methods, were often inadequately reported or not reported at all. The inconsistent reporting of crucial SMART elements has significant implications. When essential trial design elements are inadequately described, it becomes difficult for readers and reviewers to assess the trial’s internal validity and methodological rigor. It may also lead to misinterpretation or misapplication of results in the future. For example, incomplete reporting of sample size and power considerations may lead to overstating or understating the strength of evidence for the different hypotheses being tested within a single SMART. This may be particularly impactful when making regime-based comparisons. Poor reporting also hinders secondary analysts’ ability to reproduce findings; for instance, missing allocation ratios or undisclosed randomization procedures prevent the reconstruction of appropriate weights for analyzing embedded treatment sequences. Furthermore, funding agencies, institutional review boards, and regulatory bodies may struggle to assess the feasibility and risk of SMARTs, as well as the evidence generated from SMART data, without a clear reporting framework. Thus, the threat of inconsistent reporting not only weakens scientific communication but also threatens the broader adoption and credibility of this otherwise promising trial design.

One reason reporting for SMARTs is particularly challenging is a notable obstacle we encountered during this review. In our predefined extraction forms, we included fields for primary, secondary, and tertiary (or exploratory) outcomes. However, several papers described primary and secondary aims rather than explicitly stating primary and secondary outcomes. This inconsistency highlights a fundamental complexity in SMART designs, as their unique structure allows multiple research questions to be addressed within a single trial, and each of those questions may correspond to different hypotheses and endpoints. This contrasts with traditional RCTs, which typically have a narrower scope and a single primary hypothesis. SMARTs often involve distinct aims that target fundamentally different research questions and as a result what might be considered a primary outcome for one research question may not be considered the primary outcome when addressing another, obscuring traditional notions of primary, secondary, and tertiary outcomes and analyses. This complexity has important implications beyond outcome reporting; it also raises critical questions about how SMARTs are planned and powered. For example, in an RCT, the sample size is typically calculated to detect a prespecified effect size for a clearly defined primary outcome. In contrast, when SMARTs lack clear alignment among their multiple aims, hypotheses, and outcomes, it becomes unclear which comparisons the sample size calculation is intended to support, and whether one well-powered aim necessarily implies that other aims are also well-powered.

Although the SMART framework inherently facilitates the evaluation of embedded and tailored dynamic treatment regimes (DTRs), our findings suggest that this capability is underutilized in primary analyses. While we did not include secondary analysis papers in our review, the limited exploration of deeply tailored regimes in primary reports points to an opportunity to better capitalize on the full analytical potential of data from SMARTs. Possible reasons for this underuse include risk aversion, limited familiarity with advanced analytical methods, and a lack of explicit methodological guidance. The limited analysis of embedded regimes and lack of analysis of tailored regimes in primary manuscripts represent a missed opportunity to fully leverage SMART data for precision medicine applications, potentially delaying the development of optimized adaptive and personalized interventions for clinical practice.

Unlike traditional randomized controlled trials (CONSORT)(18), systematic reviews (PRISMA)(19), and observational studies (STROBE)(20), which benefit from established reporting guidelines, SMARTs currently lack analogous reporting standards. This absence threatens not only clear communication of study aims, methods, and findings but also limits the ability to properly evaluate the methodological rigor and quality of evidence generated from SMARTs. As our review demonstrates, critical elements such as sample size justification, randomization procedures, and outcome specification are often inconsistently reported, despite being essential for evaluating the validity and generalizability of trial findings.

There is a clear need to develop standardized reporting guidelines specific to SMARTs, e.g., a CONSORT extension for SMARTs. Although the CONSORT extension for adaptive clinical trials (CONSORT-ACE) provides guidance for trials in which the trial conduct is adapted based on accumulating information during the conduct of the trial(21), its scope does not fully address the unique characteristics of SMARTs, fixed trial designs in which interventions that adapt to participant response can be evaluated. For example, prespecified randomizations, explicit definitions of tailoring variables and response criteria, and definitions of embedded treatment regimes fall outside the primary scope of CONSORT-ACE. Accordingly, SMART-specific guidelines would improve the clarity and completeness of reporting, support rigorous study design, and enhance the credibility and reproducibility of this promising trial framework, ultimately accelerating the translation of adaptive interventions into evidence-based clinical practice.

This scoping review has several limitations. Our inclusion criteria were limited to protocol and primary analysis papers published up to February 9, 2024, excluding secondary analyses, registry entries, and methodological papers. Our focus on protocol and primary analysis papers is based on including paper types that characterize how investigators describe the design and report on SMARTs in practice. We acknowledge that our search is now over one year old and that additional SMARTs may have been published between our search and the time of publication of this review. The methodological and reporting practices we identified reflect enduring rather than short-term fluctuations in reporting practices. Given the incremental nature of new SMART publications, we do not anticipate that the inclusion of more recent studies would materially change the thematic conclusions of this review. Additionally, identifying SMART designs through database searches can be challenging due to variability in terminology and inconsistent reporting practices. Thus, some relevant studies might have been inadvertently excluded. Despite these limitations, our findings provide valuable insights into current practices and gaps in SMART reporting and provide the most extensive scoping review of SMARTs to date.

This scoping review provides the most comprehensive examination of SMART reporting practices in human health research to date. Our findings reinforce the need for clear alignment between study aims, outcomes, and analytic strategies; highlight gaps in how fundamental design elements are documented; and identify opportunities to leverage SMARTs more effectively to generate adaptive treatment strategies. To fully realize the promise of SMARTs and the evidence they can generate, reporting guidelines tailored to their unique, innovative design are needed. Establishing such standards would enhance the reproducibility and interpretability of SMARTs, thereby accelerating the adoption of SMART-based evidence in clinical settings.

## FUNDING

KJA, MK, and BR were supported by the National Institutes of Health through the NIH HEAL Initiative under award number 1U24AR076730. MK is additionally supported by the National Institutes of Health under award number UM1TR004406. The content is solely the responsibility of the authors and does not necessarily represent the official views of the NIH.

AK is supported by the National Institutes on Aging, National Institutes of Health, through Grant K01-AG084971-01. The content is solely the responsibility of the authors and does not necessarily represent the official views of the NIH.

SB is supported by the National Institute of Health through the UNC-Chapel Hill pre-doctoral Cardiovascular (CVD) Epidemiology Training Grant (NRSA: T32-HL007055-48). The content is solely the responsibility of the authors and does not necessarily represent the official views of the NIH.

## CONFLICT OF INTEREST

No authors report a conflict of interest with this work.

## AUTHOR CONTRIBUTIONS

NF and BR conceived this review. The protocol was developed by NF, BR, EJ, ARK, KM, MK, AI, and KA. EJ designed the search strategy. NF and BR managed title and abstract screening. NF, SB, CZ, and MH conducted the full-text screening; NF, SB, CZ, MH, and AK extracted the articles. NF and SB synthesized the extraction results and wrote the first draft of the manuscript with KA. All authors provided feedback and approved the final manuscript. NF is the guarantor of this work, and as such, has full access to all the data and takes responsibility for the integrity of the data and the accuracy of the data analysis.

## Data Availability

All data produced in the present study are available upon reasonable request to the authors

## ACKNOWLEDGEMENTS

The authors would like to thank Timothy Feeney, Anthony Dickinson, Alexis Bryant, Abbey Foes, and Annika Cleven for courageously assisting in the development of the screening and extraction process.

## Notes

### Competing Interest Statement

The authors have declared no competing interest.

### Summary of Updates

Major updates include: 1.Improved organization and readability of the tables. In response to reviewer feedback, we have reorganized Tables 1 and 2. Table 1 now exclusively focuses on the context of the studies included in the review (e.g., publication year, country). We have moved what was originally labeled as Table 2 to the supplement (now Table S2) and streamlined it to contain information regarding the population, intervention, comparators, and outcomes (PICO) for each study. Our new Table 2 summarizes our observations regarding the PICO elements of each study. As part of the summarization process, we developed an intervention coding scheme which is detailed in Supplementary Document 5 and Supplementary Table S3. 2.Expanded discussion. We have expanded the discussion to address the important points raised by the reviewers, including practical implications of poor reporting; contextualizing our call for a CONSORT extension for SMARTs to the existing CONSORT extensions, including CONSORT-ACE; and discussion of the geographic concentration of SMARTs. 3.Clarification regarding studies with both a protocol/design paper and a primary analysis paper. Initially, we reported on protocol papers and primary analysis papers. Two of the reviewers raised questions regarding the situation when a study has both a protocol and a primary analysis paper. In response, we have clarified that our unit of analysis for this review is at the study level, not the paper level. We have separately described those studies with both a protocol/design paper and a primary analysis paper, as well as those with just a protocol/design paper and those with just a primary analysis paper. Tables 1, 2, and 3, along with their associated descriptions in the results section, now report on these three categories.

