## Supplementary documents and tables for "Design characteristics of Sequential Multiple Assignment Randomized Trials (SMARTs) for human health: a scoping review of studies between 2009-2024"

Administrative information

Title

Protocol Version

3.0

Registration

[Add text here]

Authors

Contact information

Nikki L. B. Freeman, PhD (corresponding)
Department of Biostatistics and Bioinformatics
Duke Clinical Research Institute
300 W. Morgan St
Durham, NC 27701, United States

Bryce T. Rowland, PhD
Collaborative Studies Coordinating Center
Department of Biostatistics, UNC Gillings School of Global Public Health


Emily P. Jones, MLIS, AHIP
UNC Health Sciences Library


Anna R. Kahkoska, MD, PhD
Department of Nutrition, UNC Gillings School of Global Public Health
Division of Endocrinology and Metabolism, UNC School of Medicine
Center for Aging and Health, UNC School of Medicine


Katharine L. McGinigle, MD, MPH
Department of Surgery, UNC School of Medicine


Michael R. Kosorok, PhD
Department of Biostatistics, UNC Gillings School of Global Public Health


Kevin J. Anstrom, PhD
Collaborative Studies Coordinating Center
Department of Biostatistics, UNC Gillings School of Global Public Health


Contributions

NLBF and BTR generated the idea for this project. They were responsible for concept development and planning. NLBF, BTR, and EPJ were responsible for writing. NLBF, BTR, ARK, KLM, MRK, and KJA provided content expertise; NLBF, EPJ, and ARK provided scoping review methodological expertise; and KJA provided supervision over the protocol development and writing process. All authors read and approved the final protocol manuscript. NLBF is the guarantor of the protocol and will be the guarantor of the subsequent scoping review.

Amendments

Amendments to this protocol will be documented by version number and date (embedded meta-data). We will update the registry in the case of a major revision. Version control will be managed by NLBF and BTR by controlling editorial access to the primary protocol document.

Support

NLBF is supported by a grant from the National Institute on Drug Abuse (NIDA) (R01 DA048764) and an award from the American Diabetes Association (Pathway to Stop Diabetes Program Accelerator Award # 12-22-ACE-18 ).

BTR is supported by a grant from the National Institute of Arthritis & Musculoskeletal and Skin Disease (NIAMS) (U24 AT076730-01 - Back Pain Consortium (BACPAC) Research Program Data Integration, Algorithm Development and Operations Management Center).

The content of this protocol is solely the responsibility of the authors and does not necessarily represent the official views of the National Institutes of Health nor the American Diabetes Association.

**Summary of Protocol Changes**

| Protocol Version Number | Updated Contents | Date of Revision |
| --- | --- | --- |
| 2.0 | - Updated exclusion criteria to include placebo re-randomization designs and extension studies. - Updated Cohen’s Kappa threshold for benchmark screening. | 3/9/2024 |
| 3.0 | - Updated exclusion criteria to exclude trials that sequentially randomize treatments but the authors do not refer to the trial as a SMART | 6/29/2024 |

Introduction

Rationale

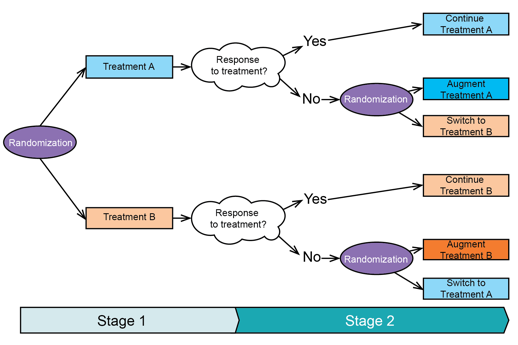
Sequential multiple assignment randomized trials (SMARTs) are increasingly being used to generate “gold standard” data for precision medicine analyses. SMART designs were formalized by Murphy and Bingham (2009), though similar precursor studies were introduced by Lavori and Dawson (2004). These designs mimic clinical decision-making where medical providers and their patients often need to make decisions over time and in response to prior treatment and health history (Kosorok & Moodie, 2015). Operationally, "a SMART is a type of multistage, factorial randomized trial, in which some or all participants are randomized at 2 or more decision points. Whether a patient is randomized at the second or a later decision point, and the available treatment options, may depend on the patient's response to prior treatment” (Kidwell & Almirall, 2023).

For example, a trial participant may initially be randomized to one of two treatments, A or B. After 3 months, they are assessed for response to the initial treatment. If they have a positive treatment response, they may be assigned to remain on treatment. Otherwise, they are randomized to either switch treatment or to an augmented version of their initially assigned treatment. *Figure 1* illustrates this example as one of the most common SMART designs in practice.

*Figure 1. An example of a SMART*

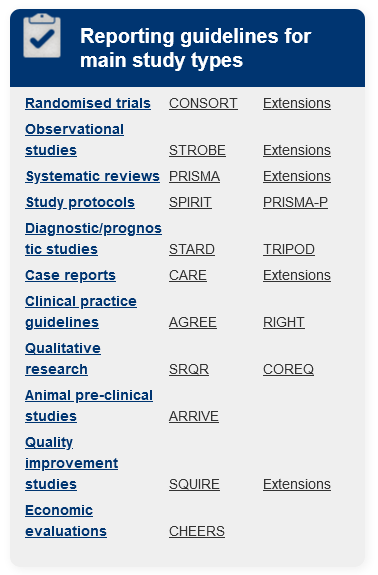
 By formalizing the treatment decision process, SMARTS generate evidence for treatment strategies that can adapt to individuals’ unique, possibly evolving characteristics over the course of treatment. Treatment strategies are often evaluated in a SMART by studying dynamic treatment regimes (DTRs), defined as a sequence of decision rules that map patient characteristics to a recommended treatment decision. One goal of precision medicine is to find DTRs that would optimize a targeted outcome on average in the study population if the DTR were used to assign treatment (Kosorok & Moodie, 2015). Additionally, SMARTs can be used to answer questions regarding main effects of first-line treatments and generating valid ITT inferences (Lavori & Dawson, 2001).

SMARTs are increasingly being adopted in a number of clinical contexts including infectious disease (Evans et al., 2019), oncology (Auyeung et al., 2009), and weight loss (Almirall et al., 2014). Yet, unlike RCTs (Schulz et al., 2010), systematic reviews and meta-analyses (Page et al., 2021), observational studies (von Elm et al., 2008), and other common study designs, no guidelines have been adopted to aid authors in how to report SMART designs and how to report results from the analysis of SMARTs. This lack of reporting guidelines creates a lack of clarity when reporting on a complex, emerging study design for researchers, and presents a challenge for journal editors in evaluating journal submissions reporting on completed SMARTs. Figure 2 lists reporting guidelines for common study types encountered in health research that are often to accompany manuscripts submitted to journals.

Figure 2. Reporting guidelines for common study designs encounter in health research

Identifying how SMARTs are being reported both in terms of design and results is an important first step for understanding what (1) the common practices are and (2) what are the gaps in reporting for this emerging trial design. Currently, only a handful of reviews have been completed regarding SMARTs in human health research. As of September 2023 and to the best of our knowledge, one review has been published and two such reviews have registered protocols on PROSPERO. They focus on highly targeted questions including methodology used to assess SMARTs that use psychosocial interventions for mental health disorders (Bhatti et al., 2023) and the effect of SMART designs among individuals with chronic disease (Kim & Song, 2021). Bigirumurame, Uwimpuhwe and Wason (2022) is a recently published systematic review of SMART reporting that described some of the operational characteristics of SMART designs and focused on the transparency of sample size estimates. The scope of our planned review is wider in terms of the search strategy (e.g., inclusion of grey literature, no disease-specific exclusion criteria) and planned data extraction. Additionally, Bigirumurame, Uwimpuhwe and Wason (2022) did not include ‘sequential multiple assignment randomized trial’ in their search terms. Further, we have designed this review to focus on SMARTs role in advancing the field of precision medicine biostatistics and the developing of repointing guidelines for SMARTs.

Objectives

This review will focus on SMART trials for human health. The goals are to (1) characterize how designs of SMART trials have been described, (2) characterize the research objectives of clinical trials using a SMART design (3) describe how scientific results from SMART trials have been reported, including reporting of recruitment diagrams, adverse events, and results of the analysis of the research objectives.

Methods

Eligibility criteria

1. Inclusion
   a. Papers about SMARTs. SMARTs are trials that have the following operational characteristics:
      i. Fixed trial design
      ii. At least 2 randomizations conducted sequentially over time
      iii. Two or more fixed treatments
      iv. The authors describe the trial using the term “SMART” or “sequential multiple assignment randomized trial.”
   b. Types of Papers
      i. Protocol and design papers
      ii. Primary analysis papers
      iii. Secondary analysis papers
      iv. Trials registered in a clinical trial registry such as clinicaltrials.gov/trial that have reported a planned SMART (protocol, design, and/or statistical analysis plan) or results from a SMART
   c. SMARTs in any human health care setting
   d. Includes pilot SMART trials
   e. Inclusion dates: 2009-2023
2. Exclusion
   a. Non-English articles
   b. Papers about SMARTs not conducted for human health (e.g., SMARTs conducted in animal models, SMARTs conducted in non-medical educational settings)
   c. Reviews, editorials, commentaries, letters, dissertations and theses, systematic reviews, scoping reviews, meta-analyses
   d. Papers about trials that do not randomize participants to interventions.
   e. Papers about trials that do not refer to the trial as a SMART.
   f. Methodological papers, e.g., papers in which the SMART is used as motivation or as an example application, but the objective of the paper is not to report on the results of the trial itself.
   g. Papers about trials that utilize response adaptive randomization in the trial design in which the characteristics of the trial are changing in response to interim analyses, e.g., the probability of assignments to treatments changes in response to interim measures of efficacy.
   h. Papers about trials that have multiple randomizations, but only re-randomize participants who receive a placebo as the first-stage treatment (e.g. placebo run-in trials)
   i. Papers that re-randomize participants receiving active treatment(s) as a part of an extension clinical trial.

Information sources

We will conduct electronic searches for eligible studies within each of the following databases, limited to publication years 2009-present:

- PubMed (National Institutes of Health, National Library of Medicine)
- Scopus (Elsevier)
- EMBASE (Elsevier
- Cochrane Central Register of Controlled Trials (CENTRAL)

Additionally, we will complete forward and backward citation searching on all included studies to further identify those meeting our inclusion criteria using Scopus (Elsevier).

Search strategy

The search strategy has been developed in PubMed and will be translated to Scopus, EMBASE and CENTRAL. The PubMed search strategy can be found below.

**PubMed (NIH/NLM): August 28, 2023**

| **Search** | **Query** | **Number of Results** |
| --- | --- | --- |
| 1 | ("sequential multiple assignment randomized trial"[tiab] OR "sequential multiple assignment randomized trials"[tiab] OR "sequential multiple assignment"[tiab] OR "sequential multiple assignments"[tiab] OR "multiple sequential assignment"[tiab] OR "multiple sequential assignments"[tiab] OR "SMART study"[tiab] OR "SMART studies"[tiab] OR "SMART trial"[tiab] OR "SMART trials"[tiab] OR "SMART design"[tiab] OR "SMART designs"[tiab] OR "multistage treatment"[tiab] OR "multi-stage treatment"[tiab] OR "multistage treatments"[tiab] OR "multi-stage treatments"[tiab] OR "dynamic treatment"[tiab] OR "dynamic treatments"[tiab] OR "multiple random"[tiab] OR multiple-random[tiab] OR "multiple randomization"[tiab] OR "multiple randomisation"[tiab] OR multiple-randomization[tiab] OR multiple-randomisation[tiab] OR "multiple randomizations"[tiab] OR "multiple randomisations"[tiab] OR multiple-randomizations[tiab] OR multiple-randomisations[tiab] OR "multiple assignment"[tiab] OR "multiple assignments"[tiab] OR "second randomization"[tiab] OR "second randomisation"[tiab] OR rerandomize[tiab] OR re-randomize[tiab] OR rerandomise[tiab] OR re-randomise[tiab] OR rerandomized[tiab] OR re-randomized[tiab] OR rerandomised[tiab] OR re-randomised[tiab] OR rerandomization[tiab] OR re-randomization[tiab] OR rerandomisation[tiab] OR re-randomisation[tiab] OR rerandomizations[tiab] OR re-randomizations[tiab] OR rerandomisations[tiab] OR re-randomisations[tiab] OR "double randomization"[tiab] OR "double randomisation"[tiab] OR "double randomizations"[tiab] OR "double randomisations"[tiab] OR "second random assignment"[tiab] OR "treatment sequence"[tiab] OR "treatment sequences"[tiab] OR "treatment sequencing"[tiab] OR "individualized treatment rule"[tiab] OR "individualized treatment rules"[tiab] OR "individualized treatment strategy"[tiab] OR "individualized treatment strategies"[tiab] OR "sequenced treatment"[tiab] OR "sequenced treatments"[tiab] OR "sequential randomized"[tiab] OR "sequential randomised"[tiab] OR "sequential randomization"[tiab] OR "sequential randomisation"[tiab] OR "sequential randomizations"[tiab] OR "sequential randomisations"[tiab] OR cluster-SMART[tiab] OR clustered-SMART[tiab]) | 6,144 |
| 2 | (sequential[tiab]) AND (multiple[tiab] OR assignment[tiab]) AND (random*[tiab]) AND (trial[tiab] or trials[tiab]) | 789 |
| 3 | #1 OR #2 | 6,692 |
| 4 | (clinical[tiab] AND trial[tiab]) OR "clinical trials as topic"[mesh] OR "clinical trial"[pt] OR random*[tiab] OR "random allocation"[mesh] OR "therapeutic use"[sh] | 6,349,836 |
| 5 | #3 AND #4 | 4,645 |
| 6 | "Animals"[Mesh] NOT "Humans"[Mesh] | 5,148,915 |
| 7 | #5 NOT #6 | 4,450 |
| 8 | (review[Publication Type] OR review[ti]) | 3,489,584 |
| 9 | #7 NOT #8 | 3,820 |
| 10 | #9 AND English[language] | 3,748 |
| 11 | #10 AND (2009:2023[pdat]) | 2,872 |
| **Total** | |  |

Study records

Data management

Data will be managed using the Covidence platform, and citations will be managed using EndNote.

Selection process

Prior to the initial abstract review, all of the information retrieval reviewers will conduct a benchmark screening for inclusion/exclusion with 100 titles/abstracts. Inter-rater reliability of the benchmark screening will be conducted. If the inter-rater reliability measured by Cohen’s kappa is >70%, we will proceed with title/abstract screening; if not, we will repeat our training and the benchmark screening process until 70% concordance is reached across all reviewer pairs.

After the benchmark screening process, we will proceed to title/abstract screening. Two independent reviewers will screen titles/abstracts. Covidence will be used to identify conflicting decisions regarding whether a study should move on to a full-text review. Conflicts will be resolved independently by a third reviewer. Reviewers will then repeat another round of screening based on full-text articles. As with title/abstract screening, two reviewers will screen each full text in Covidence, and a third independent reviewer will resolve conflicts if needed. Inter-rater reliability (Cohen’s kappa) will be calculated at the end of the screening process.

Data collection process

Data will be extracted by two independent reviewers using Covidence. After the data is extracted, a third independent reviewer will resolve conflicts. As this is a review to understand how studies are reported and to identify reporting gaps, if any, we will not attempt to contact missing study data from study investigators.

Data items

This review will include three types of papers related to SMARTs: protocol and design papers, primary analysis papers, and secondary analysis papers. A minimal set of data elements we will extract for each type of paper are included in the supplementary table. Broadly, these tables can be summarized in the following categories:

- Paper and Author Details
- Study duration, size, and setting
- Randomization characteristics
- Implementation
- Outcomes
- Aims
- Types of results and how those results were reported
  - Recruitment
  - Adverse Events
  - Methods for and analysis of the study aims
- Funding Information

Outcomes and prioritization

We do not intend to extract outcomes for this review.

Risk of bias in individual studies

As this is a scoping review of SMART design and reporting, not specific clinical outcomes, we do not plan on assessing risk of bias of individual studies.

Data synthesis

We do not plan to synthesize data quantitatively across studies identified for inclusion in our review.

Meta-bias(es)

We do not plan to assess meta-bias(es).

Confidence in cumulative evidence

We do not intend to synthesize evidence across studies.

**Supplementary Document 2: Search strategies**

**Search Strategy Documentation**

Reporting characteristics of Sequential Multiple Assignment Randomized Trials (SMARTs) for human health: a scoping review

### Database Searches

**PubMed (NIH/NLM): February 9, 2024**

| **Search** | **Query** | **Number of Results** |
| --- | --- | --- |
| 1 | ("sequential multiple assignment randomized trial"[tiab] OR "sequential multiple assignment randomized trials"[tiab] OR "sequential multiple assignment"[tiab] OR "sequential multiple assignments"[tiab] OR "multiple sequential assignment"[tiab] OR "multiple sequential assignments"[tiab] OR "SMART study"[tiab] OR "SMART studies"[tiab] OR "SMART trial"[tiab] OR "SMART trials"[tiab] OR "SMART design"[tiab] OR "SMART designs"[tiab] OR "multistage treatment"[tiab] OR "multi-stage treatment"[tiab] OR "multistage treatments"[tiab] OR "multi-stage treatments"[tiab] OR "dynamic treatment"[tiab] OR "dynamic treatments"[tiab] OR "multiple random"[tiab] OR multiple-random[tiab] OR "multiple randomization"[tiab] OR "multiple randomisation"[tiab] OR multiple-randomization[tiab] OR multiple-randomisation[tiab] OR "multiple randomizations"[tiab] OR "multiple randomisations"[tiab] OR multiple-randomizations[tiab] OR multiple-randomisations[tiab] OR "multiple assignment"[tiab] OR "multiple assignments"[tiab] OR "second randomization"[tiab] OR "second randomisation"[tiab] OR rerandomize[tiab] OR re-randomize[tiab] OR rerandomise[tiab] OR re-randomise[tiab] OR rerandomized[tiab] OR re-randomized[tiab] OR rerandomised[tiab] OR re-randomised[tiab] OR rerandomization[tiab] OR re-randomization[tiab] OR rerandomisation[tiab] OR re-randomisation[tiab] OR rerandomizations[tiab] OR re-randomizations[tiab] OR rerandomisations[tiab] OR re-randomisations[tiab] OR "double randomization"[tiab] OR "double randomisation"[tiab] OR "double randomizations"[tiab] OR "double randomisations"[tiab] OR "second random assignment"[tiab] OR "treatment sequence"[tiab] OR "treatment sequences"[tiab] OR "treatment sequencing"[tiab] OR "individualized treatment rule"[tiab] OR "individualized treatment rules"[tiab] OR "individualized treatment strategy"[tiab] OR "individualized treatment strategies"[tiab] OR "sequenced treatment"[tiab] OR "sequenced treatments"[tiab] OR "sequential randomized"[tiab] OR "sequential randomised"[tiab] OR "sequential randomization"[tiab] OR "sequential randomisation"[tiab] OR "sequential randomizations"[tiab] OR "sequential randomisations"[tiab] OR cluster-SMART[tiab] OR clustered-SMART[tiab]) | 6,447 |
| 2 | (sequential[tiab]) AND (multiple[tiab] OR assignment[tiab]) AND (random*[tiab]) AND (trial[tiab] or trials[tiab]) | 839 |
| 3 | #1 OR #2 | 7,017 |
| 4 | (clinical[tiab] AND trial[tiab]) OR "clinical trials as topic"[mesh] OR "clinical trial"[pt] OR random*[tiab] OR "random allocation"[mesh] OR "therapeutic use"[sh] | 6,451,424 |
| 5 | #3 AND #4 | 4,829 |
| 6 | "Animals"[Mesh] NOT "Humans"[Mesh] | 5,192,549 |
| 7 | #5 NOT #6 | 4,634 |
| 8 | (review[Publication Type] OR review[ti]) | 3,575,693 |
| 9 | #7 NOT #8 | 3,981 |
| 10 | #9 AND English[language] | 3,908 |
| 11 | #10 AND ("2009"[Date - Publication] : "3000"[Date - Publication]) | 3,032 |
| **Total** | | 3,032 |

**Scopus (Elsevier): February 9, 2024**

| **Search** | **Query** | **Number of Results** |
| --- | --- | --- |
| 1 | TITLE-ABS ("sequential multiple assignment randomized trial" OR "sequential multiple assignment randomized trials" OR "sequential multiple assignment" OR "sequential multiple assignments" OR "multiple sequential assignment" OR "multiple sequential assignments" OR "SMART study" OR "SMART studies" OR "SMART trial" OR "SMART trials" OR "SMART design" OR "SMART designs" OR "multistage treatment" OR "multi-stage treatment" OR "multistage treatments" OR "multi-stage treatments" OR "dynamic treatment" OR "dynamic treatments" OR "multiple random" OR multiple-random OR "multiple randomization" OR "multiple randomisation" OR multiple-randomization OR multiple-randomisation OR "multiple randomizations" OR "multiple randomisations" OR multiple-randomizations OR multiple-randomisations OR "multiple assignment" OR "multiple assignments" OR "second randomization" OR "second randomisation" OR rerandomize OR re-randomize OR rerandomise OR re-randomise OR rerandomized OR re-randomized OR rerandomised OR re-randomised OR rerandomization OR re-randomization OR rerandomisation OR re-randomisation OR rerandomizations OR re-randomizations OR rerandomisations OR re-randomisations OR "double randomization" OR "double randomisation" OR "double randomizations" OR "double randomisations" OR "second random assignment" OR "treatment sequence" OR "treatment sequences" OR "treatment sequencing" OR "individualized treatment rule" OR "individualized treatment rules" OR "individualized treatment strategy" OR "individualized treatment strategies" OR "sequenced treatment" OR "sequenced treatments" OR "sequential randomized" OR "sequential randomised" OR "sequential randomization" OR "sequential randomisation" OR "sequential randomizations" OR "sequential randomisations" OR cluster-SMART OR clustered-SMART) | 10,411 |
| 2 | TITLE-ABS((sequential) AND (multiple OR assignment) AND (random*) AND (trial OR trials)) | 917 |
| 3 | #1 OR #2 | 11,045 |
| 4 | ( TITLE-ABS ( clinical ) AND TITLE-ABS ( trial ) ) OR INDEXTERMS ( "clinical trials as topic" ) OR INDEXTERMS ( "clinical trial" ) OR TITLE-ABS ( random* ) OR INDEXTERMS ( "random allocation" ) OR INDEXTERMS ( "therapeutic use" ) | 4,242,470 |
| 5 | #3 AND #4 | 6,037 |
| 6 | INDEXTERMS(Animals) AND NOT INDEXTERMS(Humans) | 5,303,151 |
| 7 | #5 AND NOT #6 | 5,882 |
| 8 | #7 AND ( LIMIT-TO ( DOCTYPE , "ar" ) OR LIMIT-TO ( DOCTYPE , "ch" ) OR LIMIT-TO ( DOCTYPE , "bk" ) OR LIMIT-TO ( DOCTYPE , "tb" ) OR LIMIT-TO ( DOCTYPE , "er" ) ) | 4,549 |
| 9 | TITLE(review) | 1,252,552 |
| 10 | #8 AND NOT #9 | 4,499 |
| 11 | #10 AND ( LIMIT-TO ( LANGUAGE , "English" ) ) | 4,291 |
| 12 | #11 AND ( LIMIT-TO ( PUBYEAR , 2009 ) OR LIMIT-TO ( PUBYEAR , 2010 ) OR LIMIT-TO ( PUBYEAR , 2011 ) OR LIMIT-TO ( PUBYEAR , 2012 ) OR LIMIT-TO ( PUBYEAR , 2013 ) OR LIMIT-TO ( PUBYEAR , 2014 ) OR LIMIT-TO ( PUBYEAR , 2015 ) OR LIMIT-TO ( PUBYEAR , 2016 ) OR LIMIT-TO ( PUBYEAR , 2017 ) OR LIMIT-TO ( PUBYEAR , 2018 ) OR LIMIT-TO ( PUBYEAR , 2019 ) OR LIMIT-TO ( PUBYEAR , 2020 ) OR LIMIT-TO ( PUBYEAR , 2021 ) OR LIMIT-TO ( PUBYEAR , 2022 ) OR LIMIT-TO ( PUBYEAR , 2023 ) OR LIMIT-TO ( PUBYEAR , 2024 ) ) | 3,263 |
| **Total** | | **3,263** |

**EMBASE (Elsevier): February 9, 2024**

| **Search** | **Query** | **Number of Results** |
| --- | --- | --- |
| 1 | 'sequential multiple assignment randomized trial':ti,ab OR 'sequential multiple assignment randomized trials':ti,ab OR 'sequential multiple assignment':ti,ab OR 'sequential multiple assignments':ti,ab OR 'multiple sequential assignment':ti,ab OR 'multiple sequential assignments':ti,ab OR 'SMART study':ti,ab OR 'SMART studies':ti,ab OR 'SMART trial':ti,ab OR 'SMART trials':ti,ab OR 'SMART design':ti,ab OR 'SMART designs':ti,ab OR 'multistage treatment':ti,ab OR 'multi-stage treatment':ti,ab OR 'multistage treatments':ti,ab OR 'multi-stage treatments':ti,ab OR 'dynamic treatment':ti,ab OR 'dynamic treatments':ti,ab OR 'multiple random':ti,ab OR multiple-random:ti,ab OR 'multiple randomization':ti,ab OR 'multiple randomisation':ti,ab OR multiple-randomization:ti,ab OR multiple-randomisation:ti,ab OR 'multiple randomizations':ti,ab OR 'multiple randomisations':ti,ab OR multiple-randomizations:ti,ab OR multiple-randomisations:ti,ab OR 'multiple assignment':ti,ab OR 'multiple assignments':ti,ab OR 'second randomization':ti,ab OR 'second randomisation':ti,ab OR rerandomize:ti,ab OR re-randomize:ti,ab OR rerandomise:ti,ab OR re-randomise:ti,ab OR rerandomized:ti,ab OR re-randomized:ti,ab OR rerandomised:ti,ab OR re-randomised:ti,ab OR rerandomization:ti,ab OR re-randomization:ti,ab OR rerandomisation:ti,ab OR re-randomisation:ti,ab OR rerandomizations:ti,ab OR re-randomizations:ti,ab OR rerandomisations:ti,ab OR re-randomisations:ti,ab OR 'double randomization':ti,ab OR 'double randomisation':ti,ab OR 'double randomizations':ti,ab OR 'double randomisations':ti,ab OR 'second random assignment':ti,ab OR 'treatment sequence':ti,ab OR 'treatment sequences':ti,ab OR 'treatment sequencing':ti,ab OR 'individualized treatment rule':ti,ab OR 'individualized treatment rules':ti,ab OR 'individualized treatment strategy':ti,ab OR 'individualized treatment strategies':ti,ab OR 'sequenced treatment':ti,ab OR 'sequenced treatments':ti,ab OR 'sequential randomized':ti,ab OR 'sequential randomised':ti,ab OR 'sequential randomization':ti,ab OR 'sequential randomisation':ti,ab OR 'sequential randomizations':ti,ab OR 'sequential randomisations':ti,ab OR cluster-SMART:ti,ab OR clustered-SMART:ti,ab | 10,826 |
| 2 | (sequential:ti,ab) AND (multiple:ti,ab OR assignment:ti,ab) AND (random*:ti,ab) AND (trial:ti,ab OR trials:ti,ab) | 1,278 |
| 3 | #1 OR #2 | 11,793 |
| 4 | (clinical:ti,ab AND trial:ti,ab) OR 'clinical trial'/exp OR 'clinical trial':ti,ab OR 'clinical trial (topic)'/exp OR 'clinical trial (topic)' OR random*:ti,ab OR 'randomization'/exp OR 'randomization':ti,ab OR 'therapeutic use'/exp | 3,549,140 |
| 5 | #3 AND #4 | 7,587 |
| 6 | [animals]/lim NOT [humans]/lim | 6,462,262 |
| 7 | #5 NOT #6 | 7,378 |
| 8 | 'review'/exp OR 'review':ti OR 'systematic review (topic)'/exp OR 'systematic review'/exp OR 'systematic review':ti | 3,654,312 |
| 9 | #7 NOT #8 | 6,768 |
| 10 | #9 AND [English]/lim | 6,685 |
| 11 | #10 AND [2009-2024]/py | 5,886 |
| 12 | #11 AND ([article]/lim OR [article in press]/lim OR [data papers]/lim OR [erratum]/lim OR [preprint]/lim) | 2,320 |
| **Total** | | **2,320** |

**CENTRAL (Cochrane Library): February 9, 2024**

| **Search** | **Query** | **Number of Results** |
| --- | --- | --- |
| 1 | ("sequential multiple assignment randomized trial" OR "sequential multiple assignment randomized trials" OR "sequential multiple assignment" OR "sequential multiple assignments" OR "multiple sequential assignment" OR "multiple sequential assignments" OR "SMART study" OR "SMART studies" OR "SMART trial" OR "SMART trials" OR "SMART design" OR "SMART designs" OR "multistage treatment" OR "multi-stage treatment" OR "multistage treatments" OR "multi-stage treatments" OR "dynamic treatment" OR "dynamic treatments" OR "multiple random" OR multiple-random OR "multiple randomization" OR "multiple randomisation" OR multiple-randomization OR multiple-randomisation OR "multiple randomizations" OR "multiple randomisations" OR multiple-randomizations OR multiple-randomisations OR "multiple assignment" OR "multiple assignments" OR "second randomization" OR "second randomisation" OR rerandomize OR re-randomize OR rerandomise OR re-randomise OR rerandomized OR re-randomized OR rerandomised OR re-randomised OR rerandomization OR re-randomization OR rerandomisation OR re-randomisation OR rerandomizations OR re-randomizations OR rerandomisations OR re-randomisations OR "double randomization" OR "double randomisation" OR "double randomizations" OR "double randomisations" OR "second random assignment" OR "treatment sequence" OR "treatment sequences" OR "treatment sequencing" OR "individualized treatment rule" OR "individualized treatment rules" OR "individualized treatment strategy" OR "individualized treatment strategies" OR "sequenced treatment" OR "sequenced treatments" OR "sequential randomized" OR "sequential randomised" OR "sequential randomization" OR "sequential randomisation" OR "sequential randomizations" OR "sequential randomisations" OR cluster-SMART OR clustered-SMART):ti,ab,kw | 5,610 |
| 2 | ((sequential) AND (multiple OR assignment) AND (random*) AND (trial OR trials)):ti,ab,kw | 1,677 |
| 3 | #1 OR #2 | 6,925 |
| 4 | #3 with Publication Year from 2009 to present , in Trials | 5,905 |
| 5 | #4 AND Language:English | 5,871 |
| 6 | #5 AND Source: CT.gov (1212) OR Source:ICTRP (225) OR Source:CINAHL (14) | 1,451 |
| **Total** | | **1,451** |

**Supplementary Document 3: Extraction items**

Tab 1: Common data elements

| Column | Column name | Instructions |
| --- | --- | --- |
| Tab 1. Common data elements | | |
| C | SR_number | Do not modify this column |
| D | Item type | Do not modify this column |
| E | Authors | Do not modify this column |
| F | Title | Do not modify this column |
| G | Journal | Do not modify this column |
| H | Full journal | Do not modify this column |
| I | Publication year | Do not modify this column |
| J | Date published | Do not modify this column |
| K | Date accessed | Do not modify this column |
| L | URLs | Do not modify this column |
| M | DOI | Do not modify this column |
| N | Verify Inclusion/Exclusion | Select either Include or Exclude |
| O | Reason for exclusion | If the paper should be excluded, please select a reason why and move on to the next paper. Otherwise, leave this column blank. |
| P | NCT number | Please c/p in the NCT number if it’s given. Otherwise, write NA. |
| Q | Name of associated SMART | If the SMART has a short name, put it here. For example, BEST is the short name for the trial Biomarkers for Evaluating Spine Treatments; the STAR*D trial is the Sequenced Treatment Alternatives to Relieve Depression |
| R | Type of paper | Select one of: Protocol/design, Primary analysis paper, secondary analysis paper, I don’t know If you select you don’t know, ask for help and move on to the next one. |
| S | Sponsor | C/P who paid for the trial, e.g., NIH, industry sponsor |
| Tab 2. Protocol or design papers | | |
| R | Trial objective | What is the primary objective of the trial? |
| S | Pilot? | Is the trial a pilot study? Yes or no |
| T | Country | In which country/countries does/will the trial take place? |
| U | Single/multi site | Is the trial a single- or multi-site trial? |
| V | Trial start date | What date did/will the trial start on? |
| W | Trial end date | What date did/will the trial end on? |
| X | Target population | What is the target population of the trial? |
| Y | Inclusion/exclusion | C/p the inclusion and exclusion criteria for the trial |
| Z | Number of stages | How many randomizations are there? |
| AA | Interventions | Stage 1 interventions: Stage 2 interventions: … |
| AB | Re-randomization criterion | Criterion to proceed to 2^nd^ randomization:  Criterion to proceed to 3^rd^ randomization:  … |
| AC | Primary endpoint | What is the primary endpoint and estimand? |
| AD | Target N | What is the target enrollment? |
| AE | Responder/Nonresponder analysis | Do the authors provide any description of the analyses to support or rationale for the responser/non-responder criterion? |
| AF | Augment/switch/continue | Are the 2^nd^+ randomizations augmentations, switches, or continuations of the previous treatment? e.g., Stage 2 treatment A is an augmentation, Stage 2 treatment B is a swich |
| AG | Sample size calculation | Was a sample size calculation rerpoted: yes or no |
| AH | Sample size and multiple stages | Did the sample size calculation account for the multiple stages of randomization? |
| AI | Sample size methods | What methods were used for the sample size calculation? |
| AJ | Sample size assumptions | Were all the necessary assumptions for the sample size calculation described? E.g., event rates, effect sizes, responder probabilities |
| AK | Primary endpoint(s) | What is the primary endpoint? |
| AL | Secondary endpoint(s) | What are the secondary endpoints? |
| AM | Exploratory endpoint(s) | What are the exploratory endpoints? |
| AN | Blinding | Who, if anyone, was blind to the treatment assignments? |
| AO | Randomization procedures | What randomization procedures were used?  E.g., Stage 1: permuted block, stage 2: stratification |
| AP | Disease | What disease area or health condition is the trial targeting? |
| AQ | Primary analysis objective | What is the objective of the primary analysis? |
| AR | Primary analysis method | What estimation method was used for the primary analysis? |
| AS | Secondary analysis objective(s) | What is the objective(s) of the secondary analysis? |
| AT | Secondary analysis method | What method(s) was used for the secondary analysis? |
| AU | Exploratory endpoint objective | What is the objective of the exploratory analysis/analyses? |
| AV | Exploratory analysis method | What method(s) as used for the exploratory analysis? |
| AW | Ancillary analyses | List any ancillary analyses that are planned |
| AX | Missingness | Does the protocol describe how missing data will be handled? |
| AY | Missingness methods | What methods will be used to handle missing data? |
| AZ | Missingnes sensitivity | Will there be any sensitivity analyses related to missing data? |
| BA | Follow up length | How long is the planned the follow-up period? |
| BB | Sensitivity analyses | Are any additional sensitivity analyses planned? |
| BC | Embedded regimes | Do the authors plan on analyzing any embedded regimes? |
| BD | Embedded regime objective | What is the objective of the analysis of the embedded regimes? |
| BE | Embedded regime methods | What methods will be used to analyze the embedded regimes? |
| BF | Deeply tailored regimes | Do the authors plan on learning deeply tailored regimes? |
| BG | Deeply tailored regimes objective | What is the objective of the analysis of the deeply tailored regimes? |
| BH | Deeply tailored regimes methods | What methods will be used for the analysis of the deeply tailored regimes? |
| BI | Visual schematic | Was a visual representation of the trail schematic included in the paper? |
| Tab 3. Primary analysis papers | | |
| R | Trial objective | What is the primary objective of the trial? |
| S | Pilot? | Is the trial a pilot study? Yes or no |
| T | Country | In which country/countries does/will the trial take place? |
| U | Single/multi site | Is the trial a single- or multi-site trial? |
| V | Trial start date | What date did/will the trial start on? |
| W | Trial end date | What date did/will the trial end on? |
| X | Target population | What is the target population of the trial? |
| Y | Inclusion/exclusion | C/p the inclusion and exclusion criteria for the trial |
| Z | Number of stages | How many randomizations are there? |
| AA | Interventions | Stage 1 interventions: Stage 2 interventions: … |
| AB | Re-randomization criterion | Criterion to proceed to 2^nd^ randomization:  Criterion to proceed to 3^rd^ randomization:  … |
| AC | Primary endpoint | What is the primary endpoint and estimand? |
| AD | Target N | What is the target enrollment? |
| AE | Responder/Nonresponder analysis | Do the authors provide any description of the analyses to support or rationale for the responser/non-responder criterion? |
| AF | Augment/switch/continue | Are the 2^nd^+ randomizations augmentations, switches, or continuations of the previous treatment? e.g., Stage 2 treatment A is an augmentation, Stage 2 treatment B is a swich |
| AG | Sample size calculation | Was a sample size calculation reported: yes or no |
| AH | Sample size and multiple stages | Did the sample size calculation account for the multiple stages of randomization? |
| AI | Sample size methods | What methods were used for the sample size calculation? |
| AJ | Sample size assumptions | Were all the necessary assumptions for the sample size calculation described? E.g., event rates, effect sizes, responder probabilities |
| AK | Primary endpoint(s) | What is the primary endpoint? |
| AL | Secondary endpoint(s) | What are the secondary endpoints? |
| AM | Exploratory endpoint(s) | What are the exploratory endpoints? |
| AN | Blinding | Who, if anyone, was blind to the treatment assignments? |
| AO | Randomization procedures | What randomization procedures were used?  E.g., Stage 1: permuted block, stage 2: stratification |
| AP | Disease | What disease area or health condition is the trial targeting? |
| AQ | Primary analysis objective | What is the objective of the primary analysis? |
| AR | N primary analysis | How many observations were included in the primary analysis? |
| AS | Primary analysis method | What estimation method was used for the primary analysis? |
| AT | Primary analysis uncertainty | Was uncertainty quantified for the primary analysis? |
| AU | Primary analysis uncertainty methods | What method was used to quantify uncertainty for the primary analysis? |
| AV | Secondary analysis objective(s) | What is the objective(s) of the secondary analysis? |
| AW | N secondary analysis | How many observations were included in the secondary analysis/analyses? |
| AX | Secondary analysis method | What method(s) was used for the secondary analysis? |
| AY | Exploratory endpoint objective | What is the objective of the exploratory analysis/analyses? |
| AZ | Exploratory analysis method | What method(s) as used for the exploratory analysis? |
| BA | Ancillary analyses | List any ancillary analyses that were conducted |
| BB | Missingness | Was missingness accounted for in the analyses? |
| BC | Missingness methods | What methods were used to handle missing data? |
| BD | Missing sensitivity | Were there any sensitivity analyses related to missing data? |
| BE | Follow up length | How long was the follow-up period? |
| BF | Retention | What was the retention rate? |
| BG | Sensitivity analyses | Were any additional sensitivity analyses conducted? |
| BH | Embedded regimes | Did the authors analyze any embedded regimes? |
| BI | Embedded regime objective | What is the objective of the analysis of the embedded regimes? |
| BJ | Embedded regime methods | What methods were used to analyze the embedded regimes? |
| BK | Deeply tailored regimes | Did the authors learn/analyze deeply tailored regimes? |
| BL | Deeply tailored regimes objective | What is the objective of the analysis of the deeply tailored regimes? |
| BM | Deeply tailored regimes methods | What methods were used for the analysis of the deeply tailored regimes? |
| BN | Visual schematic | Was a visual representation of the trail schematic included in the paper? |
| BO | EID descriptors | Did the authors use appropriate population descriptors such as ancestry, geographic and sociodemographic characteristics of all participants, particularly those in under-represented groups? |
| BP | Inclusion/exclusions EID implications | Did the authors describe the implications of inclusion and/or exclusion of people who are understudied in precision medicine research or underserved by health services? For example, did the authors describe implications for successful extrapolation of study findings to other groups, particularly those typically under-represented in precision medicine research |
| BQ | PPIE in research | Did the authors describe patient and public involvement and engagement (PPIE) in any aspect of the study design, conduct and/or reporting |
| BR | PPIE in contextualization | Did the authors, where possible, and ideally with guidance from patient and public involvement and engagement (PPIE) representatives, describe the potential impact of the study’s results from a lived-experience perspective, especially the impact of the research on people living with disease? |
| BS | N(%) female | From “Table 1”, how many women and what percent of the analyzed population were women |
| BT | Race/ethnicity | From “Table 1”, give the N and % of each racial and ethnic category |
| BU | SOGI | From “Table 1”, give the N and % of each sexual orientation and gender identity reported |
| BV | Education | Give the N and % of individuals in each education category |
| BW | Income | Give the N and % of individuals in each income category |

**Supplementary Document 4: PRISMA-ScR Checklist**

**Preferred Reporting Items for Systematic reviews and Meta-Analyses extension for Scoping Reviews (PRISMA-ScR) Checklist**

| **SECTION** | **ITEM** | **PRISMA-ScR CHECKLIST ITEM** | **REPORTED ON PAGE #** |
| --- | --- | --- | --- |
| **TITLE** | | | |
| Title | 1 | Identify the report as a scoping review. | 1 |
| **ABSTRACT** | | | |
| Structured summary | 2 | Provide a structured summary that includes (as applicable): background, objectives, eligibility criteria, sources of evidence, charting methods, results, and conclusions that relate to the review questions and objectives. | 2 |
| **INTRODUCTION** | | | |
| Rationale | 3 | Describe the rationale for the review in the context of what is already known. Explain why the review questions/objectives lend themselves to a scoping review approach. | 4-5 |
| Objectives | 4 | Provide an explicit statement of the questions and objectives being addressed with reference to their key elements (e.g., population or participants, concepts, and context) or other relevant key elements used to conceptualize the review questions and/or objectives. | 5 |
| **METHODS** | | | |
| Protocol and registration | 5 | Indicate whether a review protocol exists; state if and where it can be accessed (e.g., a Web address); and if available, provide registration information, including the registration number. | 5 |
| Eligibility criteria | 6 | Specify characteristics of the sources of evidence used as eligibility criteria (e.g., years considered, language, and publication status), and provide a rationale. | 5; Supplementary Document 1 |
| Information sources* | 7 | Describe all information sources in the search (e.g., databases with dates of coverage and contact with authors to identify additional sources), as well as the date the most recent search was executed. | 5-6 |
| Search | 8 | Present the full electronic search strategy for at least 1 database, including any limits used, such that it could be repeated. | Supplementary File 2 |
| Selection of sources of evidence† | 9 | State the process for selecting sources of evidence (i.e., screening and eligibility) included in the scoping review. | 6 |
| Data charting process‡ | 10 | Describe the methods of charting data from the included sources of evidence (e.g., calibrated forms or forms that have been tested by the team before their use, and whether data charting was done independently or in duplicate) and any processes for obtaining and confirming data from investigators. | 6 |
| Data items | 11 | List and define all variables for which data were sought and any assumptions and simplifications made. | Supplementary Table 3 |
| Critical appraisal of individual sources of evidence§ | 12 | If done, provide a rationale for conducting a critical appraisal of included sources of evidence; describe the methods used and how this information was used in any data synthesis (if appropriate). | NA |
| Synthesis of results | 13 | Describe the methods of handling and summarizing the data that were charted. | 6 |
| **RESULTS** | | | |
| Selection of sources of evidence | 14 | Give numbers of sources of evidence screened, assessed for eligibility, and included in the review, with reasons for exclusions at each stage, ideally using a flow diagram. | 6-7; Figure 2 |
| Characteristics of sources of evidence | 15 | For each source of evidence, present characteristics for which data were charted and provide the citations. | Tables S1, S2, S3, S4, and S5 |
| Critical appraisal within sources of evidence | 16 | If done, present data on critical appraisal of included sources of evidence (see item 12). | NA |
| Results of individual sources of evidence | 17 | For each included source of evidence, present the relevant data that were charted that relate to the review questions and objectives. | Tables S1, S2, S3, S4, and S5 |
| Synthesis of results | 18 | Summarize and/or present the charting results as they relate to the review questions and objectives. | 6-9; Tables 1, 2, 3, 4 |
| **DISCUSSION** | | | |
| Summary of evidence | 19 | Summarize the main results (including an overview of concepts, themes, and types of evidence available), link to the review questions and objectives, and consider the relevance to key groups. | 9-12 |
| Limitations | 20 | Discuss the limitations of the scoping review process. | 11 |
| Conclusions | 21 | Provide a general interpretation of the results with respect to the review questions and objectives, as well as potential implications and/or next steps. | 12 |
| **FUNDING** | | | |
| Funding | 22 | Describe sources of funding for the included sources of evidence, as well as sources of funding for the scoping review. Describe the role of the funders of the scoping review. | 12 |

JBI = Joanna Briggs Institute; PRISMA-ScR = Preferred Reporting Items for Systematic reviews and Meta-Analyses extension for Scoping Reviews.

* Where *sources of evidence* (see second footnote) are compiled from, such as bibliographic databases, social media platforms, and Web sites.

† A more inclusive/heterogeneous term used to account for the different types of evidence or data sources (e.g., quantitative and/or qualitative research, expert opinion, and policy documents) that may be eligible in a scoping review as opposed to only studies. This is not to be confused with *information sources* (see first footnote).

‡ The frameworks by Arksey and O’Malley (6) and Levac and colleagues (7) and the JBI guidance (4, 5) refer to the process of data extraction in a scoping review as data charting*.*

§ The process of systematically examining research evidence to assess its validity, results, and relevance before using it to inform a decision. This term is used for items 12 and 19 instead of "risk of bias" (which is more applicable to systematic reviews of interventions) to include and acknowledge the various sources of evidence that may be used in a scoping review (e.g., quantitative and/or qualitative research, expert opinion, and policy document).

*From:* Tricco AC, Lillie E, Zarin W, O'Brien KK, Colquhoun H, Levac D, et al. PRISMA Extension for Scoping Reviews (PRISMAScR): Checklist and Explanation. Ann Intern Med. 2018;169:467–473. [doi: 10.7326/M18-0850](http://annals.org/aim/fullarticle/2700389/prisma-extension-scoping-reviews-prisma-scr-checklist-explanation).

**Supplementary Document 5. Intervention component coding definitions**

| Domain | Intervention type | Intervention subtype | Definition |
| --- | --- | --- | --- |
| Intervention type | Biomedical/clinical interventions | Pharmacologic | Interventions that are defined by direct manipulation of a drug, supplement, or biological therapy. This includes starting, switching, or combining medications, as well as, varying doses, formulation, or pharmacological classes. This excludes situations interventions aimed at improving medication adherence. |
| Intervention type | Biomedical/clinical interventions | Devices and procedures | Devices, procedures, manipulation of physical and chemical devices |
| Intervention type | Behavioral, lifestyle, and psychosocial interventions | Psychological and psychotherapy | Interventions grounded in formal psychological theory or clinical psychotherapy frameworks, aimed at changing thoughts, emotions, or behaviors through a structured therapeutic process. Examples of structured processes include cognitive behavioral therapy, motivational interviewing, mindfulness-based stress reduction, acceptance and commitment therapy, behavioral activation, and interpersonal psychotherapy. |
| Intervention type | Behavioral, lifestyle, and psychosocial interventions | Peer and social support | Interventions utilizing trained peers or social networks to provide emotional support, shared experiences, and practical guidance |
| Intervention type | Behavioral, lifestyle, and psychosocial interventions | Financial and economic interventions | Incentives, cash, lottery, vouches, subsidies |
| Intervention type | Behavioral, lifestyle, and psychosocial interventions | Education, self-management, and adherence | Interventions focused on increasing knowledge, skills, and self-efficacy to manage one’s own health condition. Typically characterized by information/skill-building, coaching, disease management, symptom monitoring, and adherence behavior. Interventions may be based on health behavior theories such as the health belief model or social cognitive theory. |
| Intervention type | Behavioral, lifestyle, and psychosocial interventions | Lifestyle modification | Voluntary interventions that include dietary and nutritional counseling, exercise programs, smoking cessation, sleep hygiene, and weight management. |
| Intervention type | Organizational interventions | Patient navigation | Interventions in which a navigator provides personalized assistance to help patients overcome modifiable barriers to healthcare access and treatment completion. This includes helping with scheduling appointments, arranging transportation, assisting with insurance, providing language interpretation and cultural mediation, help with completing forms and paperwork, and connection with financial assistance. Notably, navigation activities work with and for individual patients to overcome barriers. |
| Intervention type | Organizational interventions | Care coordination | Interventions that organize and management patient care activities across multiple providers, settings, and timepoints to ensure appropriate, timely, and continuous care delivery. Care coordination focuses on information transfer and communication among all team members involved in a patient’s care. |
| Intervention type | Rehabilitative and therapeutic physical interventions | Rehabilitation and therapeutic physical interventions | Therapeutic interventions focused on improving, maintaining, or restoring physical function, mobility, or physical health through movement, manual techniques, physical modalities, or adaptive equipment. This includes physical therapy, occupational therapy, manual therapy (massage, manipulation, mobilization), physical modalities (ultrasound, heat/cold therapy), rehabilitation programs, and acupuncture. This excludes interventions focused on the psychological aspects of rehabilitation, exercise advice without a structured program, surgical procedures for repair or reconstruction, and general exercise and physical activity programs. |
| Intervention type | Implementation | Implementation | Interventions designed to promote the adoption, integration, and sustainability of practices into routine healthcare. Typically these are focused on changing provider behavior, organizational readiness, or system capacity rather than directly targeting patient outcomes. Strategies might include training, audit/feedback, and facilitation. |
| Delivery | Delivery | Digital health | Interventions delivered via SMS, app, or telehealth, as well as web modules, wearable devices, clinical decision support systems, and web-based intervention platforms. Digital health interventions are classified here only when the technology is essential to the intervention mechanism and not merely a communication mechanism. |
| Delivery | Delivery | Direct touch | Interventions delivered through direct interactions between healthcare providers and patients or self-administered by patients using established face-to-face or manual methods without requiring digital technology. Characterized by the use of conventional clinical settings and roles. This includes face-to-face clinical encounters, group sessions, self-administered treatments, traditional outreach programs (home visits, community-based programs), standard referrals, and paper-based tools. |

**TABLE S1. Study characteristics, objectives, and rationale for SMART design**

| **SR_number** | **First author, publication year** | **Study objectives or aims** | **Trial start and end dates (year)** | **Trial location (country)** | **Target enrollment/**  **number of participants randomized to first stage intervention** | **Pilot** | **Efficacy and/or effective-ness** | **Implementation, feasibility, and/or acceptability** | **Cost-effective-ness** |
| --- | --- | --- | --- | --- | --- | --- | --- | --- | --- |
| **Protocol papers** | | | | | | | | | |
| 1296 | Abuogi LL et al., 2023(1) | Evaluate which individual interventions and dynamic sequence of combined interventions (e.g., strategies) improve sustained viral suppression and HIV care engagement in adolescents and young adults living with HIV (AYAH). | NA | Kenya | 880 | No | Yes | No | Yes |
| 1105 | Arean P et al., 2021(2) | To compare the effectiveness of message-based care, videoconference-psychotherapy, and a combination of the two treatments in depressed adults. | 2021-2024 | USA | 1000 | No | Yes | No | No |
| 1247 | August GJ et al., 2016(3) | Develop adaptive treatment strategies to reduce the risk of first-time offenders developing serious conduct problems and becoming chronic offenders. | NA | USA | 100 | Yes | No | Yes | No |
| 1072 | Auyeung SF et al., 2009(4) | To propose a design for utilizing a sequential, multiple assignment, randomized trial design for patients with malignant melanoma to test the relative efficacy of drugs that target serotonin versus dopamine metabolism during 4 weeks of intravenous, then 8 weeks of subcutaneous, interferon-alpha therapy. | NA | USA | 70 | No | Yes | No | No |
| 1102 | Bahraini NH et al., 2020(5) | To examine the effectiveness of an adaptive implementation strategy to improve the uptake of suicide risk screening and evaluation in Veterans Health Administration (VHA) ambulatory care settings. | 2020- | USA | 140 | No | Yes | Yes | No |
| 1290 | Belzer ME et al., 2018(6) | Compare adaptive mobile health (mHealth) interventions that could increase antiretroviral therapy (ART) adherence among youth living with HIV (YLH) aged 15 to 24 years. | 2018-2020 | USA | 190 | No | Yes | Yes | Yes |
| 1129 | Berget C et al., 2019(7) | To test four interventions that couple developmentally tailored behavioral supports with education to optimize use of diabetes devices and reduce psychosocial distress for parents of young children with type 1 diabetes (T1D). | NA | USA | 90 | No | Yes | No | No |
| 1035 | Buchholz SW et al., 2020(8) | Determine the most effective adaptive intervention to increase physical activity (steps, moderate-to-vigorous physical activity) and improve cardiovascular health among employed women who are not regularly physically active. | NA | USA | 312 | No | Yes | No | Yes |
| 1082 | Carr E et al., 2024(9) | To construct an adaptive physical activity (PA) intervention that will subsequently be evaluated against treatment-as-usual using a standard two-arm trial design. This will allow us to determine the optimum sequence of embedded treatments to improve PA in community-based people who are independently mobile (with or without a mobility aid), who are no longer receiving inpatient or outpatient or PA rehabilitation. | 2023-2024 | Ireland | 117 | No | No | Yes | No |
| 1094 | Comins CA et al., 2019(10) | Evaluate the effectiveness , durability, and cost-effectiveness of different nurse-led adaptive HIV treatment interventions for cisgender female sex workers living with HIV in Durban, South Africa. | 2018-2020 | South Africa | 800 | No | Yes | Yes | Yes |
| 1147 | Davis-Ewart L et al., 2023(11) | Feasibility, acceptability and preliminary effectiveness of distinct combinations of telehealth MI and CM in 70 cisgender sexual minority men who use stimulants that are not currently taking PrEP. | NA | USA | 70 | Yes | Yes | Yes | No |
| 1182 | Doorenbos AZ et al., 2023(12) | To evaluate guided relaxation and acupuncture to improve pain control in sickle cell disease, to determine the most appropriate and effective treatment sequence for any given patient based on their unique characteristics, and to describe the processes and structures required to implement guided relaxation and acupuncture within health care systems. | NA | USA | 366 | No | Yes | Yes | No |
| 1065 | Drake CL et al., 2022(13) | To determine the effectiveness of digital cognitive-behavioral therapy for insomnia (dCBT-I) alone and in combination with clinician-led CBT-I for insomnia and the prevention of MDD incidence and relapse. | 2018-2023 | USA | 1000 | No | Yes | No | No |
| 1044 | Edelman EJ et al., 2021(14) | To identify the optimal adaptive approach involving first-line tobacco medications and contingency management (CM) to promote exhaled carbon monoxide (eCO)-confirmed smoking abstinence and its impact on HIV-specific outcomes. | 2020- | USA | 632 | No | Yes | Yes | No |
| 1036 | Eldridge-Smith ED et al., 2022(15) | The main objective of this study is to determine if augmentation of usual care PAP therapy with online CBT-I (OCBT-I) improves insomnia and OSA outcomes, as well as to determine the added benefit of providing a higher intensity, second stage, therapist-led CBT-I (TCBT-I) to patients who demonstrate sub-optimal short-term outcomes with OCBT-I. | NA | USA | 384 | No | Yes | No | No |
| 1101 | Fernandez ME et al., 2020(16) | To evaluate multi-level implementation strategies to increase the reach and impact of a tobacco cessation treatment and to evaluate characteristics of healthcare system, providers, and patients that may influence tobacco-use outcomes. | NA | USA | 6000 | No | Yes | Yes | No |
| 1053 | Flynn D et al., 2018(17) | Determine the optimal treatment combination, sequence, and duration of standard rehabilitative care (SRC) and complementary and integrative health (CIH) therapies among active duty service members with chronic pain; and identify predictors (e.g., biomarkers) of positive treatment response. | NA | USA | 280 | No | Yes | No | No |
| 1047 | Fox CK et al., 2024(18) | Examine when to start anti-obesity medication (AOM) (specifically phentermine) in adolescents who are not responding to lifestyle therapy and how to modify AOM when there is a sub-optimal response to the initial pharmacological intervention (specifically, for phentermine non-responders, is it better to add topiramate to phentermine or switch to topiramate monotherapy). | NA | USA | 150 | No | Yes | No | No |
| 1073 | Fritz JM et al., 2020(19) | Compare the effectiveness of different nonpharmacological treatments for chronic low back pain in the military health system. | NA | USA | 1200 | No | Yes | Yes | No |
| 1111 | Fu SS et al., 2017(20) | To test whether incomplete responders to initial phase of tobacco longitudinal care (TLC) treatment benefit from the addition of medication therapy management (MTM). | NA | USA | 1000 | No | Yes | No | No |
| 1019 | Germeroth LJ et al., 2019(21) | To determine the optimal sequence of prenatal and postpartum lifestyle interventions to optimize maternal weight, cardiometabolic health, and psychosocial outcomes at 12 months postpartum. | 2016- | USA | 300 | No | Yes | No | No |
| 1176 | Hassett AL et al., 2023(22) | To perform an interventional response phenotyping study in a cohort of chronic low back pain (cLBP) patients and to show that currently available, clinically derived measures, can predict differential responsiveness to treatments for cLBP. | NA | USA | 400 | No | Yes | No | No |
| 1159 | Hibbard JC et al., 2018(23) | Investigate and evaluate the efficacy of certain sequences of laser treatment on hypertrophic burn scars. | NA | USA | 180 | No | Yes | No | No |
| 1185 | Inwani I et al., 2017(24) | To optimize engagement of adolescent girls and young women (AGYW) in both the HIV prevention and care continuum and to determine the recruitment and testing strategies that identify the highest proportion of previously undiagnosed HIV infections. | 2017-2019 | Kenya | 108 | Yes | Yes | Yes | Yes |
| 1145 | Jain S et al., 2023(25) | To determine the anti-inflammatory therapy from first randomization with the lowest rate of second randomization in the treatment of Multisystem Inflammatory Syndrome in Children (MIS-C) and evaluate the best order in which the therapies should be given to achieve the greatest therapeutic effect. | NA | USA | 180 | No | Yes | No | No |
| 1104 | Johnson JE et al., 2018(26) | To determine the minimum necessary intervention to maintain a postpartum depression prevention program in prenatal clinics serving low-income women. | NA | USA | 90 | No | Yes | Yes | Yes |
| 1103 | Kilbourne AM et al., 2014(27) | To determine, among sites not initially responding to Replicating Effective Programs (REP), the effect of adaptive implementation strategies that begin with an External Facilitator (EF) or with an External Facilitator plus an Internal Facilitator (IF) on improved evidence-based practices (EBP) use and patient outcomes in 12 months. | 2014- | USA | 80 | No | Yes | Yes | Yes |
| 1170 | Kopelowicz A et al., 2023(28) | To describe an adaptive intervention that integrates community mental health workers, diabetes nurse educators, family members, and patients as partners in care while promoting diabetes self-management for Mexican American individuals with Type 2 diabetes mellitus, and to determine what sequence of intervention strategies works most efficiently and for whom. | 2017-2021 | USA | 330 | No | Yes | No | No |
| 1084 | Kor PP et al., 2023(29) | To develop and identify a two-stage adaptive intervention with prespecified rules guiding whether, how or when to offer different interventions initially/over time to reduce depressive symptoms in family caregivers of people with dementia (FG-of-PWD). | 2022-2025 | China | 272 | No | Yes | No | No |
| 1179 | Levy R et al., 2019(30) | To identify evidence-based strategies for first line and second-line treatment for mental disorders delivered by non-specialists integrated with primary care, to investigate presumed mediators of treatment outcome, and to determine patient-level moderators of treatment effect to inform personalized, resource-efficient, non-specialist treatments and sequencing. | NA | Kenya | 2710 | No | No | Yes | No |
| 1071 | Li X et al., 2021(31) | To compare the effectiveness of commonly used antipsychotic drugs in first episode of schizophrenia (FES) patients using a SMART design, which is based on the combination of sequential therapy and dynamic therapy. | NA | China | 720 | No | Yes | No | Yes |
| 1070 | Lion KC et al., 2023(32) | Compare two discrete implementation strategies for improving interpreter use: (1) enhanced education targeting intrapersonal barriers to use delivered in a scalable format (interactive web-based educational modules) and (2) a strategy targeting system barriers to use in which mobile video interpreting is enabled on providers’ own mobile devices. | NA | USA | 55 primary care providers, 648 patients | No | Yes | Yes | Yes |
| 1083 | Liu H et al., 2021(33) | To develop a post-discharge suicide intervention strategy based on BCIs and evaluate its implementability under the implementation outcome framework. | 2021-2023 | USA | 312 | No | Yes | Yes | Yes |
| 1131 | Markland AD et al., 2023(34) | To increase access to behavioral treatment of urinary incontinence for women Veterans by comparing the effectiveness of two virtual care delivery modalities. | 2020-2023 | USA | 286 | No | Yes | No | No |
| 1106 | Micheletti RG et al., 2020(35) | To determine the optimal management of patients with chronic skin-limited vasculitis. | 2017-2020 | USA, Canada, Japan, and other countries | 90 | Yes | Yes | No | No |
| 1038 | Nelson B et al., 2018(36) | To test outcomes of ultra-high risk (UHR) patients, primarily functional outcome, in response to a sequential intervention strategy consisting of support and problem solving (SPS), cognitive-behavioral case management and antidepressant medication. | NA | Australia | 500 | No | Yes | No | No |
| 1199 | O'Keefe VM et al., 2019(37) | Evaluate which brief interventions, alone or in combination, have the greater effect on suicide ideation (primary outcome) and resilience (secondary outcome) among AI youth ages 10–24 ascertained for suicide-related behaviors by the tribal surveillance system. | NA | USA | 304 | No | Yes | No | No |
| 1034 | Osilla KC et al., 2023(38) | To develop an adaptive concerned partners (CP) intervention to decrease CP drinking and increase service member (SM) help-seeking. | NA | USA | 530 | No | Yes | No | No |
| 1018 | Peter SC et al., 2023(39) | Compare two psychological interventions targeting buprenorphine-naloxone adherence for persons with opioid use disorder: (1) contingency management (CM)  and (2) brief motivational interviewing plus substance-free activities session plus mindfulness (BSM). | 2022-2023 | USA | 280 | No | Yes | No | No |
| 1079 | Peterson BS et al., 2021(40) | Evaluate the sequencing of cognitive behavioral therapy (CBT) and fluoxetine medication in the treatment of pediatric anxiety disorders. | NA | USA | 404 | No | Yes | No | No |
| 1045 | Pfammatter AF et al., 2019(41) | To identify the optimal first line obesity treatment, testing the hypothesis that app alone will be non-inferior to app plus coaching, measured by weight change from baseline to 6 months. | NA | USA | 400 | No | Yes | No | Yes |
| 1251 | Quanbeck A et al., 2020(42) | Provide guidance on how best to sequence and combine implementation strategies for opioid prescribing to meet the needs of different primary care clinics. | 2020- | USA | 256 clinician prescribers (40-45 clinics) | No | Yes | Yes | Yes |
| 1088 | Rabin BA et al., 2023(43) | Optimize a multicomponent health program to promote COVID-19 vaccine uptake and engagement in preventive healthcare using our established co-creation approach to address multi-level (individual, community, systemic) barriers to vaccine uptake and preventive services engagement. | NA | USA | 300 | No | Yes | Yes | No |
| 1030 | Sabri B et al., 2021(44) | Evaluate an adaptive, trauma-informed, culturally tailored technology-delivered intervention tailored to the needs of immigrant women who have experienced intimate partner violence (IPV). | NA | USA | 1266 | No | Yes | No | No |
| 1166 | Schlechter CR et al., 2023(45) | To evaluate the feasibility and acceptability of dissemination strategies designed to increase the reach of evidence-based interventions for weight management. | 2022-2023 | USA | 200 | Yes | No | Yes | No |
| 1140 | Skolasky RL et al., 2020(46) | To evaluate evidence-based, protocol-driven treatments using physical therapy, cognitive behavioral therapy, or mindfulness to examine comparative effectiveness and optimal sequencing for patients with chronic low back pain. | NA | USA | 945 | No | Yes | Yes | No |
| 1063 | Smith SK et al., 2021(47) | The objective of this trial is to optimize the delivery of an mHealth intervention for cancer survivors with posttraumatic stress symptoms using a Sequential Multiple Assignment Randomized Trial (SMART) design. | NA | USA | 400 | No | Yes | No | No |
| 1027 | Sripada RK et al., 2023(48) | Test the effectiveness of initiating treatment with prolonged exposure for primary care (PE-PC) versus clinician-supported (CS) post-traumatic stress disorder (PTSD) coach and test the effectiveness of second-stage strategies (continue or step up to Full PE) for slow responders. | NA | USA | 430 | No | Yes | Yes | Yes |
| 1146 | Tamura K et al., 2020(49) | To determine if a SMART-designed adaptive mHealth intervention with remote messaging tailored to neighborhood-environment resources will increase physical activity more than standard remote messaging. | 2021-2023 | USA | 180 | No | Yes | Yes | No |
| 1076 | van Heerden A et al., 2023(50) | To determine whether community-based ART initiation and maintenance increase the proportion of ART-eligible persons living with HIV who achieve viral suppression at 18 months. | NA | South Africa | 900 | No | Yes | No | Yes |
| 1286 | Velloza J et al., 2022(51) | Test a stepped model of scalable adherence support strategies of young South African women who initiate PrEP. | 2019 – 2022 | South Africa | 500 | No | Yes | Yes | No |
| 1281 | Walton MA et al., 2023(52) | Test the efficacy of adaptive interventions designed to reduce alcohol misuse and violent behaviors among adolescents and emerging adults visiting the emergency department (ED) | 2018 – 2022 | USA | 700 | No | Yes | No | No |
| 1057 | Wan Y et al., 2023(53) | To establish a collaborative care model (CCM) for home-cared older adults with dementia to determine the best strategy to deliver collaborative care, including both the channel and frequency of delivery. | 2023-2025 | China | 358 dyads | No | Yes | No | Yes |
| 1167 | Wilroy J et al., 2023(54) | To test the feasibility of a tele-exercise program that applies an adaptive intervention design for 30 adults with spinal cord injury (SCI), targeting increases in adherence to the exercise program and physical activity participation. | 2022-2023 | USA | 40 | Yes | No | Yes | No |
| 1141 | Windsor L et al., 2022(55) | To optimize an adaptive intervention that will increase rates of testing and adherence to New Jersey State COVID-19 recommendations among high-risk populations and to identify predictors of testing completion and adherence to those recommendations. | 2021-2022 | USA | 582 | No | Yes | No | No |
| 1123 | Yan X et al., 2022(56) | To assess the feasibility of using the custom-built, native menu planner mobile application supported by nutrition coaching in improving carbohydrate periodization behavior. | 2020-2021 | United Kingdom | 900 | Yes | No | Yes | Yes |
| 1278 | Zhao SZ et al., 2022(57) | Evaluate the effectiveness of mHealth intervention in increasing smoking cessation | NA | Hong Kong | 1200 | No | Yes | No | No |
| 1279 | Zhou G et al., 2020(58) | Design optimal adaptive combinations of vector control interventions to maximize reductions in malaria burden based on local malaria transmission risks, vector ecology, and the available mix of interventions approved by the Ministry of Health of Kenya. | 2020 – 2022 | Kenya | 36 clusters, each with a population of about 500 participants | No | Yes | No | Yes |
| 1155 | Zullig LL et al., 2021(59) | To determine an optimal intervention that will improve cancer patient outcomes. | NA | USA | 800 | No | Yes | No | No |
| **Primary analysis papers** | | | | | | | | | |
| 1099 | Butzer JF et al., 2023(60) | To compare the effectiveness of two different interventions that promote physical activity in individuals with traumatic spinal cord injury (SCI) and determine the effect of relapse prevention | NA | USA | 79 | No | Yes | No | No |
| 1282 | Czyz EK et al., 2021(61) | Investigate the feasibility and acceptability of a SMART for adolescents at elevated suicide risk following psychiatric hospitalization | 2019- | USA | 80 | Yes | No | Yes | No |
| 1271 | Fatori D et al., 2018(62) | Test the effect of beginning treatment of childhood OCD with fluoxetine (FLX) or group cognitive-behavioral therapy (GCBT) accounting for treatment failures over time | 2010 – 2013 | Brazil | 83 | No | Yes | No | No |
| 1081 | Gao K et al., 2020(63) | To sequentially study the effectiveness of lithium and divalproex monotherapy and adjunctive therapy with quetiapine or lamotrigine in the acute and continuation treatment of bipolar I or II disorder at any phase of illness and at least mild symptom severity | 2011-2016 | USA | 112 | No | Yes | No | No |
| 1273 | Geng EH et al., 2023(64) | Evaluate 15 adaptive sequences using previously established interventions including counseling, SMS messages, conditional cash transfers (CCT), and peer navigators | 2015 – 2019 | Kenya | 1809 | No | Yes | No | No |
| 1124 | Kruse GR et al., 2023(65) | To test the feasibility of a SMART to deliver a stepped-care intervention among primary care patients who smoked daily | 2020 | USA | 35 | Yes | No | Yes | No |
| 1269 | Lambert SD et al., 2022(66) | To assess the feasibility and acceptability of using a Sequential Multiple Assignment Randomized Trial (SMART) to optimize the delivery of a web-based, stress management intervention for patients with a cardiovascular disease (CVD) | NA | Canada | 59 | Yes | No | Yes | No |
| 1203 | McKay JR et al., 2015(67) | Evaluate the effect of providing choice of treatment alternatives to patients who fail to engage in or drop out of intensive outpatient programs (IOPs) for substance dependence. | 2008 – 2012 | USA | 500 | No | Yes | No | No |
| 1200 | Morgenstern J et al., 2021(68) | To address gaps in knowledge about effective adaptive interventions for alcohol use disorder (AUD), the current study aimed to test drinking outcome effects of stepped care treatment for individuals with AUD. | NA | USA | 160 | No | Yes | No | No |
| 1078 | Naar-King S et al., 2016(69) | To develop an adaptive behavioral treatment for African American adolescents with obesity | NA | USA | 18 | No | Yes | No | No |
| 1020 | Pelham Jr WE et al., 2016(70) | To address whether endpoint outcomes are better depending on which treatment is initiated first and, in case of insufficient response to initial treatment, whether increasing dose of initial treatment or adding the other treatment modality is superior | 2006-2009 | USA | 152 | No | Yes | No | Yes |
| 1219 | Pistorello J et al., 2017(71) | The potential to utilize adaptive treatment strategies for treating moderate to severe suicidal risk among college students, including feasibility and acceptability findings. | NA | USA | 62 | Yes | No | Yes | No |
| 1062 | Sauer-Zavala S et al., 2022(72) | To collect preliminary data regarding the feasibility, acceptability, and potential effectiveness of personalizing the selection and sequencing of cognitive behavioral therapy (CBT) skills from the Unified Protocol (UP) | NA | USA | 70 | Yes | No | Yes | No |
| 1005 | Schlam TR et al., 2024(73) | Compare the effects of different smoking cessation interventions on smoking abstinence among individuals who had relapsed following an initial quit attempt | 2015-2019 | USA | 1154 | No | Yes | No | No |
| 1075 | Sikorskii A et al., 2023(74) | To test optimal sequencing of two evidenced-based interventions for symptom management | 2018- | USA | 451 | No | Yes | No | No |
| 1002 | Stanger C et al., 2020(75) | Examine if (a) adding working memory training to contingency management (CM) for youth with cannabis use disorder (CUD) and (b) switching nonresponding youth to higher magnitude CM incentives boosts outcomes | 2015-2018 | USA | 59 | Yes | Yes | No | No |
| **Protocol + primary analysis papers** | | | | | | | | | |
| Primary: 1232  Protocol: 1032 | Primary: Fortney JC et al., 2021(76)  Protocol: Fortney JC et al., 2020(77) | Compare two clinic-to-clinic interactive video approaches to delivering evidence-based mental health treatments to patients in primary care clinics. | 2016 – 2019 | USA | Protocol sample size estimate: 1000  Primary participants included: 1004 | No | Yes | No | No |
| Primary: 1014  Protocol: 1054 | Primary: Gonze BB et al., 2020(78)  Protocol: Morais Pereira Simões et al., 2019(79) | Investigate the feasibility of the SMART design for assessing the effects of a smartphone app intervention to improve physical activity in adults; describe the participants’ perception regarding the protocol and the use of the app for physical activity qualitatively | NA | Brazil | Protocol sample size estimate: N/A  Primary participants included: 18 | No | Yes | Yes | No |
| Primary: 1223  Protocol: 1122 | Primary: Gunlicks-Stoessel M et al., 2019(80)  Protocol: Gunlicks-Stoessel M et al., 2016(81) | Compare two time points (week 4 and week 8) for assessing symptoms during interpersonal psychotherapy for depressed adolescents (IPT-A) and explore four algorithms that used symptom assessments to select the subsequent treatment | NA | USA | Protocol sample size estimate: 32  Primary participants included: 40 | Yes | Yes | No | No |
| Primary: 1008  Protocol: 1297 | Primary: Igudesman D et al., 2023(82)  Protocol: Corbin KD et al., 2022(83) | Evaluate the effect of a hypocaloric low carbohydrate, hypocaloric moderate low fat, and Mediterranean diet without calorie restriction on weight and glycaemia in young adults with T1D and overweight or obesity | 2018-2021 | USA | Protocol sample size estimate: 72  Primary participants included: 38 | Yes | Yes | Yes | No |
| Primary: 1177  Protocol 1239 | Primary: Karp JF et al., 2019(84)  Protocol: Karp JF et al., 2016(85) | Evaluate the effect of cognitive behavioral therapy (CBT) and physical therapy (PT), together with the temporal ordering of these interventions, on patient reported global impression of change (P-GIC), mood, anxiety, and pain, and compare the strategies’ impact on incidence of common psychiatric disorders over 12-months | 2012-2016 | USA | Protocol sample size estimate: 136  Primary participants included: 99 | Yes | Yes | Yes | No |
| Primary: 1216  Protocol: 1299 | Primary: Morin CM et al., 2020(86)  Protocol: Morin CM et al., 2016(87) | To evaluate the comparative efficacy of 4 treatment sequences involving psychological and medication therapies for insomnia and examine the moderating effect of psychiatric disorders on insomnia outcomes. | 2012 – 2017 | Canada and USA | Protocol sample size estimate: 224  Primary participants included: 211 | No | Yes | No | No |
| Primary: 1298   Protocol: 1193 | Primary: Mustanski B et al., 2023(88)  Protocol: Mustanski B et al., 2020(89) | To assess the impact of the newly-adapted SMART Sex Ed on 18 HIV and sexual health-related variables (e.g., attitudinal, motivational, and behavioral outcomes). We also aimed to explore differences in intervention effects across key groups, including differences in demographics (e.g., race/ethnicity, age) and live experiences (e.g., SES, rural, ever had anal sex prior to enrollment) | 2018-2020 | USA | Protocol sample size estimate: 1878  Primary participants included: 983 | No | Yes | No | No |
| Primary: 1154  Protocol: 1080 | Primary: Patrick ME et al., 2021(90)  Protocol: Patrick ME et al., 2020(91) | To develop a universal and resource-efficient adaptive preventive intervention (API) for incoming first-year students as a bridge to indicated interventions to address alcohol-related risks | 2019-2020 | USA | Protocol sample size estimate: 675  Primary participants included: 891 | No | Yes | No | No |
| Primary: 1228  Protocol: 1077 | Primary: Schmitz JM et al., 2024(92)  Protocol: Schmitz JM et al., 2018(93) | Test an adaptive intervention for optimizing abstinence outcomes over phases of treatment for cocaine use disorder using a SMART design. | NA | USA | Protocol sample size estimate: 160  Primary participants included: 118 | No | Yes | No | No |
| Primary: 1121  Protocol: 1125 | Primary: Schoenfelder EN et al., 2019(94)   Protocol: Chronis-Tuscano A et al., 2016(95) | To evaluate the feasibility and acceptability of sequencing medication and behavioral treatments for mothers with ADHD to target outcomes, including maternal ADHD, parenting, and child ADHD symptoms/impairment in multiplex ADHD families | NA | USA | Protocol sample size estimate: 40 dyads  Primary participants included: 35 mothers | Yes | No | Yes | No |
| Primary: 1246  Protocol 1245: | Primary: Sherwood NE et al., 2022  Protocol : Sherwood NE et al., 2016 | Determine the optimal time to identify state-of-the-art behavioral weight loss treatment (SBT) and whether it is better to switch suboptimal responders to portion-controlled meals (PCM) or acceptance-based treatment (ABT). | 2015 – 2017 | USA | Protocol sample size estimate: 500  Primary participants included: 468 | No | Yes | No | No |
| Primary: 1112  Protocol: 1274 | Primary: Smith SN et al., 2022(96)  Protocol: Kilbourne AM et al., 2018(97) | Development of a school-level adaptive implementation strategy for adopting and scaling up school professionals (SP) delivery of cognitive behavioral therapy (CBT) | 2018-2020 | USA | Protocol sample size estimate: 100 (≥200 SPs)  Primary participants included: 94 schools (169 SPs) | No | Yes | Yes | No |
| Primary: 1250  Protocol: 1137 | Primary: Somers TJ et al., 2023(98)  Protocol : Kelleher SA et al., 2017(99) | Evaluate whether varying doses of Pain Coping Skills Training (PCST) and response-based dose adaptation can improve pain management in women with breast cancer | 2016 – 2020 | USA | Protocol sample size estimate: 327  Primary participants included: 327 | No | Yes | No | No |
| Primary: 1097  Protocol: 1012 | Primary: Wyatt G et al., 2021(100)  Protocol: Sikorskii et al., 2017(101) | To investigate the optimal sequencing of two evidence‐based complementary therapies to determine whether it is best to use only one therapy or start with one and add another based on demonstrated need | NA | USA | Protocol sample size estimate: 331 dyads  Primary participants included: 347 dyads | No | Yes | No | No |

**Table S2. Populations, interventions, and outcomes**

| First author, year | Target population | Age group | Disease area | Number of stages | First stage interventions | Criterion for proceeding to 2^nd^ stage randomization | Second and tertiary stage interventions | Primary efficacy outcome(s) | Secondary efficacy outcome(s) | Other outcome(s) |
| --- | --- | --- | --- | --- | --- | --- | --- | --- | --- | --- |
| Protocol papers |  |  |  |  |  |  |  |  |  |  |
| Abuogi LL et al., 2023(1) | Adolescents and young adults living with HIV | Adolescents and young adults | HIV | 2 | - Standard of Care – Routine Education & Counseling (SOC-REC) - Electronic peer navigation (eNAV) | Non-responders had a lapse in engagement, defined as either a missed clinic visit by >=14 days or HIV viral load >= 1000 copies/ml  Non-responders were re-randomized to second-stage interventions | Responders **continue** their stage 1 intervention in stage 2  All non-responders were re-randomized to:   - **Augment** or **switch** to standard of care with outreach and intensified counseling (SOC-OIC) - **Switch** to conditional cash transfer (CCT) for re-engagement - **Switch** to in-person peer navigation (IP-NAV) | Engagement, viral suppression | Time to return for subset of patients with lapse due to missed visits, time to viral-re-suppression for those who lapsed through an elevated viral load, retention alone, medication possession ratio, HIV RNA levels | Cost-effectiveness |
| Arean P et al., 2021(2) | Adults with depression | Adults | Mental and behavioral health | 2 | - Video conferencing-based psychotherapy (VCP) - Message-based psychotherapy (MBP) | Non-responders failed to make a 50% reduction in PHQ-9 scores at 5-weeks  Non-responders were re-randomized to second-stage interventions | Responders **continue** their stage 1 intervention in stage 2  All non-responders are re-randomized to an **augmented** treatment:   - Weekly VCP+MBP - Monthly VCP+MBP | PHQ-9 (depression) and SDS | NA | Generalized Anxiety Scale (GAD-7), intervention intensity, working alliance (WAI-SR), timeliness of treatment, Experience of Care and Health Outcomes Survey (ECHO) |
| August GJ et al., 2016(3) | Youth (13 to 17 years of age) identified by law enforcement as early-stage offenders and referred to pre-court juvenile diversion programming. | Youth | Mental and behavioral health | 2 | - TI-Brief: Youth-focused motivational interviewing and cognitive behavioral therapy - EP-Brief: Parent-focused use of the EcoFIT family intervention model | Non-responders were youth showing evidence of any of the following: (a) conduct behaviors in the at-risk range or higher on the Behavioral Assessment System for Children–2 (i.e., a T-score of 60 or above), or (b) impaired functioning as rated by practitioners in the School, Home, Community=Delinquency, or Behavior Towards Others domains on the CAFAS, or (c) elevated exposure to deviant peer influences on the Friendship scale (i.e., a T-score of 60 or above on either scale)  Non-responders were re-randomized to second-stage interventions | Responders are s**tepped down** and monitored over time for the maintenance of intervention effects  Non-responders are randomized to either:   - **Augment** the first stage intervention with additional content/training (TI-Extended or EP-Extended) - **Switch** to the alternative extended intervention option | NA | NA | Primary and secondary efficacy endpoints were not specified  Target endpoints: Feasibility endpoints |
| Auyeung SF et al., 2009(4) | Individuals with completely resected T4 Stage III or IV malignant melanoma at a single clinical center who will receive IFN-alpha treatment | Adults | Cancer | 2 | - Escitalopram - Methylphenidate (MPH) | Non-responders were those with a Hamilton Depression Rating (HAM-D) score is >12  Non-responders were re-randomized to second-stage interventions | Responders **continue** their stage 1 intervention in stage 2  Non-responders are re-randomized to either:   - Stage 1 treatment **augmented** with the other treatment - **Switched** to other treatment | Adherence to 12 weeks of IFN-alpha treatment (number of IFN-alpha treatments tolerated and study drop-out rate due to all causes) | Mood symptoms (depression, anxiety, and irritability), neurovegetative symptoms (fatigue, anorexia, altered sleep, and psychomotor slowing) | Evaluation of augmentation versus switching for non-remitters |
| Bahraini NH et al., 2020(5) | Veterans Affairs Medical Centers (VAMCs) | Centers, practices, or providers | Mental and behavioral health | 2 | - Audit and Feedback (A/F) - Implementation as usual (IAU) & Follow-up | Non-responders in the A/F group: failure to complete secondary screening and comprehensive suicide risk evaluation (CSRE) for at least 80% of eligible patients  A/F responders and non-responders were re-randomized to stage 2 interventions | Stage 1 IAU and follow-up sites **continue** IAU and follow-up.  Stage 1 A/F sites who implemented adequately are re-randomized to either:   - **Continue** A/F - **Switch** to IAU   Stage 1 A/F sites who did not implement adequately are re-randomized to either:   - **Continue** A/F - **Augment** A/F with facilitation | C-SSRS Screener uptake, C-SSRS uptake | Clinical performance measures that reflect key practice elements of risk identification: secondary suicide screening and the CSRE | Clinical impact endpoints, implementation (reach, sustainability, fidelity, barriers/facilitators, organizational factors) |
| Belzer ME et al., 2018(6) | Youth living with HIV (YLH) aged 15-24 | Youth and young adults | HIV | 2 | - CPS: Cell-phone support including daily voice calls - SMS: Text-message support including personalized text reminders | Non-responders had a viral load greater than 200 copies/mL or were missing at 3 months  Responders and non-responders were re-randomized to stage 2 interventions | Responders were re-randomized to either:   - **Stepped Down** intervention: same mode delivered 2x/week - **Switch** to Standard of care: no further intervention   Non-responders were re-randomized to either:   - CPS-I: CPS **augmented** with incentives - SMS-I: SMS **augmented** with incentives | Viral load | Self-reported medication adherence | Cost-effectiveness, the 5 components of the Self-Management Model over time, barriers and facilitation |
| Berget C et al., 2019(7) | Parents of young children 2-6 years old with T1D | Caregivers | Diabetes | 6 | - Two interventions focused on optimizing adherence   - Developmental Demands: education on diabetes technology and problem-solving skills   - Distress Reduction - Two interventions focused on glucose control   - Fear of Hypoglycemia (behavioral based)   - Remote Monitoring | Adherence target: sensor wear at least 6 of 7 days on average per week or <= 4 missed days in the previous 4 weeks  Glucose target: sensor glucose values in the range of 70-180 mg/dl at least 60% of the time    Non-responders were re-randomized to second-stage interventions | Participants who meet both targets are not randomized to second stage intervention  Participants who fail to achieve adherence target or both targets are randomized to either **switch** or **augment** to:   - No intervention - Developmental Demands - Distress Reduction   Participants who fail to achieve the glucose target are randomized to either **switch** or **augment** to:   - No intervention - Fear of Hypoglycemia - Remote monitoring | Glycemic control, quality of life | Psychosocial variables to assess quality of life (health related quality of life, parents’ general depression and anxiety symptoms, T1D-related distress, hypoglycemia worries) | Feasibility |
| Buchholz SW et al., 2020(8) | Employed women who are not regularly physically active | Adults | Mental and behavioral health | 2 | - Enhanced physical activity monitor: wearable activity monitor and mobile app with goal setting and physical activity prescription - Enhanced physical activity monitor + motivational text messages | Non-responders fail to wear the Fitbit or does not have a valid day (wearing the Fitbit a minimum of 3 of 7 days in weeks 6-8, with either 10 hours of wear time or exceeding baseline step average) or does not exceed the short-term goal of 600 steps above baseline average for two of the three weeks in weeks 6 to 8  Non-responders were re-randomized to second-stage interventions | Responders **continue** their stage 1 intervention in stage 2  Non-responders are randomized to **augment** their stage 1 intervention one of two interventions**:**   - Personal calls: motivational interviewing calls - Group meetings: structured group meetings for peer support | Physical activity (number of steps/day, minutes of moderate/vigorous physical activity/week) and aerobic fitness (V02max) | Self-reported physical activity behavior (International Physical Activity Questionnaire (IPQQ) - Long 7 Days Self-Administered), BMI, and waist circumference | Cost-effectiveness |
| Carr E et al., 2024(9) | Post-stroke community-dwelling individuals who are able to mobilize (with or without a mobility aid) | Adults | Post-rehabilitation well-being | 2 | - Structured exercise (EX) - Lifestyle physical activity (LPA) | Non-responders: average 7-day step count does not meet the short-term goal of 5% more than the 7-day average from the previous week for 2 of the 3 weeks in weeks 4-6 OR fails to wear their Fitbit or does not have a valid day (ie, wears the Fitbit a minimum of 3 of 7 days in weeks 4-6, with either 10 hours of wear time for that day or exceed the target step count for that day). | Responders **continue** their same stage 1 intervention for another 6 weeks.  Non-responders are re-randomized to:   - **Continue** stage 1 intervention - **Switch** to other intervention | Mean steps/day over 7days measured using the Fitbit Charge 4 on the non-paretic limb | Sedentary behavior (Sedentary Behavior Questionnaire), Fatigue (Fatigue Severity Scale (FSS)), Quality of Life (Stroke-Specific Quality of Life Scale (SS-QOL), EuroQol-5 Dimension-5 Levels (EQ-5D-5L)), Depression and Anxiety (Hospital Anxiety and Depression Scale (HADS)), Activities of Daily Living (Reintegration into Normal Living Index (RNLI), Physical Activity Self-Efficacy (Short Self-Efficacy for Exercise Scale (SSEE)), Cognitive Function (Cognitive Assessment Scale for Stroke Patients (CASP)), Stroke recurrence, Adverse events (death, falls) | Feasibility (recruitment rates, retention and adherence rates, time required for participant enrollment, recruitment site capacity), fidelity (intervention design fidelity, training fidelity, delivery fidelity, receipt fidelity, enactment fidelity) |
| Comins CA et al., 2019(10) | Female sex workers living with HIV | Adults | HIV | 2 | - Decentralized treatment program (DTP) - Individualized Case Management (ICM) | Non-responders are those with viral load ≥50 copies/mL at 6 months  Responders and non-responders are re-randomized in stage 2 | Responders are re-randomized to:   - **Switch** to standard of care (SoC) - **Continue** stage 1 intervention   Non-responders are re-randomized to:   - **Continue** stage 1 intervention - **Augment** stage 1 intervention with other intervention | Combined intention-to-treat outcome of retention in ART care and viral suppression at 18 months | Retention and viral suppression at 18 months among month 6 non-responders randomized to continuation of either intervention versus combined interventions, Risk factors of uncontrolled viremia and/or loss to follow-up, Durability of retention and viral suppression among 6-month responders continuing decentralized treatment or case management versus those randomized to revert to South African standard of care, Self-reported adherence and ART refill data to assess adherence across arms, Viral suppression by arms among retained participants, Loss-to-follow-up across arms, Participant reported intervention acceptability, Characterize and compare resistance across arms, Comparison of intervention cost-effectiveness according to order or intervention delivery and duration of intervention received | Cost-effectiveness, implementation (decentralized treatment deliveries, case management phone-based contacts, case management in-person meetings) |
| Davis-Ewart L et al., 2023(11) | Sexual minority men who use stimulants and are not taking PrEP | Adults | HIV | 2 | - Motivational Interviewing (MI): 2-session MI intervention focused on PrEP use and concomitant stimulant use or condomless anal sex - Contingency management (CM): Financial incentives for PrEP clinical evaluation and filling a PrEP prescription | Responders filled a prescription for PrEP at 3 months | Non-responders to MI were randomized to either:   - Assessment only - Switch to CM   Non-responders to CM were randomized to either:   - Assessment only - MI | Filled a PrEP prescription | PrEP clinical evaluation, stimulant use severity, receptive and insertive anal sex, PrEP intentions, PrEP self-efficacy, PrEP acttitudes, PrEP stigma | Feasibility, experience |
| Doorenbos AZ et al., 2023(12) | Adults with sickle cell disease | Adults | Sickle cell disease | 2 | - Acupuncture - Guided relaxation - Usual care | Responder/non-responder definition depends on pain impact score (not otherwise specified) | Non-responders are randomized to either:   - **Continue** first-stage therapy or - **Switch** (first line acupuncture to guided relaxation and first line guided relaxation to acupuncture) | Pain impact score (PEG) | Opioid use, PROMIS pain interference, PROMIS sleep disturbance, PROMIS gastrointestinal constipation, Generalized Anxiety Disorder-7 (GAD-7), Patient Health Questionnaire-9 (PHQ-9), Patient’s Global Impression of Change, Tobacco, Alocohol, Prescription medication, and other Substance use (TAPS) Tool, Pain Catastrophizing Scale (PCS) | Implementation |
| Drake CL et al., 2022(13) | Adults with clinically significant insomnia who are at risk for major depressive disorder, but do not currently have a depression diagnosis | Adults | Sleep | 2 | - Digital Cognitive Behavioral Therapy for Insomnia (dCBT-I) - Sleep Education Control (SEC) | Non-responders: <8-point reduction in Insomnia Severity Index (ISI) score  Non-responders in the dCBT-I group are re-randomized | All SEC participants and dCBT-I responders **continue** their assigned intervention  Non-responders are re-randomized to:   - **Switch** to clinician-led CBT-I - **Switch** to SEC | Aim 1: Insomnia Severity Index (ISI)  Aim 2: Diagnosis of major depressive disorder via clinical interview using the structured clinical interview for DSM-5 disorders  Aim 3: Pre-Sleep Arousal Scale’s cognitive factor (PSASC) to measure nocturnal rumination | Secondary outcomes were not explicitly defined | Surveys include the QIDS, PSASC, other clinical symptom measures; interviews include the SCID-5 module, new involvement in psychotherapy and/or pharmacotherapy |
| Edelman EJ et al., 2021(14) | Persons with HIV and tobacco use disorder | Adults | Substance use | 2 | - Nicotine replacement therapy (NRT) - NRT + contingency management (CM) | Responders are those whose exhaled carbon monoxide (eCO)-confirmed smoking abstinence at week 12  Non-responders in both groups are re-randomized to second-stage interventions | NRT and NRT+CM responders **continue** their stage 1 intervention in stage 2.  Non-responders in the NRT only group are re-randomized to:   - NRT **augmented** with CM - **Switch** to Varenicline   Non-responders in the NRT+CM group are re-randomized to:   - NRT + CM^+^ (**augmented** to enhanced CM) - **Switch** to Varenicline + CM | Smoking abstinence based on a negative response to “Have you smoked even a puff in the last 7 days?” confirmed with eCO ≤ 6 ppm | CD4 cell count, HIV viral load suppression, and the Veterans Aging Cohort Study (VACS) index 2.0 score | Implementation (feasibility, cost, and future implementation) |
| Eldridge-Smith ED et al., 2022(15) | Individuals with comorbid obstructive sleep apnea and insomnia | Adults | Sleep | 2 | - Positive airway pressure therapy (PAP) + sleep hygiene education - PAP + online cognitive behavioral insomnia therapy (OCBT-I) | Remission criteria is defined as an Insomnia Severity Index score <10    Non-remitters in the stage 1 PAP + OCBT-I group are re-randomized in stage 2 | Stage 1 PAP + hygiene remitters have no additional treatment, but continue to complete all study assessments  Stage 1 PAP + hygiene non-remitters **continue** PAP + hygiene  Stage 1 PAP + OCBT-I remitters have no additional treatment but continue to complete all study assessments  Stage 1 PAP + OCBT-I non-remitters are re-randomized to:   - **Continue** PAP + OCBT - **Switch** to PAP + therapist-delivered CBT-I (TCBT-I) | Remission of insomnia (Insomnia Severity Index (ISI; ISI < 8)) | Subjective and objective sleep data, including sleep time, sleep efficiency, fatigue ratings, PAP adherence, sleepiness ratings, sleep/wake functioning ratings, and objective daytime alertness | Global outcome measures (total wake time, general sleep quality (Pittsburgh Sleep Quality Index – PSQI), Overall daytime functioning (Functional Outcomes of Sleep Questionnaire), safety, acceptability |
| Fernandez ME et al., 2020(16) | Adult tobacco users | Adults | Substance use | 3 | - AAC-Opt In (AAC-In): EHR-based point of care reminder that allows medical staff to choose when to perform Advise and Connect - AAC-Opt Out (AAC-Out): EHR-based point of care that requires clinic staff to Advise and Connect tobacco users to the UTQL or to opt out. | Responders are patients who enroll in the Quitline  Non-responders are patients who do not enroll in the Quitline | First stage non-responders are re-randomized to **switch** to either:   - Text messaging (TM) - Continued AAC (CO)   Non-responders to second-stage TM are randomized to either:   - **Continue** text message (TM-Cont) - **Augment** text message with MAPS (TM+MAPS): continued text messages with two brief phone calls with patient navigators/health educators trained in MAPS counseling | Reach (proportion of tobacco users who enroll in UTQL treatment), impact (the product of reach and efficacy where efficacy is patient abstinence status at 12 months following patient’s initial encounter among those enrolled in the UTQL) | NA | Implementation |
| Flynn D et al., 2018(17) | Active duty service members with chronic pain | Adults | Pain | 2 | - Standard rehabilitative care (SRC) - Complementary and integrative health therapies (CIH) | Non-responders are those who experienced a Pain Impact Score improvement of <3 points  Non-responders are re-randomized in stage 2 | Responders **continue** their stage 1 intervention in stage 2.  Non-responders are re-randomized to:   - **Augment** their stage 1 treatment with the other treatment - **Switch** to the other treatment | Pain impact score | PROMIS measures (global health, depressive symptoms, anxiety, emotional distress (anger), sleep disturbance, and fatigue), functional capacity tests (Modified Naughton treadmill test, floor-waist lift test, waist-shoulder lift test, 40-ft carry test), and force readiness (Military Readiness Category status) | Pain and function surveys, biological measures |
| Fox CK et al., 2024(18) | Adolescents with severe obesity | Adolescents | Overweight and obesity | 2 | - Lifestyle therapy (LST) for 12 weeks with added phentermine (Phen) for an additional 12 weeks if BMI did not decrease by ≥5% - LST for 24 weeks with added phentermine (Phen) for an additional 12 weeks if BMI did not decrease by ≥5% | Those with the added 12 weeks of Phen who did not decrease their BMI by ≥5% were re-randomized in stage 2 | Those in the LST x 12 weeks group who decreased their BMI by ≥5% at 12 weeks **continued** LST for an additional 36 weeks.  Those in the LST x 12 group who did not decrease their BMI by ≥5% at 12 weeks added Phen for an additional 12 weeks. After the Phen x 12 weeks, those who decreased their BMI by ≥5% **continue** Phen + LST for 24 weeks. For those who did not decrease their BMI by ≥5% after Phen x 12 weeks, they were re-randomized to:   - **Augment** with topiramate (TPX) for 24 weeks (i.e. TPX + Phen + LST x 24 weeks - **Switch** to TPX + Placebo + LST x 24 weeks   Those in the LST x 24 weeks group who decreased their BMI by ≥5% at 12 weeks **continued** LST for an additional 24 weeks.  Those in the LST x 24 group who did not decrease their BMI by ≥5% at 12 weeks added Phen for an additional 12 weeks. After the Phen x 12 weeks, those who decreased their BMI by ≥5% **continue** Phen + LST for 12 weeks. For those who did not decrease their BMI by ≥5% after Phen x 12 weeks, they were re-randomized to:   - **Augment** with topiramate (TPX) for 12 weeks (i.e. TPX + Phen + LST x 12 weeks - **Switch** to TPX + Placebo + LST x 12 weeks | Percent change in BMI from baseline measured 48 weeks after baseline | Changes in body composition, cardiometabolic profile (blood pressure, heart rate, lipids, glucose, hemoglobin A1c, ALT, AST), and quality of life (IWQOL-Kids) between baseline and 48 weeks | Safety, biopsychosocial moderators |
| Fritz JM et al., 2020(19) | Active-duty service members and other TRICARE-eligible beneficiaries seeking care for chronic lower back pain | Adults | Pain | 2 | - Physical therapy (PT) - Move 2 health (M2H) | An improvement from a baseline of ≥7 T score points on the pain interference computer-adapted test (PI-CAT) is required to be considered a responder  Treatment non-responders are re-randomized in stage 2 | Treatment responders **continue** up to 2 more sessions of their stage 1 treatment for step-down care.  Treatment non-responders are re-randomized to:   - **Augment** their stage 1 treatment with the other treatment - **Switch** to mindfulness-oriented recovery enhancement (MORE) | Patient-Reported Outcomes Measurement Information System (PROMIS) pain interference computer-adapted test (PI-CAT) | PROMIS physical function CAT (PF-CAT), PROMIS CAT assessments of health domains relevant to chronic lower back pain, including sleep disturbance, depression, and anxiety, Patient Acceptable Symptom State (PASS), European Quality of Life 5-Dimension Instrument (EQ-5D), Defense and Veterans Pain Rating Scale (DVPRS), Pain Intensity, Enjoyment of Life, and General Activity Three-Item Scale (PEG-3), Perceived readiness for duty | Cost-effectiveness, feasibility |
| Fu SS et al., 2017(20) | Current daily smokers with interest in quitting | Adults | Substance use | 2 | - Tobacco Longitudinal Care (TLC): Telephone coaching for 4 weeks - TLC for 8 weeks | Non-responders: any smoking, even just a puff, during the 7 days preceding the data of assessment  Responder: No smoking during the 7 days preceding the date of assessment | Non-responders are re-randomized to either:   - **Continue** TLC - **Augment** TLC+MTM (Medication Therapy Management)   Responders are re-randomized to either:   - **Continue** TLC - **Step down** to TLC-Quarterly: Less intensive TLC | 6-month prolonged abstinence measured 18 months after the beginning of TLC | 30- and 7- day point prevalent abstinence, number of quit attempts, satisfaction with study treatment, self-assessment of cancer risk, cigarettes per day | Other measures include: Physical health, mental health and alcohol use, regulatory focus, smoking history, social support for cessation, smoking cessation-related beliefs and stress, treatment utilization and satisfaction, risk-perception for lung cancer, lung cancer screening outcomes |
| Germeroth LJ et al., 2019(21) | Pregnant women (≤16 weeks pregnant) with overweight/obesity (BMI ≥ 25 kg/m2). | Women of reproductive age | Overweight and obesity and pregnancy | 2 | - Prenatal lifestyle intervention (HABITpreg) - Treatment as usual (TAUpreg) | This is a non-restricted SMART, meaning the second randomization is not influenced by participants meeting specified criteria, and all women are re-randomized prior to delivery | All participants are re-randomized to **switch** to:   - Postpartum lifestyle intervention (HABITpost) - Treatment as usual (TAUpost) | Weight at 12 months postpartum | Cardiometabolic health indicators and psychosocial well-being (insulin resistance, lipids, inflammation markers), depressive symptoms (CES-D, EPDS), perceived stress (PSS scale), sleep quality (PSQI), dietary intake (Nutrition Data System for Research), physical activity) | NA |
| Hassett AL et al., 2023(22) | Adults with chronic lower back pain | Adults | Pain | 2 | - Mindfulness-Based Stress Reduction (MBSR) - Physical Therapy and Exercise (PT) - Acupressure mHealth self-management - Duloxetine | Non-responders: no or minimal improvement in pain (PGIC >=2)  Non-responders to stage 1 interventions are re-randomized in stage 2 | Non-responders are re-randomized to **switch** to one of the first-stage interventions they did not initially receive.  Responders do not receive additional intervention | PROMIS Pain interference short form score at T3 taken at the conclusion of Treatment 1 | NA | Predictive endpoints for phenotyping, mechanistic endpoints for phenotyping, causal analysis endpoints |
| Hibbard JC et al., 2018(23) | Individuals with burn injuries resulting in hypertrophic burn scars | Not specified | Dermatopathy | 3 | - CO2 laser + non-surgical medical therapy (CO2) - Pulse-dye laser + non-surgical medical therapy (PDL) - Non-surgical medical therapy alone (MED) | All participants are re-randomized, independent of on-going response | Stage 2 and 3 interventions are the same as those in Stage 1.  Allowable treatment sequences (including stage 1) are to **continue** or **switch** as follows:   - CO2-CO2-MED - CO2-MED-CO2 - CO2-MED-PDL - CO2-PDL-MED - MED-CO2-CO2 - MED-CO2-PDL - MED-PDL-CO2 - MED-PDL-PDL - PDL-CO2-MED - PDL-MED-CO2 - PDL-MED-PDL - PDL-PDL-MED | Vancouver Scar Score | Quality of life, alternative scar scores | NA |
| Inwani I et al., 2017(24) | Adolescent women and girls with HIV | Adolescents and young adults | HIV | 2 | - Standard referral to receive HIV care and treatment services - Standard referral + SMS reminder to seek care | Non-responders: Not enrolled in care within 2 weeks of the initial intervention  Non-responders to stage 1 interventions are re-randomized in stage 2 | Responders receive no further interventions  Non-responders are randomized to **switch** to either:   - SMS reminder to seek care - Unconditional cash transfer to overcome structural barriers to care | The primary end point for comparisons of recruitment and testing strategies is newly diagnosed HIV infection. | NA | Cost-effectiveness, linkage to care, retention and adherence, HIV prevention-related endpoints, linkage to care |
| Jain S et al., 2023(25) | Children with Multisystem Inflammatory Syndrome (MIS-C) | Children | Inflammatory disorder | 2 | - Infliximab - Steroids - Anakinra | Non-responders meet at least one of the following: fever, persistent or worsening inflammation, or organ damage | Responders receive no further intervention  Non-responders are randomized to **switch** to one of the two treatments not previously assigned | Rate of randomization to secondary treatment | NA | Safety |
| Johnson JE et al., 2018(26) | Outpatient prenatal clinics serving women on public assistance | Adults | Mental and behavioral health | 2 | - Continue Enhanced implementation as usual (EIAU) - EIAU + Low-Intensity Coaching and Feedback (LICF) | Non-responders to EIAU+LICF: Either no ROSE intervention in 3 months and none planned or less than 75% fidelity to ROSE core elements | Those randomized to continue EIAU in stage 1 receive no further intervention  Non-responders to EIAU+LICF are re-randomized to either:   - **Continue** EIAU + LICF - **Augment** to EIAU + LICF + High intensity coaching and feedback (HICF)   Responders to EIAU+LICF **continue** with EIAU+LICF | Percent sustainment of core program elements at each timepoint | PPD rates over time at each clinic | Reach, cost-effectiveness, implementation |
| Kilbourne AM et al., 2014(27) | Community-based mental health or primary care clinics in Michigan or Colorado | Medical centers, practices, or providers | Mental and behavioral health | 2 | - (REP) with external facilitation (REP+EF) - REP with internal and external facilitation (REP+EF/IF) | Non-responders were sites with <50% of patients receiving >=3 EBP sessions | Non-responders to REP+EF are randomized to either:   - **Continue** REP+EF - **Augment** to REP+EF/IF   Non-responders to REP+EF/IF **continue** REP+EF/IF  Responders do not receive additional intervention | Mental health-related quality of life | Receipt of LG sessions, mood symptoms, implementation costs, and organizational change | Implementation, cost-effectiveness, organizational and provider factors |
| Kopelowicz A et al., 2023(28) | Mexican American individuals with type II diabetes mellitus | Adults | Diabetes | 2 | - Tomando control (TC) intervention led either by a community health worker or promotore (TC-promotore) - TC led by a registered nurse (TC-Nurse) | Non-responders: 50% gain over baseline on the Summary of Diabetes Self-Care Activities (SDSCA) | Non-responders are randomized to either:   - First stage intervention **augmented** by family-focused TC - **Switch** to Multifamily Group (MFG)   Responders **continue** their first-stage intervention | Difference in the SDSCA between the groups at 3, 6, and 12 months after baseline | Diabetes self-efficacy, diabetes knowledge, degree of family support, collaborative goal setting, treatment session attendance, glycemic control | Attendance, vitals |
| Kor PP et al., 2023(29) | Community-dwelling full-time caregivers (FC) of persons with dementia (PWD) | Caregivers | Neurological disease or injury | 2 | - Behavioral activation (BA) - a psychological intervention that encourages engagement in meaningful, healthful, and enjoyable activities to improve mood - Mindfulness practice (MP) - a psychosocial intervention focused on increasing mindfulness through exercises like meditation, mindful breathing, and body scanning | Non-responders are those with a reduction in PHQ-9 scores of less than 50%  Non-responders are re-randomized in the second stage | Responders proceed to monthly booster sessions.  Non-responders are re-randomized to:   - **Augment** their stage 1 intervention with an additional self-efficacy enhancing strategy - **Switch** to the other intervention | Depressive symptoms (PHQ-9) | Perceived caregiving stress, positive aspects of caregiving (PAC), sleep quality, and health-related quality of life (HRQoL) | Engagement in meaningful activities and level of mindfulness (patient logbooks and Five Facet Mindfulness Questionnaire) |
| Levy R et al., 2019(30) | Adults with major depressive disorder and/or PTSD in low- and middle-income countries | Adults | Mental and behavioral health | 2 | - Interpersonal psychotherapy (IPT) - Fluoxetine (F) | Non-responders are those who continue to meet MINI criteria for MDD and/or PTSD diagnosis | Non-responders are randomized to either:   - **Switch** to the other first-stage treatment - Combination of first-stage treatments (**augment**) | Major Depressive Disorder (MDD) and Posttraumatic Stress Disorder (PTSD) using MINI-MDD and MINI-PTSSD modules | Accessibility; affordability; domestic violence, alcohol, drugs, and trauma; physical health; emotional reactivity; social support | Economic, implementation |
| Li X et al., 2021(31) | Individuals with at least one episode of schizophrenia, schizophreniform, or schizoaffective disorder | Adolescents and adults | Mental and behavioral health | 2 | - Olanzapine - Risperidone - Amisulpride - Aripiprazole - Perphenazine (a first-generation antipsychotic) | Response is defined as a ≥40% reduction in Positive and Negative Syndrome Scale (PANSS) total score  Stage 1 non-responders (or those with intolerable side effects) are re-randomized to the second-stage intervention  Stage 2 non-responders are re-randomized to the third-stage intervention | Stage 1 responders **continue** their original treatment    Phase 1 non-responders are re-randomized to **switch** to one of the following three treatments (no one is assigned to the same drug as phase 1):   - Olanzapine - Amisulpride - Clozapine (which is often reserved for treatment-resistant schizophrenia)   Phase 2 clozapine non-responders are re-randomized to:   - **Continue** clozapine (clozapine extended treatment) - **Switch** to modified electroconvulsive therapy (MECT) add-on therapy   Phase 2 non-clozapine non-responders are re-randomized to:   - **Switch** to clozapine - **Switch** to another SGA not previously used in phase 1 or 2 | Treatment phase: treatment efficacy rate, which is defined as a 40% reduction or more of the total score in the Positive and Negative Syndrome Scale (PANSS)  Naturalistic follow-up phase: time to all-cause treatment failure, marked by its discontinuation, and all-cause discontinuation is defined as discontinuing for any reasons (including poor efficacy, intolerance of adverse reactions, poor compliance et al.) | Secondary outcomes were not specified | Clinician and functional assessments, neurocognitive assessments, neuroimaging, pharmacoeconomic and cost-effectiveness assessments |
| Lion KC et al., 2023(32) | Primary care providers in Washington state and adult patients or parents of pediatric patients who use a language other than English. | Medical centers, practices, or providers | Health care delivery | 2 | - Web-based education modules - Mobile video interpreting access | Providers with interpreter use in the top tertile (within strategy) will remain with the original strategy; those in the bottom two tertiles will be re-randomized | Providers in the top tertile remain in their original intervention  Providers in the bottom two tertiles were randomized to either:   - **Continue** with their original intervention - Receive both interventions (combination, **augment)** | Interpreter use | Patient comprehension, determined by comparing patient-reported to provider-documented visit diagnosis | Cost-effectiveness |
| Liu H et al., 2021(33) | Individuals with psychotic symptoms or major depressive disorder | Adults | Mental and behavioral health | 2 | - Monthly Brief Contact Interventions (BCIs) - Weekly BCIs | Participants initially randomized to monthly BICs will be rerandomized if their suicide risk increases, while participants initially randomized to weekly BCIs will be rerandomized if their suicide risk decreases. Suicide risk is defined as the probability of an individual’s death by suicide over a given time interval reflected by the intensity and frequency of suicide ideation, suicide plan, suicide preparation and suicide attempts. Suicide risk will be evaluated by the Beck Suicide Ideation Scale-Chinese Version (BSI-CV) and the suicidality module of the Mini-International Neuropsychiatric Interview (M.I.N.I-Suicidality) | For those initially assigned to monthly BCIs:   - If suicide risk increases, they are randomized to **augment** to either:   - Weekly BCIs   - Biweekly BCIs - If suicide risk is decreases or remains the same, **continue** with monthly BCIs   For those initially assigned to weekly BCIs   - If suicide risk increases or remains the same, **continue** weekly BCIs - If suicide risk decreases, they are re-randomized to **augment** to either: - Monthly BCIs - Biweekly BCIs | Trajectories of suicide risk (suicide ideation and suicidality) from baseline to 3-month and 12-month post-discharge | Trajectories of suicide risk from 3-month to 12-month post-discharge, trajectories of social connectedness and social support from baseline to 3-month and 12-month post-discharge | Implementation and service outcomes |
| Markland AD et al., 2023(34) | Women veterans with urinary incontinence (UI) | Adults | Urological disorder | 2 | - MyHealtheBladder: Interactive mHealth treatment for urinary incontinence (UI) - VA Video Connect (VVC): single virtual visit with a UI clinician | International Consultation on Incontinence Questionnaire – Urinary Incontinence Short From (ICIQ-UI) < 2.52 at 8 weeks | Responders **continue** their initial treatment  Non-responders are re-randomized to either:   - **Continue** first-stage treatment - **Switch** or **augment** to VA Video Connect Booster Visit | International Consultation on Incontinence Questionnaire-Urinary Incontinence Short Form (ICIQ-UI SF) | Other lower urinary tract symptoms, impact on quality of life, adherence to the pelvic floor muscle exercises, retention rates for program completion, satisfaction with treatment, perceptions of improvement, costs of incontinence care, miles saved, usability of the mHealth app, and adaptive behaviors related to UI | Cost, usability, implementation feasibility (miles saved) |
| Micheletti RG et al., 2020(35) | Adults with chronic skin-limited forms of vasculitis | Adults | Inflammatory disorder | 2 | - Azathioprine - Colchicine - Dapsone | Non-response: Patients who subsequently discontinued the study drug during the 6 months of the first stage of the study or the follow-up period (month 6 to month 12) because of any one of the following endpoints per protocol-defined definitions: a lack of response or failure, a relapse (month 6 to month 12), or a side effect necessitating drug discontinuation 2. Patients with a history of significant intolerance, allergy, or serious adverse events to one of the study medications, deficit in glucose-6-phosphate dehydrogenase (G6PD) or thiopurine methyltransferase (TPMT) or history of hemolytic anemia, or who failed to respond (according to the study definition) to one of the study drugs prior to screening | Responders **continue** initial treatment  Non-responders are re-randomized to **switch** to one of the other two drugs not assigned in the first stage | Proportion of participants with a response to therapy at month 6 of the pooled study Stages 1 and 2 | Proportion of patients with complete response to therapy (no new lesions within the preceding 3 months) at months 3, 6, and 12; proportion of patients with significant response to therapy (three or fewer minimally symptomatic lesions per flare, with no more than one flare per month in the preceding 3 months) at months 3, 6, and 12; time to achieve complete or significant response; time to vasculitis flare for patients who achieved a complete or significant response before month 6 but subsequently relapsed; frequency of vasculitis flares/new lesions compared to baseline; physicians' global assessments of response; patients' global assessments of response; prednisone use during the study period; Skindex29 score at months 1, 3, 6, 9, and 12 (Captures skin-related pain, pruritus, discomfort); patient-reported outcomes/disease burden at months 1, 3, 6, 9, and 12; response according to patient and disease characteristics; grading of standardized photographs by blinded investigators" | NA |
| Nelson B et al., 2018(36) | Young people at ultra-high risk (UHR) of psychosis | Youth and young adults | Mental and behavioral health | 2 | Open label support and problem solving (SPS) will be provided to all participants.  Responders continue SPS monthly until month 12  Non-responders to SPS will be randomized to either:   - Cognitive behavior case management (CBCM) - SPS | Response is defined as:  The Global Rating Scale or the Frequency score need to be less than 3 for all of the four positive symptoms (unusual thought content, non-bizarre ideas, perceptual abnormalities and disorganized speech) and there is an improvement of at least 5 points in Social and Occupational Functioning Assessment Scale (SOFAS) compared with baseline or the SOFAS score is at least 70  Non-responders are re-randomized | Responders will **continue** or **switch** to SPS monthly until month 12  CBCM non-responders are re-randomized to:   - **Augment** with SSRI - **Augment** with Placebo   SPS non-responders are re-randomized to:   - **Switch** to CBCM + SSRI - **Switch** to CBCM + placebo | 6-month Global Functioning Social and Role Scale | Secondary outcomes were not distinguished | Safety, psychopathology, functioning and quality of life, biological, neurocognition, neuroimaging, cerebrospinal fluid |
| O'Keefe VM et al., 2019(37) | American Indian youth aged 10-24 years old who reside on or near the Fort Apache Indian Reservation | Youth and young adults | Mental and behavioral health | 2 | - New Hope + Optimized Case Management (OCM): A two-to-four-hour session with an Apache Community Mental Health Specialist focused on reducing immediate suicide risk - OCM alone: Monthly case management visits by independent evaluators who connect youth to behavioral health or community services and administer suicide risk assessments. | All participants are re-randomized regardless of first-stage intervention or response to first-stage intervention | Everyone was re-randomized to:   - **Switch** to Elders’ Resilience + OCM: A two-to-four hour session with Apache Elders aimed to promote resilience - **Continue** or **step down** to OCM: same as the first stage | Suicidal ideation (SIQ/SIQ-JR) and resilience (modified Resiliency Scales for Children and Adolescents (RSEA)) | Depressive and anxiety symptoms, impulsivity, self-efficacy and communal mastery, importance of following AI values and cultural practices, self-esteem, hope, substance use, and a subset of local WMAT generated items relating to changes observed among youth who have previously received New Hope or Elders’ Resilience interventions | NA |
| Osilla KC et al., 2023(38) | Spouses and partners of active-duty service members who are concerned about their partner’s drinking | Caregivers or partners | Substance use | 2 | - Partners Connect: a four-session web-based intervention that utilizes CRAFT principles that was designed to help concerned partners (CPs) increase self-care and improve their communication skills regarding their service members (SMs) drinking. - Gottman Resources: The Gottman Institute website was developed based on decades of observational research with couples, showing how negative communication and emotional flooding are highly predictive of poorer family and individual well-being | Non-responders are those where the SM does not access PNF or click “done” | Partners Connect non-responders are re-randomized to:   - **Switch** to CRAFT Workbook: The CRAFT workbook helps us evaluate what combination of self-directed materials could help dyads who may not otherwise seek treatment - **Switch** to Phone-CRAFT: The phone-based CRAFT intervention consists of six individual sessions with a CRAFT-trained clinician reviewing the CRAFT workbook content.   Gottman Resources non-responders are re-randomized to:   - **Switch** to Partners Connect - **Switch** to CRAFT Workbook | CP primary outcome: Drinking (number of drinks per week in the past month)  SP primary outcome: help-seeking through response to the SMART intervention (clicking “done” at the end of the PNF program) | CP outcomes: mental health symptoms (anxiety/depression), communication via the Communication Patterns Questionnaire, social support from the Medical Outcomes Study Social Support Scale  SM outcomes: additional help seeking behaviors (preparatory behaviors, intentions, or actual care initiation) | CP communication, CP drinking, SM perceived drinking norms |
| Peter SC et al., 2023(39) | Individuals seeking treatment for opioid use disorder | Adults | Substance use | 2 | - Brief Motivational Interviewing + Substance Free Reinforcement + Mindfulness Sessions (BSM): weekly discussion centered around increasing engagement in substance-free activities that are enjoyable and/or consistent with the individual’s goals or values - Contingency Management (CM): a behavioral treatment where prizes or vouchers are provided to patients based upon proof of a desired behavior such as abstinence or adherence to a medication | A participant is considered non-adherent if they meet either of the following criteria:   - Missed a physician visit for buprenorphine-naloxone treatment. - Buprenorphine is absent in their urine toxicology screen.   Non-adherent participants are re-randomized in stage 2 | Participants who are adherent will **continue** their initial treatment for up to 6 months  Non-adherent participants are re-randomized to:   - **Augment** their original intervention with the other treatment (combination of the two) for 6 months - **Switch** to the other treatment for 6 months | Medication adherence | Secondary outcomes were not distinguished | Behavioral economic-related variables, mental health measures |
| Peterson BS et al., 2021(40) | Children and adolescents aged 8-17 years who have been diagnosed with an anxiety disorder | Children and adolescents | Mental and behavioral health | 2 | - Fluoxetine: SSRI medication targeting the biological mechanisms of anxiety by increasing serotonin levels - Cognitive behavioral therapy (CBT): focuses on exposure therapy, coping skills development, and cognitive restructuring | Criteria for remission includes: (a) a youth screen for child anxiety related disorders (SCARED-41) score less than diagnostic threshold for any single anxiety disorder, together with (b) a total youth SCARED-41 score < 10, and (c) a score of ≤8 on the CAIS (Child Anxiety Impact Scale).  Non-remitters were re-randomized in the second stage | Remitters **continue** their stage 1 treatment  Non-remitters are re-randomized to:   - **Continue** their stage 1 treatment for 12 weeks - **Augment** their stage 1 treatment with the other treatment for 12 weeks | Self-report score on the 41-item youth SCARED (Screen for Child Anxiety Related Disorders) | Parent ratings of anxiety symptom severity on the Youth SCAR ED-41, and parent and youth ratings on the CAIS (Child Anxiety Impact Scale) | Patient-centered outcomes; social, family, and school functioning; mental health and emotional regulation; sleep and side effects; treatment experience and acceptability |
| Pfammatter AF et al., 2019(41) | Adults with overweight and obesity | Adults | Overweight and obesity | 2 | - mHealth SMART app (App): app used to record dietary intake and to monitor graphical feedback that displays their dietary intake, physical activity, and weight relative to daily and long-term goals - mHealth SMART app + coaching (App + C): diet and activity coaching are provided together with the app. Coaching calls last approximately 10-15 min and include feedback and problem solving delivered within a motivational interviewing (MI) framework | If the average weekly weight loss is found to be <0.5 pounds at the 2, 4, or 8 week time point, the participant will be classified as a non-responder  Non-responders are re-randomized in the second stage | Responders **continue** their stage 1 intervention  Non-responders are re-randomized to:   - Modestly step up (**augment**) their initial treatment with texts (+ T) - Vigorously step up (**augment**) their initial treatment with texts and coaching for the stage 1 App group (App + T + C) and with texts and meal replacement for the stage 1 app + C group (App + T + C + M) | Change in weight (kilograms) from baseline to 6 months | Secondary outcomes were not distinguished | Cost-effectiveness, self-efficacy, autonomous motivation |
| Quanbeck A et al., 2020(42) | Primary care clinics from two health systems in the Midwestern US and their providers. Individuals with 3+ consecutive months of opioid prescriptions will be used to assess outcomes | Medical centers, practices, or providers | Substance use | 2 | - EM/AF: Engagement meetings and audit and feedback which included educational information and engagement through in-person and webinar delivery and performance feedback on opioid prescribing practices - EM/AF + PF: Engagement meetings and audit feedback plus practice facilitation defined as EM/AF + an external facilitator focused improving clinic-level opioid prescribing workflows | All clinics are re-randomized regardless of first-stage intervention or response to first-stage intervention | First stage EM/AF clinics were re-randomized to either:   - **Continue** EM/AF only - **Augment** EM/AF + PPC   First stage EM/AF + PF clinics were re-randomized to either   - **Continue** EM/AF + PF - **Augment** EM/AF + PF + PPC   PPC is prescriber peer consulting, defined as quarterly access to physician or pharmacist experts experienced in opioid management | Average of morphine-milligram equivalent (MME) dose per day per opioid patient, calculated over a 3-month period | NA | Cost estimates of the 4 sequences, implementation outcomes, adoption and reach metrics |
| Rabin BA et al., 2023(43) | Adults in underserved communities who receive care at federally qualified health centers | Adults | Vaccine uptake | 2 | - Standard of care (SOC; control group): patients receive routine healthcare services at their Federally Qualified Health Center (FQHC). Includes routine immunization efforts, annual physical exams, and preventive screenings - mHealth Outreach (intervention group – lower intensity): patients receive mobile health (mHealth) messages (text messages and automated voice calls). Messages are culturally tailored and linguistically appropriate | Non-response is defined as not being up to data on COVID-19 vaccination OR having at least one outstanding preventive service need  Non-responders are re-randomized in stage 2 | Responders **continue** their stage 1 intervention  Non-responders in the mHealth group are re-randomized to:   - **Continue** mHealth outreach - **Augment** mHealth outreach with care coordination: the deliberate organization of patient care activities between two or more participants involved in a patient’s care to facilitate the appropriate delivery of health care services   Non-responders in the SOC group are re-randomized to:   - **Switch** to mHealth outreach - **Switch** to mHealth outreach + care coordination | COVID-19 vaccine completion | Increase in preventive service engagement, increase in vaccine confidence | Implementation, reach, adoption, and maintenance |
| Sabri B et al., 2021(44) | Immigrant women who have experienced intimate partner violence (IPV) | Adults | Intimate partner violence | 2 | - myPlan app: a safety decision and planning tool delivered through a free and secure web-based app. - Usual care control | Non-responders: women who report both no improvement in safety (i.e., significant reduction in severity and frequency of IPV) and empowerment scores (i.e., significant improvement in empowerment) from baseline to 3 months  Non-responders are re-randomized to second stage interventions | Responders **continue** their original intervention.  Non-responders in both groups are re-randomized to:   - **Switch** to safety and empowerment strategies delivered via text - **Switch** to combination of text and phone calls with trained advocates | Severity and frequency of IPV (Revised Conflict Tactics Scale (CTS2)), empowerment (Personal Progress Scale-Revised (PPS-R), MOVERS scale) | Depression (PHQ-9), Post-traumatic stress disorder (PTSD) | NA |
| Schlechter CR et al., 2023(45) | Adult Medicaid patients with obesity | Adults | Overweight and obesity | 2 | - Single Text Message (TM1) - Multiple Text Message (TM+) | Non-responders: No enrollment in evidence-based interventions, including improving diet and nutrition, increasing physical activity, and behavioral strategies such as goal setting and self-monitoring | Non-responders to TM1 are randomized to either:   - **Step down** to no additional text messages - **Switch** to Motivation and Problem Solving Counselling (MAPS)   Non-responders to TM+ are randomized to either   - **Continue** TM+ - **Augment** TM+ and MAPS   Responders receive no further intervention | NA | NA | Feasibility, acceptability |
| Skolasky RL et al., 2020(46) | Individuals with nonspecific lower back pain | Adults | Pain | 2 | - Physical Therapy (PT) - Cognitive Behavioral Therapy (CBT) | Non-responders: Not experiencing at least 50% reduction in pain-related disability | Non-responders to PT are randomized to either:   - **Switch** to CBT - **Switch** to Mindfulness-Oriented Recovery Enhancement (MORE)   Non-responders to CBT are randomized to either:   - **Switch** to PT - **Switch** to MORE   Responders **continue** first-stage therapy | Disability (Oswestry Disability Index), pain intensity (Numerical Pain Rating Scale) | General health (PROMIS-29 SF), long term opioid use, health care utilization | Implementation, physical adverse effects, risk for poor outcomes |
| Smith SK et al., 2021(47) | Hematopoietic stem cell transplantation (HCT) survivors reporting posttraumatic stress disorder (PTSD) | Adults | Mental and behavioral health | 2 | - Cancer Distress Coach (CaDC) App: a mobile mHealth intervention based on the PTSD Coach app. Includes educational content, self-guided coping strategies, symptom tracking, and access to resources - Usual care control: standard mental health services available to all cancer patients at their institution | Non-responders: <5-point reduction in PTSD symptom severity score  Non-responders are re-randomized to second stage interventions | Responders **continue** their initial intervention  Non-responders in the CaDC group are re-randomized to:   - **Augment** CaDC with mCoaching: virtual clinician support - **Switch** to mHealth cognitive behavioral therapy (mCBT): HCT-specific telephone administered CBT protocol   Non-responders in the usual care control group are re-randomized to:   - **Switch** to CaDC + mCoaching - **Switch** to mCBT | PTSD Checklist for DSM5 (PCL5) with items modified to key on cancer-related symptoms, as has been validated in the HCT population | Cancer-related distress, depression, and anxiety (Distress Thermometer v.2018); general perceptions of health (NIH PROMIS Global QOL); depression, anxiety, and pain interference (PROMIS 8-item instruments); how capable one feels in managing symptoms and performing health-related tasks (Self-efficacy for Managing Chronic Disease Scale); post-intervention survey includes questions regarding the participant's study experience (i.e., perceived helpfulness and satisfaction) to inform future improvements to the protocol and/or programs; CaDC clickstream activity (i.e., usage) | Participant satisfaction, adherence |
| Sripada RK et al., 2023(48) | Adults with post-traumatic stress disorder (PTSD) | Adults | Mental and behavioral health | 2 | - Prolonged Exposure for Primary Care (PE-PC): 30-45 minute sessions of in-vivo and imaginal exposure - Clinician Support (CS) PTSD Coach: incorporates evidence-based assessment, psychoeducation, cognitive behavioral therapy and self-management strategies that are customizable to the user | Early responders are participants who experience at least a 15-point symptom reduction or have a PCL-5 score <29  Slow responders are re-randomized to second stage interventions | Early responders to PE-PC **step down** to every other week  Early responders to CS PTSD Coach **step down** to self-managed (SM) PTSD Coach  Slow responders in both groups are re-randomized to:   - **Continue** first line treatment - **Step up** to Full PE: in-vivo exposure, imaginal exposure to the trauma memory, and facilitated emotional processing of the trauma | 3-month score on the Clinician-Administered PTSD Scale for DSM-5 (CAPS-5) | PCL-5, a 20-item self-report measure of PTSD symptoms as defined by the DSM-5 | Process evaluation, fidelity measures, risk assessment, cost-effectiveness |
| Tamura K et al., 2020(49) | African American women at risk for cardiovascular disease | Adults | CVD and CVD risk | 2 | - Tailored-to-place message (TPM) - Standard remote messaging (SRM) | Non-responders: those who do not meet the PA goal of 10,000 steps/day after 3 months to more intensive treatment | Responders **continue** initial intervention  Non-responders to TPM are re-randomized to either:   - **Augment** TPM + face-to-face coaching - **Step up** TPM with greater frequency   Non-responders to SRM are re-randomized to either:   - **Augment** SRM + face-to-face coaching - **Switch** to TPM | NA | NA | Primary and secondary outcomes were not distinguished.  Survey data (health history, access and the behavioral, psychosocial, cultural, and perceived environmental factors that may influence the impact of intervention), neighborhood environment measures, mHealth data (step count, minutes of vigorous activity, sleep time), clinical measures |
| van Heerden A et al., 2023(50) | Adults living with HIV who are eligible for ART and have a detectable viral load and/or are not engaged in care | Adults | HIV | 2 | - Best Clinic Practices (Standard of Care – SOC): Welcome back service (friendly providers, SMS, adherence support) and optimized ART refills - Best Clinic Practices + Lottery Incentives: Conditional lottery incentives, welcome back service, and optimized ART refills | Responders are participants who are virally suppressed and engaged in care at month 6  Non-responders are participants with a detectable viral load or not engaged in care/lost to follow-up (LTFU)  Non-responders are re-randomized to second stage interventions | Responders **continue** their original intervention  Non-responders are re-randomized to:   - **Continue** original intervention - **Switch** to smart lockers (Pele boxes): decentralized ART refills and monitoring and adherence support - **Switch** to home-delivery and monitoring of ART: Home ART refills and monitoring and adherence support | Viral suppression at 18 months | Retention in care, time to ART initiation | Qualitative preferences of clients and providers, cost-effectiveness and economic measures |
| Velloza J et al., 2022(51) | Sexually active women between ages 18 and 25 who are newly initiating PrEP and have regular access to a mobile phone | Young adults | HIV | 2 | - Two-way SMS: Weekly SMS messages with follow-up if participants respond that they are not feeling well or fail to respond - WhatsApp support group: Virtual peer support groups moderated by a staff facilitator and “PrEP Ambassador” | Responders: Dried blood spot (DBS) tenofovir diphosphate (TFV-DP) levels >= 500 fmol/punch at month 2  Non-responders: TFV-DP levels <500 fmol/punch or have missed PrEP refills | Responders **continue** first-stage treatment  Non-responders are re-randomized to either:   - First-stage intervention **augmented** by quarterly drug-level feedback counseling where participants get personalized adherence feedback - First-stage intervention **augmented** by intensive problem-focused counseling sessions during which adherence barriers and facilitators are discussed | PrEP adherence measured as DBS TFV-DP drug concentration >=700 fmol/punch at nine monhts | PrEP adherence, analyzed as continuous TFV-DP levels as well as in a five-category variable (below the limit of quantification [BLQ], BLQ–349 fmol/punch, 350–699 fmol/punch, 700–1249 fmol/punch, and >=1250 fmol/punch), shorter-term impact of interventions on TFV-DP levels at two months | Acceptability and feasibility of interventions, adherence, and discontinuation |
| Walton MA et al., 2023(52) | Adolescents and young adults ages 14-20 presenting to EDs who screen positive for binge drinking and violent behaviors | Adolescents and young adults | Substance use | 2 | - TM: Tailored text messages automatically sent 2/day for 4 weeks - HC: Health coach telephone call 1/week for 4 weeks | Responder: Not reporting any binge drinking or aggression in weeks 3 and 4  Non-responders: Reporting binge drinking or aggression in weeks 3 or 4, those missing survey data in weeks 3 or 4 | Responders were re-randomized to either **continue** condition or **stepped down**:   - Responders to TM or HC who are randomized to the stepped down condition receive a brochure   Non-responders were re-randomized to either **continue** condition or **stepped-up** condition   - Non-responders to TM who are randomized to the **stepped-up** condition receive HC calls - Non-responders to HC who are randomized to the **stepped-up** condition receive the intensified HC condition (HCs send supportive text messages between calls) | Alcohol consumption and violence, both using 30-day timeline follow back (TLFB) interview | Alcohol-related consequences (modified Brief Young Adult Alcohol Consequences Questionnaire (B-YAACQ)), violence consequences | Other drug use (ASSIST-Lite) and victimization (past 30-day TLFB) |
| Wan Y et al., 2023(53) | Community-dwelling people living with dementia and their caregivers | Caregiver-individual dyads | Neurological disease or injury | 2 | - Monthly Online Collaborative Care (WeChat Mini Program-based Intervention: a digital platform designed for dementia care - Monthly Offline Collaborative Care (Traditional Face-to-Face Home Visits): care provided in person via home visits by healthcare providers | Responders are those who experience improvement on patients' quality of life AND reduction of caregivers' burden. An improvement in quality of life for patients with dementia (response) is defined as scoring 10 points higher on the QOL-AD at the checkpoint than at the baseline. A reduction in caregivers’ burden (response) is defined as a decrease in burden severity of at least one level on the Zarit Burden Interview (ZBI) compared with baseline, such as from severe to mild or from moderate to mild.  Responders are re-randomized to stage 2 interventions | Non-responders **step up** their stage 1 intervention to biweekly  Responders are re-randomized to:   - **Continue** their stage 1 intervention monthly - **Step down** their stage 1 intervention to bimonthly | Proportion of patients demonstrating an improvement in quality of life and the proportion of caregivers exhibiting a reduction in caregiver burden (improvement in quality of life (10 points higher on QOL-AD), reduction in caregiver burden (decrease in burden severity of at least one level on ABI compared to baseline)) | Cost-effectiveness of collaborative care, cognitive change of patients, change in activities of daily living of patients with dementia, change in the number of behavioural and psychological symptoms of patients with dementia, change in depressive symptoms of patients with dementia and caregivers, change in anxiety symptoms of caregivers | Feasibility |
| Wilroy J et al., 2023(54) | Individuals with spinal cord injury (SCI) | Adults | Neurological disease or injury | 2 | - Asynchronous Movement-to-Music (M2M) for 3 weeks - Asynchronous M2M for 6 weeks | Non-responders <95% of video watch minutes | Non-responders are randomized to either:   - **Augmentation** of Asynchronous M2M with Individual Behavioral Coaching (IBC) - **Switch** to M2M live   Responders **continue** asynchronous M2M | NA | NA | Feasibility (primary aim), acceptability (secondary aim), exercise adherence, physical activity, physiological outcomes, psychosocial outcomes, app quality and usability |
| Windsor L et al., 2022(55) | Individuals at high risk for COVID-19 infection | Adults | Mental and behavioral health | 2 | - Navigation Services (NS) - Digital Brochure (DB) | Non-responders: Failure to complete COVID-19 testing with 7 days after the first intervention session | NS responders are randomized to either:   - **Continue** NS - **Switch** to Critical Dialogue (CD)   NS non-responders are randomized to either:   - **Continue** NS - **Switch** to Brief Counseling (BC)   DB responders are randomized to either:   - **Continue** DB - **Switch** to BC   DB non-responders are randomized to either:   - **Continue** DB - **Switch** to CD | Completion of at least one COVID-19 test within 1 week of the first intervention session | Adherence to the prevention guidelines in the past month (Multi-ethnic Study of Atherosclerosis (MESA)) | Process (intervention fidelity, quality assurance, and facilitator competence), integrity assessments, predictor and outcome standardized measures to assess social determinants of health, demographics, COVID-19 risk (including comorbidities, exposure to SARS-CoV-2, adherence to prevention guidelines), attitudes about COVID-19 testing, health and social services used, knowledge about COVID-19, attitudes about COVID-19 vaccines and booster shots, mental health, protective factors and barriers to health care, confidence in government, and the healthcare system |
| Yan X et al., 2022(56) | Elite or amateur athletes taking part in regular training | Adults | Nutrition | 3 | - App + relaxed - App + stringent | Non-responders to stage 1 intervention: if randomized to App + relaxed in stage 1, failure to engage with the app at least once; if randomized to App + stringent in stage 1, failure to engage with the app at least twice  Non-responders to stage 2 intervention: if randomized to App + relaxed in stage 1, failure to engage with the app at least twice; if randomized to App + stringent in stage 1, failure to engage with the app at least three times" | Stage 1 responders **continue** their original intervention  Non-responders to App + relaxed in Stage 1 are randomized to either:   - **Continue** App + relaxed - **Augment** App + relaxed + Nutritional Coaching (NC)   Non-responders to App + stringent in Stage 1are randomized to either:   - **Continue** App + stringent - **Augment** App + Stringent + NC   Stage 2 responders **continue** original intervention  Stage 2 non-responders who were assigned App + relaxed in Stage 1 are randomized to either:   - **Continue** App + relaxed - **Augment** App + relaxed + NC   Stage 2 non-responders were who assigned App + stringent in Stage 1 are randomized to either:   - **Continue** App + stringent - **Augment** App + stringent + NC | Mobile application and coach uptake, retention rate at week 5, app engagement, and carbohydrate periodization behavior at week 5 | Self-reported weight, goal, carbohydrate periodization self-efficacy, and belief about consequences | Impact of personality and need for autonomy on application usage, cost-effectiveness, feasibility |
| Zhao SZ et al., 2022(57) | Cigarette smokers aged 18 years or older in Hong Kong who are motivated to quit or reduce smoking | Adults | Substance use | 2 | - PIM: Personalized instant messaging including real-time interactive conversation with smoking cessation conselors - RIM: Regular instant messaging with generic, fixed-schedule messages on smoking cessation | Non-responders were those still smoking at one month follow-up | Responders **continue** stage 1 intervention  Non-responders to PIM were re-randomized to either:   - **Switch** to OCI: Optional combined intervention, where participants can choose from a number of interventions or combinations thereof - **Continued** PIM   Non-responders to RIM were re-randomized to either:   - **Continue** RIM - **Switch** to PIM | Biochemically validated abstinence at 6 months after treatment initiation using exhaled carbon monoxide (<4 ppm) and salivary cotinine (<10 ng/ml) | Validated abstinence at 3 months (end of treatment), self-reported past 7 days abstinence, smoking reduction by at least 50% of baseline consumption, quit attempt (abstinence for ≥24 h), and smoking cessation services use, defined by any use of provided treatments (e.g. counselling, medication, acupuncture) | Patient experience, adherence |
| Zhou G et al., 2020(58) | Individuals living in clusters (villages or several neighboring villages) southeast of Kisumu County, western Kenya | Villages or clusters of villages | Infectious disease | 2 | - LLIN: Long-lasting insecticidal nets - PRO LLIN: Piperonyl butoxide-treated long-lasting insecticidal nets - LLIN + IRS: Long-lasting insecticidal nets combined with indoor residual spraying of Actellic 300CS | Responders are clusters with a statistically significant reduction in malaria incidence and greater than the pre-defined threshold value set by the Ministory of Health of Kenya  Non-responders are clusters that do not meet the responder criteria | Those original randomized to LLIN **continue** LLIN in stage 2  PBO LLIN responders **continue** PBO LLIN in stage 2  Non-responders to PRO LLIN are re-randomized to either:   - **Augment** PBO LLIN + larval source management (LSM) - **Switch** to enhanced method: an intervention determined adaptively based on stage 1 effectiveness analyses   LLIN + IRS responders **continue** LLIN + IRS  Non-responders to LLIN + IRS are re-randomized to either:   - **Augment** to LLIN + IRS + LSM - **Augment** to PRO LLN + IRS | Clinical malaria incidence rate | Malaria vector abundance and transmission intensity | Cost-effectiveness |
| Zullig LL et al., 2021(59) | Cancer patients with cardiovascular risk factors/comorbidities | Adults | Cancer | 2 | - iGuide intervention (self-guided) - Control | Non-responders: PCP clinics with 90% or more of their participating patients do not meet the modified HEDIS quality metrics at the 6-month measurement  Stage 1 non-responders are re-randomized | Those originally randomized to control **continue** control  iGuide responders continue iGuide intervention in stage 2  Non-responders to stage 1 iGuide intervention are re-randomized to either:   - **Continue** iGuide - **Step up** to iGuide 2 (tailored and targeted) | Health Effectiveness Data and Information Set (HEDIS), Proportion of Days Covered (PDC), Patient-centered Communication in Cancer (PCC-Ca-36) | Self-reports of medication adherence, cancer care-related financial toxicity, out-of-pocket expenses by category of spending, engagement, and care coordination | Cost, patient engagement, care coordination |
| Primary analysis papers |  |  |  |  |  |  |  |  |  |  |
| Butzer JF et al., 2023(60) | Persons with traumatic spinal cord injury | Adults | Neurological disease or injury | 2 | - Bridge program (personalized approach) - Structured-exercise program (group-based) | All participants were re-randomized | All participants were re-randomized to either:   - **Switch** to Relapse Prevention Group - **Switch** to no Relapse Prevention Group (control) | Time and intensity of postintervention physical activity and psychosocial assessments of depression, anxiety, self-efficacy, and function | Cardiovascular fitness measured with the Fatigue Index derived from the Wingate Anaerobic Test | NA |
| Czyz EK et al., 2021(61) | Psychiatrically hospitalized adolescents aged 13 to 17 years presenting with suicide risk concerns | Adolescents | Mental and behavioral health | 2 | - Motivational Interviewing-Enhanced Safety Plan (MI-SP) Plus Text-based support (MI-SP + texts) - MI-SP Only | All participants were re-randomized | All participants were re-randomized to either:   - First stage intervention **augmented** with booster calls - **Continue** first stage intervention (no booster calls) | NA | NA | Primary, secondary, and exploratory/tertiary outcomes were not distinguished.  Mechanisms of change: safety plan use, coping, self-efficacy  Distal: suicidal ideation and behavior |
| Fatori D et al., 2018(62) | Children and adolescents aged 7 to 17 with obsessive compulsive disorder (OCD) | Children and adolescents | Mental and behavioral health | 2 | - Fluoxetine (FLX) - Group cognitive-behavioral therapy (GCBT) | Non-responders are those with less than 50% reduction in baseline Yale-Brown Obsessive Compulsive Scale (Y-BOCS) scores | Non-responders were re-randomized to either:   - **Switch** to the other first-stage treatment - **Augment** intial treatment with the other first-stage treatment   Responders receive no further intervention | Severity of obsessive-compulsive symptoms (Yale-Brown Obsessive-Compulsive Scale (Y-BOCS) at baseline, weeks 7, 14, 21, and 28. | NA | Safety/adverse events |
| Gao K et al., 2020(63) | Adults (18 years or older) with bipolar I or II disorder | Adults | Mental and behavioral health | 2 | - Lithium monotherapy (Li) - Divalproex monotherapy (Div) | Non-responders: Participants who continued to have clinically significant depression (CGI-S-BD depression score >=3 for 2 weeks at any time after the initial 2-week period) | Responders **continue** their first stage intervention  Non-responders were randomized to:   - **Continue** first stage therapy - Mood stabilizer (Li or Div) **augmented** with adjunctive quetiapine (QTP) - Mood stabilizer (Li or Div) **augmented** with adjunctive lamotrigine (LTG) | NA | NA | Primary and secondary outcome measures were not specified. The rates of early termination and time to termination after randomization were used to measure effectiveness of each intervention. Changes from baseline to the end of study in BISS and CGI-S-BD scores were used to measure the efficacy of each intervention. |
| Geng EH et al., 2023(64) | Adults aged 18 years or older living with HIV who had initiated antiretroviral therapy within the previous 90 days | Adults | HIV | 2 | - Standard of Care (SOC) Oureach - Short Message Service (SMS) - Conditional Cash Transfers (CCT) | Responders did not have a retention lapse (missed a scheduled clinic visit by day 14 or more ) | SOC responders **continue** SOC, while SOC non-responders are re-randomized to:   - **Continue** SOC-Outreach - **Switch** to SMS+CCT - **Switch** to peer navigation   Responders to SMS or CCT were re-randomized to either:   - **Continue** with first-stage intervention - **Discontinue** first-stage intervention   Non-responders to SMS or CCT were re-randomized to either:   - **Switch** to SOC-Outreach - Initial intervention **augmented** with the other stage 1 intervention - **Switch** to Peer Navigation | Stage 1: retention in care without lapse (>= 14 days late for an appointment) for 1 year following initial randomization  Stage 2a: Among those with a retention lapse, return to clinic by 1 year following randomization  Stage 2b: Among those with no lapse in year 1 wile in either the SMS or CCT group, retention in care without lapse for 1 year following randomization  Comparision of the 15 adaptive strategies: proportion of time spent actively engaged in care (proportion of days during 2-year follow-up pateint was alive and had not been 14 days or more later for a scheduled appointment) | Stage 1: Composite of viral suppression and retention at the end of year 2  Stage 2a (among those with a lapse by end of 1 year), Stage 2b (among those without a lapse by end of 1 year), full SMART: adjudicated viral suppression 2 years after enrollment | NA |
| Kruse GR et al., 2023(65) | Adults primary care patients who are daily smokers | Adults | Substance use | 2 | - SMS with early assessment - SMS with late assessment | Non-responders: Not achieving 7 days abstinence from cigarettes | Responders receive no further intervention  Non-responders were randomized to either:   - SMS **augmented** with Nicotine replacement therapy (NRT) - SMS **augmented** with NRT + one session with behavioral coach | Self-reported 7 day point prevalent abstinence (PPA) at 12-weeks | Not explicitly labeled as secondary: 7-day PPA at 4- and 8-weeks, number of last 30 days with no smoking, use of cessation treatments, changes in cigarettes per day, motivation, confidence to quit, distress, biochemically verified abstinence | Feasibility, intervention fidelity, intervention engagement, treatment acceptability |
| Lambert SD et al., 2022(66) | Adults with cardiovascular disease who reported moderate stress | Adults | Mental and behavioral health | 2 | - Website only - Website + coach | Non-responders: Stress score did not improve by at least 50% or who were not below the threshold DASS score of 16 | Responders **continue** initial intervention  Non-responders were re-randomized to either:   - **Continue** initial intervention - **Switch** to Website + Motivational Interviewing (MI) | Stress and quality of life | Self-efficacy, patient illness appraisal, individual coping strategies, self-reported activity levels | Feasibility, acceptability, fidelity of the coaching intervention |
| McKay JR et al., 2015(67) | Adults aged 18 to 65 with DSM-IV alcohol or cocaine dependence, who used alcohol or cocaine in the past 3 months. | Adults | Substance use | 2 | Those not engaged at 2 weeks and those who are engaged at 2 weeks, but disengaged in weeks 3-8 are initially randomized to:   - MI-OP: Motivational Interviewing – Intensive Outpatient Program, telephone calls that focused on engagement in IOP - MI-PC: Motivational Interviewing – Patient Choice, telephone calls giving participants the choice between continuing IOP or selecting one of three treatments (cognitive-behavioral therapy (CBT), telephone stepped care (TSC), or medication management (MM)) | Week 2 Non-responders: Failed to attend 2 or more IOP sessions in week 2 or failed to attend the second orientation and the first scheduled IOP session  Week 3-8 Non-responders: Initially engaged but failed to attend any IOP sessions for two consecutive weeks  Week 2 and week 8 Non-responders: Not engaged at week 2 and also failed to attend any IOP sessions in weeks 7 and 8 | Those in the disengaged in weeks 3-8 group are not re-randomized  Responders in the not engaged at 2 weeks group received no further intervention  Non-responders who disengaged at both weeks 2 and 8 were randomized to either:   - **Continue** or **switch** to MI-PC - **Switch** to no further outreach | Alcohol and drug use within each 30-day period of follow-up. For participants with alcohol dependence, the outcomes were dichotomous measures of any alcohol and any heavy alcohol use, and continuous measures of percent days alcohol and heavy alcohol use from the TLFB. Heavy alcohol use was defined as ≥ 5 drinks in a day for men, ≥ 4 drinks for women. For participants with cocaine dependence, the outcomes were a dichotomous measure of any cocaine use and percent days cocaine use from the TLFB, and cocaine urine toxicology | NA | Attendance |
| Morgenstern J et al., 2021(68) | Individuals ages 18 to 75 who meet the criteria for alcohol use disorder (AUD) | Adults | Substance use | 2 | - MI: Motivational interviewing - BA Plus: An additional Brief Advice session | Responders were defined as reducing their drinking to the level of the NIAAA safe drinking guidelines, i.e. for men under the age of 65; <=14 standard drinks per week and <5 standard drink on one occasion; women and men over the age of 65 <= 7 standard drinks per week and <4 standard drinks on one occasion. | Responders receive no further intervention  Non-responders to BA Plus were re-randomized to either:   - **Switch** to two sessions of MI - **Switch** to five sessions of MI with four sessions of behavioral self-control therapy (BSCT)   Non-responders to MI were randomized to either:   - **Continue** MI for an additional session - **Switch** to four sessions of BSCT   Responders did not receive further treatment | NA | NA | Primary, secondary, and exploratory/tertiary outcomes were not distinguished.  Sum of standard drinks (SSD) and count of heavy drinking days (HDD) for the week prior to the affiliated assessment period (baseline, week 4, week 8, and week 13). |
| Naar-King S et al., 2016 | African American adolescents with obesity | Adolescents | Overweight and obesity | 2 | - Home-based Motivational Interviewing and Skills (HB-MIS) - Office-based Motivational Interviewing and Skills (OB-MIS) | Non-responders: Weight loss of less than 3%  Responders: Weight loss greater than or equal to 3% | Non-responders were randomized to either:   - **Switch** to continued skills (CS) - **Switch** to contingency management (CM)   Responders received Relapse Prevention (RP) | Percent overweight | Treatment engagement | NA |
| Pelham Jr WE et al., 2016(70) | Children aged 5-12 years old with ADHD | Children | Mental and behavioral health | 2 | - BehFirst: Low-intensity behavior modification - MedFirst: Low dose medication | Non-responder: child experienced continued impairment in the school and/or home setting | All responders continue initial intervention and reassess monthly; randomize if deteriorate  Non-responders to BehFirst are randomized to either:   - B-then-M: **Augment** with medication - B-then-B: **Step up** behavior modification   Non-responders to MedFirst are randomized to either:   - M-then-B: **Augment** with behavior modification - M-then-M: **Step up** the medication dose | Classroom rule violations | Number of out-of-class disciplinary events, parent teacher ratings | Fidelity |
| Pistorello J et al., 2017(71) | College students aged 18 to 25 years enrolled at a mid-size public university seeking services at the campus clinic and reporting moderate to severe suicide ideation | Young adults | Mental and behavioral health | 2 | - Collaborative Assessment and Management of Suicidality (CAMS): suicide-specific, collaborative approach to identify suicidal “drivers” that are treated over the course of care - Treatment-as-usual (TAU): customary treatment a study counselor would use in regular clinical practice | Responders: an Improvement score of ≤ 2 (much improved or very much improved) combined with a Severity score of ≤ 3 (Not at all suicidal, minimally suicidal, or mildly suicidal) | Responders proceed to monitoring, post and follow-up  Non-responders were randomized to either:   - **Continue** or **switch** to CAMS - **Switch** to comprehensive Dialectical Behavioral Therapy (DBT): individual and group sessions along with peer consultation meetings for therapists and as-needed phone/text coaching | Feasibility of the study in terms of participant recruitment (numbers and diversity) and clinician adherence to treatment, and the acceptability of the adaptive strategies in terms of clients’ attendance, treatment dropout, and response to treatment and clients’ and clinicians’ satisfaction with treatment | NA | Stage 2 satisfaction and treatment response |
| Sauer-Zavala S et al., 2022(72) | Adults seeking treatment for anxiety and depressive disorders | Adults | Mental and behavioral health | 2 | - Standard condition (fixed standard order as described in the Unified Protocol manual) - Capitalization condition (strength-based sequencing) - Compensation condition (deficit-based sequencing) | All participants were randomized to the second stage which was solely based on session timing | All participants were re-randomized to **switch** to:   - Brief treatment (6 sessions total) - Full treatment (12 sessions total) | Primary outcomes were not distinguished | Secondary outcomes were not distinguished | Clinical severity rating (CSF), Overall anxiety severity and impairment scale (OASIS), Overall depression severity and impairment scale (ODSIS), feasibility, acceptability, satisfaction |
| Schlam TR et al., 2024(73) | Adult primary care patients who smoked at least 4 cigarettes per day consistently for the last 6 months and wanted to quit smoking within the last month | Adults | Substance use | 2 | All participants initially received a smoking cessation treatment.  Participants that relapsed after the initial treatment were randomized to either:   - Preparation Treatment (Active) - Recycling Treatment - Quitline referral (control) | Non-responders (relapse) to be eligible for the first randomization: smoking on seven consecutive days at any time up to 6 months after the initial quit attempt.  Transition to the second randomization depended on participants’ decision to attempt quitting again. | Controls do not receive further intervention  Optional randomization for those initially randomized to Preparation or Recycling who also want to make another quit attempt.  Participants received 8 weeks of nicotine patches plus mini-lozenges and were randomly assigned in a 2x2 factorial design to **switch** to:   - Skill training (on/off) - Supportive counseling (on/off) | CO-confirmed 7-day point-prevalence abstinence 14 months post-phase 2 treatment initiation | Self-reported 7-day point-prevalence abstinence at week 26 post the phase 3 TQD | Abstinence at earlier time points, treatment re-engagement |
| Sikorskii A et al., 2023(74) | Survivors of solid tumors who had completed or were scheduled to complete curative-intent adjuvant chemotherapy or chemoradiation, without additional planned cancer treatments except for radiation therapy, hormonal therapy, or trastuzumab for breast cancer | Adults | Cancer | 2 | - Symptom Management and Survivorship Handbook (SMSH) - SMSH combined with Telephone Interpersonal Counseling (TIP-C) | Non-responders to SMSM: Survivors who started at severe at baseline and ended at moderate or none/mild at week four, and survivors who started at moderate and ended at none/mild, were categorized as responders after four weeks of the SMSH. | Those in the TIP-C group did not receive additional intervention  Responders to SMSH **continued** SMSH  Non-responders to SMSH were re-randomized to either:   - **Continue** SMSH alone - SMSH **augmented** with TIP-C | Depressive symptoms and a summary index of severity of other symptoms during weeks one to 13 | Secondary outcomes were not distinguished | PROMIS short forms for physical function (4 items), anxiety (8 items) |
| Stanger C et al., 2020(75) | Youth with cannabis use disorder | Youth | Substance use | 2 | - STANDARD contingency management (CM) alone - STANDARD CM + working memory training (WMT) | Responders: Youth who were abstinent in Week 4  Non-responders: Youth who were not abstinent in Week 4 | Responders **continued** their Phase 1 intervention  Non-responders were randomized to either:   - **Continue** standard CM - **Step up** to enhanced CM   Assignment to WMT or no WMT was unchanged in stage 2 | Primary outcomes were not distinguished | Secondary outcomes were not distinguished | Cannabis use and abstinence, working memory |
| Protocol + primary analysis papers | | | | | | | | | | |
| Primary: Fortney JC et al., 2021(76)  Protocol: Fortney JC et al., 2020(77) | Adults treated at primary care clinics who screen positive for posttraumatic stress disorder and/or bipolar disorder | Adults | Substance use | 2 | - Telepsychiatry/Telepsycholoy-enhanced referral (TER): Telepsychiatrists assumed responsibility for treatment - Telepsychiatry Collaborative Care (TCC): Telepsychiatrists provided consultation to the primary care team | Non-responders: Patients assigned to TER with <= 2 telehealth encounters | Everyone randomized to TCC and TER responders received no further intervention  TER non-responders were randomized to either:   - **Continue** TER - **Switch** to Phone Enhanced Referral (PER): telephone outreach by pscyhologist aimed to increase patient engagement in treatment | Veterans RAND 12-item Health Survey Mental Component Summary (MCS) score | PTSD Checklist-5 (PCL-5) for PTSD, Hopkins Symptom Checklist (SCL-20) for depression, Altman Mania Rating Scale (AMRS) for mania, Internal State Scale (ISS) for mood state, General Anxiety Disorder-7 for general anxiety, Recovery Assessment Scale, and number of moderate and severe adverse effects from psychotropic medications | Accessibility, engagement, and adherence |
| Primary: Gonze BB et al., 2020(78)   Protocol: Morais Pereira Simões et al., 2019(79) | Insufficiently active adults | Adults | Mental and behavioral health | 2 | - Pacer App only - App+tailored messages - Control group | To define responders and non-responders in this study, a linear regression was fitted for each participant with the relationship between weeks on the x-axis and the number of steps per day on the y-axis. Responders were with any positive slope at the end of the 12-week first stage of the intervention. Those with zero or negative slopes were considered as non-responders. | Responders **continue** their initial intervention  Non-responders to either active treatment were randomized to **switch** to one of the following which was not the same as the assigned first stage intervention:   - Pacer App only - App+tailored messages   App + tailored messages + gamification | NA | NA | Feasibility, participant perceptions |
| Primary: Gunlicks-Stoessel M et al., 2019(80)  Protocol: Gunlicks-Stoessel M et al., 2016(81) | Adolescents aged 12 to 17 years who had a DSM-IV-TR diagnosis of major depressive disorder, dysthymia, or depressive disorder | Adolescents | Mental and behavioral health | 2 | - Interpersonal Psychotherapy for Depressed Adolescents (IPT-A) assessed at 4 weeks (early): 12-session evidence-based psychotherapy aimed to decrease depressive symptoms by helping adolescents improve their relationships and interpersonal interactions - Interpersonal Psychotherapy for Depressed Adolescents (IPT-A) assessed at 8 weeks (late) | Non-responders: <20% reduction in Hamilton Rating Scale for Depression (HRSD) at week 4 or <40% reduction in HRSD score at week 8) | Responders **continue** IPT-A  Non-responders are randomized to either:   - **Step up** frequency of IPT-A - **Augmentation** of IPT-A with fluoxetine | Children’s Depression Rating Scale-Revised (CDRS-R) | (Children’s Global Assessment Scale) CGAS, (Social Adjustment Scale-Self Report) SAS-SR | Safety (Harm and suicide-related adverse events) |
| Primary: Igudesman D et al., 2023(82)  Protocol: Corbin KD et al., 2022(83) | Young adults with type 1 diabetes with overweight or obesity | Young adults | Overweight or obesity | 3 | - Hypocaloric Low-carbohydrate diet - Hypocaloric Look AHEAD diet (moderate low-fat) - Mediterranean diet | Non-responders: <2% weight reduction (unless weight loss resulted in a body mass index <25 kg/m2), HbA1c increase ≥0.5% and self-reported increased or problematic hypoglycaemia and self-reported diet unacceptability | All non-responders were randomized to **switch** one of the alternative diets | Change in weight (kg), haemoglobin A1c (HbA1c) level, and percentage of time below range (%TBR; <70 mg/dl) as assessed by continuous glucose monitoring (CGM) at baseline and following each of three, 3-month, dietary periods | Change in percentage body fat as assessed by dual-energy X-ray absorptiometry (DXA), percentage of time in target glucose range (%TIR; 70-180 mg/dl) and %TBR (<54 mg/dl) | Percentage of time above range (%TAR; glucose levels 181-250 mg/dL and >250 mg/dL), coefficient of variation (CV) in glucose levels as measured by CGM |
| Primary: Karp JF et al., 2019(84)  Protocol: Karp JF et al., 2016(85) | Older adults (aged >=60) with knee osteoarthritis | Older adults | Osteoarthritis | 2 | - Cognitive Behavioral Therapy (CBT) - Physical Therapy (PT) - Enhanced Usual Care (EUC) | Non-responders: Lack of clinically significant response to the interventions defined as (1) much better or (2) very much better on the P-GIC | Everyone randomized to EUC and responders to CBT and PT receive no further intervention  CBT and PT non-responders are re-randomized to either:   - **Continue** stage 1 treatment - **Switch** to the other treatment | Patient-reported global impression of change (P-GIC) | Changes in depressive symptoms (PHQ-9), changes in anxiety symptoms (GAD-7), pain (numeric rating scale) | NA |
| Primary: Morin CM et al., 2020(86)  Protocol: Morin CM et al., 2016(87) | Individuals with insomnia | Adults | Sleep | 2 | - Behavioral Therapy (BT): sleep restriction and stimulus control procedures - Zolpidem: medication therapy | Responders: reduction of 8 points or more on the Insomnia Severity Index (ISI) compared with baseline | Responders receive no further intervention  Non-responders to Zolpidem were re-randomized to either:   - **Continue** with Zolpidem - **Switch** to Trazodone   Non-responders to BT were randomized to either:   - **Continue** with BT - **Switch** to Cognitive Therapy (CT) | Treatment response and remission rates, defined by the Insomnia Severity Index total score. | Secondary endpoints were derived from sleep diaries | NA |
| Primary: Mustanski B et al., 2023(88)  Protocol: Mustanski B et al., 2020(89) | Adolescent men who have sex with men (AMSM) | Adolescents | HIV | 2 | All participants receive SMART Sex Ed (SSE) prior to first randomization.  Respondents are randomized to:   - follow-up only - SMART Squad   Nonrespondents are randomized to:   - SMART Squad + Booster 1 (embedded regimes 1 and 2) - SMART Sex Ed (SSE2.0) + Booster 1 (embedded regimes 3 and 4) | Those originally randomized to follow-up only or SMART Squad are not re-randomized. Non-responders to SMART SSE2.0 or SMART Squad + Booster 1 are re-randomized.  Response is defined as meeting each of these 3 criteria: (1) 100% condom use, if the participant is sexually active in the assessment period, (2) intentions for condom use during all instances of penetrative sex (regardless of reported sexual activity), and (3) reporting a high degree of self-efficacy for achieving condom use during all instances of penetrative sex (regardless of sexual reported activity) | Responders **augment** their first intervention (SMART Squad or SSE2.0) + Booster 2 + follow-up assessment  Non-responders **continue** their embedded regime (ER) path:   - ER1 continues to SMART Sessions - ER2 continues to SMART Squad Booster 2 + access to SMART Squad - ER3 continues to SMART Squad - ER4 continues to SMART Sessions | Sexual risk, condom use intentions and self-efficacy, HIV testing | HIV knowledge, motivation and behavior skills, condom errors | Substance use, PrEP |
| Primary: Patrick ME et al., 2021(90)  Protocol: Patrick ME et al., 2020(91) | Incoming first year college students | Young adults | Substance use | 2 | - Personalized Normative Feedback (PNF) + Self-monitoring (SM) prior to first semester (early group) - PNF + SM during first month of first semester (late group) - Assessment only (control) | Non-responders: Report of heavy drinking (two or more instances of binge drinking or one or more instance of high-intensity drinking) | Assessment only controls **continue** assessment only  Responders in the early and late group **continue** SM  Non-responders are re-randomized to either:   - **Switch** to resource email - **Switch** to online health coach | Frequence of binge drinking | Alcohol-related consequences, health services utilization | NA |
| Primary: Schmitz JM et al., 2024(92)  Protocol: Schmitz JM et al., 2018(93) | Treatment-seeking adults aged 18 to 60 years who met current (past month) DSM-5 criteria for cocaine use disorder (CUD) | Adults | Substance use | 2 | - Acceptance and Commitment Therapy (ACT) + Contingency Management (CM) - Drug counseling (DC) + CM | Responders: submit 6 consecutive cocaine-negative urine drug screens | Responders continue phase 1 intervention in phase 2  Non-responders to ACT are re-randomized to either   - ACT + CM **augmented** with Modafinil (MOD) - ACT + CM **augmented** with placebo (PLA)   Non-responders to DC + CM were randomized to either   - DC + CM **augmented** with MOD - DC + CM **augmented** with PLA | Response rate defined as two weeks of consecutive abstinence by the end of Phase 1 and proportion of cocaine-negative UDS for phase 1 and 2 | Reduction in cocaine use | Retention, adverse events, adherence |
| Primary: Schoenfelder EN et al., 2019(94)   Protocol: Chronis-Tuscano A et al., 2016(95) | Mother-child dyads with ADHD | Mother-child dyads | Mental and behavioral health | 2 | - Maternal stimulant medication (MSM) - Behavioral parent training (BPT) | All families were re-randomized regardless of initial response | All families re-randomized to either:   - **Continue** with first-stage intervention - **Augment** first-stage intervention with the other first-stage intervention | NA | NA | Feasibility, acceptability |
| Primary: Sherwood NE et al., 2022(102)  Protocol : Sherwood NE et al., 2016(103) | Adults aged 21 to 70 years with obesity (BMI >= 30 and <= 45 kg/m^2^) | Adults | Overweight and obesity | 2 | - State-of-the-art Behavioral Weight Loss Treatment (SBT) with early assessment: assessment at 3 weeks - SBT with late assessment: assessment at 7 weeks | Non-responders to Early TRA: lost less than 2.5% of their session 1 weight by Session 3 or 28 days following Session 1, which ever came first.  Non-responders to Late TRA: lost less than 5.0% of their session 1 weight by Session 7 or 63 days following Session 1, whichever came first. | Responders **continued** with SBT  Non-responders were randomized to either **switch** to:   - Portion-controlled meals (PCM) - Acceptance-based Behavioral Treatment (ABT) | Body weight change from baseline at 6 months and 18 months | NA | Adherence, process/implementation, avoidance of treatment contamination, satisfaction and perceived helpfulness, safety/adverse events |
| Primary: Smith SN et al., 2022(96)  Protocol: Kilbourne AM et al., 2018(97) | School professionals (SPs) in high schools | Medical centers, practices, or providers | Mental and behavioral health | 2 | - Replicating Effective Programs (REP) - REP + Coaching | Non-responders: Any SP at a particular school not providing >=3 CBT components to >=10 students or SPs at a particular school reporting on average >2 barriers to CBT delivery or any SP at a particular school failed to complete the monitoring assessment | Responders continue phase 1 intervention in phase 2  Non-responders are re-randomized to either:   - **Continue** phase 1 intervention - Phase 1 intervention **augmented** with facilitation   All participants continue REP in phase 3 | Total number of SP-reported CBT sessions delivered to students by SPs over the 18-month study period | Number of CBT sessions delivered by type of session: group vs. individual brief vs. individual full | Implementation |
| Primary: Somers TJ et al., 2023(98)  Protocol : Kelleher SA et al., 2017(99) | Women with stage I-IIIC breast cancer, within 2 years of diagnosis or recurrence, aged 18 or older, with a life expectancy >= 12 months, and with worst pain severity >=5 out of 10 | Adults | Pain | 2 | - Pain Coping Skills Training Full (PCST-Full): Five weekly 60-minute sessions - Pain Coping Skills Training Brief (PCST-Brief): One 60-minute session | Non-responders: <30% reduction or an increase in pain severity after first intervention | Responders to PCST-Full were re-randomized to either:   - **Continue** PCST-Full (maintenance) - No further intervention   Non-responders to PCST-Full were re-randomized to either:   - **Step up** to PCST-Plus: two additional sessions - **Continue** PCST-Full (maintenance)   Responders to PCST-Brief were randomized to either:   - **Continue** PCST-Brief (maintenance) - No further intervention   Non-responders to PCST-Brief were randomized to either:   - **Step up** to PCST-Full - **Continue** PCST-Brief (maintenance) | Brief Pain Inventory – pain severity subscale (percent change in pain severity from Assessment 1 to Assessment 2) | NA | Adherence, skills practice |
| Primary: Wyatt G et al., 2021(100)  Protocol: Sikorskii et al., 2017(101) | Dyads of patients with solid tumor cancers and their friend/family caregivers | Caregiver-individual dyads | Cancer | 2 | - Reflexology - Meditative practices - Control (usual care) | Responders: had improvement in fatigue from moderate to mild or severe to moderate | Those receiving usual care **continued** usual care  Responders to reflexology or meditative practices **continued** their initial treatment  Non-responders to reflexology were re-randomized to either:   - **Continue** reflexology - **Augment** reflexology with meditative practices   Non-responders to meditative practices were re-randomized to either:   - **Continue** meditative practices - **Augment** meditative practices with reflexology | Fatigue severity from the 9-item Brief Fatigue Inventory | Summed Symptom Severity Index (depression, anxiety, and the severity index of other symptoms) | Physical function and cancer and its treatment |

**Table S3. Intervention components**

| Domain | Intervention type | | | | | | | | | | | Intervention delivery | |
| --- | --- | --- | --- | --- | --- | --- | --- | --- | --- | --- | --- | --- | --- |
| First author, year | Biomedical/clinical | | Behavioral, lifestyle, and psychosocial interventions | | | | | Organizational interventions | |  |  |  |  |
|  | Pharmacological | Devices and procedures | Psychological and psychotherapy | Peer and social support | Financial and economic incentives | Education, self-management, and adherence | Lifestyle modification | Patient navigation | Care coordination | Rehabilitation and therapeutic physical intervention | Implementation | Digital Health | Direct touch or traditional |
| Protocol Papers | | | | | | | | | | | | | |
| Abuogi LL et al., 2023(1) | No | No | No | Yes | Yes | No | No | Yes | No | No | No | Yes | Yes |
| Arean P et al., 2021(2) | No | No | Yes | No | No | No | No | No | No | No | No | Yes | No |
| August GJ et al., 2016(3) | No | No | Yes | No | No | Yes | No | No | No | No | No | No | Yes |
| Auyeung SF et al., 2009(4) | Yes | No | No | No | No | No | No | No | No | No | No | No | Yes |
| Bahraini NH et al., 2020(5) | No | No | No | No | No | No | No | No | No | No | Yes | Yes | Yes |
| Belzer ME et al., 2018(6) | No | No | No | Yes | Yes | Yes | No | No | Yes | No | No | Yes | No |
| Berget C et al., 2019(7) | No | No | Yes | No | No | Yes | No | No | No | No | No | Yes | No |
| Buchholz SW et al., 2020(8) | No | No | Yes | Yes | No | Yes | Yes | No | No | No | No | Yes | Yes |
| Carr E et al., 2024(9) | No | No | No | No | No | Yes | Yes | No | No | Yes | No | Yes | No |
| Comins CA et al., 2019(10) | Yes | No | No | Yes | No | Yes | No | Yes | No | No | Yes | No | Yes |
| Davis-Ewart L et al., 2023(11) | No | No | Yes | No | Yes | No | No | Yes | No | No | No | Yes | No |
| Doorenbos AZ et al., 2023(12) | No | No | Yes | No | No | Yes | No | No | No | No | No | Yes | Yes |
| Drake CL et al., 2022(13) | No | No | Yes | No | No | Yes | No | No | No | No | No | Yes | Yes |
| Edelman EJ et al., 2021(14) | Yes | No | No | No | Yes | Yes | Yes | No | No | No | No | No | Yes |
| Eldridge-Smith ED et al., 2022(15) | No | Yes | Yes | No | No | Yes | No | No | No | No | No | Yes | Yes |
| Fernandez ME et al., 2020(16) | No | No | Yes | No | No | Yes | Yes | No | No | No | Yes | Yes | Yes |
| Flynn D et al., 2018(17) | No | No | Yes | No | No | Yes | Yes | No | No | Yes | No | No | Yes |
| Fox CK et al., 2024(18) | Yes | No | No | No | No | Yes | Yes | No | No | No | No | No | Yes |
| Fritz JM et al., 2020(19) | No | No | Yes | No | No | Yes | No | No | No | Yes | No | No | Yes |
| Fu SS et al., 2017(20) | Yes | No | No | No | No | Yes | Yes | No | Yes | No | No | Yes | Yes |
| Germeroth LJ et al., 2019(21) | No | No | Yes | No | No | Yes | Yes | No | No | No | No | Yes | Yes |
| Hassett AL et al., 2023(22) | Yes | No | Yes | No | No | Yes | No | No | No | Yes | No | Yes | Yes |
| Hibbard JC et al., 2018(23) | No | Yes | No | No | No | No | No | No | No | No | No | No | Yes |
| Inwani I et al., 2017(24) | No | No | No | No | Yes | Yes | No | Yes | Yes | No | Yes | Yes | Yes |
| Jain S et al., 2023(25) | Yes | No | No | No | No | No | No | No | No | No | No | No | Yes |
| Johnson JE et al., 2018(26) | No | No | No | No | No | No | No | No | No | No | Yes | Yes | No |
| Kilbourne AM et al., 2014(27) | No | No | No | No | No | Yes | No | No | No | No | Yes | No | Yes |
| Kopelowicz A et al., 2023(28) | No | No | No | Yes | No | Yes | Yes | Yes | No | No | No | No | Yes |
| Kor PP et al., 2023(29) | No | No | Yes | No | No | Yes | No | No | No | No | No | Yes | No |
| Levy R et al., 2019(30) | Yes | No | Yes | No | No | No | No | No | No | No | No | No | Yes |
| Li X et al., 2021(31) | Yes | Yes | No | No | No | No | No | No | No | No | No | No | Yes |
| Lion KC et al., 2023(32) | No | No | No | No | No | No | No | No | No | No | Yes | Yes | No |
| Liu H et al., 2021(33) | No | No | No | No | No | Yes | No | No | No | No | No | Yes | No |
| Markland AD et al., 2023(34) | No | No | No | No | No | Yes | No | No | No | Yes | No | Yes | Yes |
| Micheletti RG et al., 2020(35) | Yes | No | No | No | No | No | No | No | No | No | No | No | Yes |
| Nelson B et al., 2018(36) | Yes | No | Yes | No | No | No | No | No | Yes | No | No | Yes | Yes |
| O'Keefe VM et al., 2019(37) | No | No | Yes | Yes | No | Yes | No | Yes | Yes | No | No | No | Yes |
| Osilla KC et al., 2023(38) | No | No | Yes | No | No | Yes | No | No | No | No | No | Yes | Yes |
| Peter SC et al., 2023(39) | No | No | Yes | No | Yes | Yes | No | No | No | No | No | No | Yes |
| Peterson BS et al., 2021(40) | Yes | No | Yes | No | No | No | No | No | No | No | No | Yes | Yes |
| Pfammatter AF et al., 2019(41) | No | No | Yes | No | No | Yes | Yes | No | No | No | No | Yes | Yes |
| Quanbeck A et al., 2020(42) | No | No | No | No | No | No | No | No | No | No | Yes | Yes | Yes |
| Rabin BA et al., 2023(43) | No | No | No | Yes | No | Yes | No | Yes | Yes | No | Yes | Yes | Yes |
| Sabri B et al., 2021(44) | No | No | Yes | No | No | Yes | No | No | No | No | No | Yes | No |
| Schlechter CR et al., 2023(45) | No | No | Yes | No | No | Yes | Yes | Yes | No | No | No | Yes | Yes |
| Skolasky RL et al., 2020(46) | No | No | Yes | No | No | No | No | No | No | Yes | No | No | Yes |
| Smith SK et al., 2021(47) | No | No | Yes | No | No | Yes | No | No | Yes | No | No | Yes | Yes |
| Sripada RK et al., 2023(48) | No | No | Yes | No | No | Yes | No | No | No | No | No | Yes | Yes |
| Tamura K et al., 2020(49) | No | No | No | No | No | Yes | Yes | No | No | No | No | Yes | No |
| van Heerden A et al., 2023(50) | No | No | No | No | Yes | No | No | Yes | Yes | No | Yes | Yes | Yes |
| Velloza J et al., 2022(51) | No | No | Yes | Yes | No | Yes | No | No | No | No | No | Yes | Yes |
| Walton MA et al., 2023(52) | No | No | Yes | No | No | Yes | No | No | No | No | No | Yes | Yes |
| Wan Y et al., 2023(53) | No | No | No | No | No | Yes | No | No | Yes | No | No | Yes | Yes |
| Wilroy J et al., 2023(54) | No | No | Yes | No | No | Yes | Yes | No | No | Yes | No | Yes | No |
| Windsor L et al., 2022(55) | No | No | Yes | Yes | No | Yes | No | Yes | No | No | No | Yes | Yes |
| Yan X et al., 2022(56) | No | No | No | No | No | Yes | Yes | No | No | No | No | Yes | Yes |
| Zhao SZ et al., 2022(57) | Yes | No | Yes | Yes | Yes | Yes | Yes | No | No | No | No | Yes | Yes |
| Zhou G et al., 2020(58) | No | Yes | No | No | No | No | No | No | No | No | No | No | Yes |
| Zullig LL et al., 2021(59) | No | No | Yes | No | No | Yes | No | No | Yes | No | Yes | Yes | Yes |
| Primary analysis papers | | | | | | | | | | | | | |
| Butzer JF et al., 2023(60) | No | No | No | No | No | Yes | Yes | No | No | No | No | No | Yes |
| Czyz EK et al., 2021(61) | No | No | Yes | No | No | Yes | No | No | No | No | No | Yes | Yes |
| Fatori D et al., 2018(62) | Yes | No | Yes | No | No | No | No | No | No | No | No | No | Yes |
| Gao K et al., 2020(63) | Yes | No | No | No | No | No | No | No | No | No | No | No | Yes |
| Geng EH et al., 2023(64) | No | No | No | No | Yes | Yes | No | Yes | No | No | No | Yes | Yes |
| Kruse GR et al., 2023(65) | Yes | No | No | No | No | Yes | No | No | No | No | No | Yes | Yes |
| Lambert SD et al., 2022(66) | No | No | Yes | Yes | No | Yes | Yes | No | No | No | No | Yes | Yes |
| McKay JR et al., 2015(67) | Yes | No | Yes | No | No | Yes | No | No | No | No | No | Yes | Yes |
| Morgenstern J et al., 2021(68) | No | No | Yes | No | No | Yes | No | No | No | No | No | No | Yes |
| Naar-King S et al., 2016(69) | No | No | Yes | Yes | No | Yes | Yes | No | No | No | No | No | Yes |
| Pelham Jr WE et al., 2016(70) | Yes | No | Yes | No | No | Yes | No | No | No | No | No | No | Yes |
| Pistorello J et al., 2017(71) | No | No | Yes | No | No | Yes | No | No | No | No | No | No | Yes |
| Sauer-Zavala S et al., 2022(72) | No | No | Yes | No | No | No | No | No | No | No | No | No | Yes |
| Schlam TR et al., 2024(73) | Yes | No | Yes | No | No | Yes | Yes | No | No | No | No | Yes | No |
| Sikorskii A et al., 2023(74) | No | No | Yes | No | No | Yes | No | No | No | No | No | No | Yes |
| Stanger C et al., 2020(75) | No | No | No | No | Yes | Yes | No | No | No | No | No | No | Yes |
| Protocol + Primary analysis papers | | | | | | | | | | | | | |
| Primary: Fortney JC et al., 2021(76)  Protocol: Fortney JC et al., 2020(77) | No | No | Yes | No | No | Yes | Yes | No | Yes | No | No | Yes | No |
| Primary: Gonze BB et al., 2020(78)   Protocol: Morais Pereira Simões et al., 2019(79) | No | No | No | Yes | No | Yes | Yes | No | No | No | No | Yes | No |
| Primary: Gunlicks-Stoessel M et al., 2019(80)  Protocol: Gunlicks-Stoessel M et al., 2016(81) | Yes | No | Yes | No | No | No | No | No | No | No | No | No | Yes |
| Primary: Igudesman D et al., 2023(82)  Protocol: Corbin KD et al., 2022(83) | No | No | No | No | No | Yes | Yes | No | No | No | No | Yes | Yes |
| Primary: Karp JF et al., 2019(84)  Protocol: Karp JF et al., 2016(85) | No | No | Yes | No | No | No | No | No | Yes | Yes | No | No | Yes |
| Primary: Morin CM et al., 2020(86)  Protocol: Morin CM et al., 2016(87) | Yes | No | Yes | No | No | No | No | No | No | No | No | No | Yes |
| Primary: Mustanski B et al., 2023(88)  Protocol: Mustanski B et al., 2020(89) | No | No | Yes | Yes | No | Yes | No | Yes | No | No | No | Yes | No |
| Primary: Patrick ME et al., 2021(90)  Protocol: Patrick ME et al., 2020(91) | No | No | Yes | No | No | Yes | No | No | No | No | No | Yes | No |
| Primary: Schmitz JM et al., 2024(92)  Protocol: Schmitz JM et al., 2018(93) | Yes | No | Yes | No | Yes | Yes | No | No | No | No | No | No | Yes |
| Primary: Schoenfelder EN et al., 2019(94)   Protocol: Chronis-Tuscano A et al., 2016(95) | Yes | No | Yes | No | No | No | No | No | No | No | No | No | Yes |
| Primary: Sherwood NE et al., 2022(102)  Protocol : Sherwood NE et al., 2016(103) | No | No | Yes | No | No | Yes | Yes | No | No | No | No | No | Yes |
| Primary: Smith SN et al., 2022(96)  Protocol: Kilbourne AM et al., 2018(97) | No | No | Yes | No | No | No | No | No | No | No | Yes | No | Yes |
| Primary: Somers TJ et al., 2023(98)  Protocol : Kelleher SA et al., 2017(99) | No | No | Yes | No | No | Yes | No | No | No | No | No | Yes | Yes |
| Primary: Wyatt G et al., 2021(100)  Protocol: Sikorskii et al., 2017(101) | No | No | Yes | No | No | Yes | No | No | No | Yes | No | No | Yes |

**TABLE S4. Sample size considerations, blinding, and randomization**

| **SR_number** | **First author** | **Was a sample size calculation reported?** | **What is the target sample size?** | **Did the sample size calculation account for the multiple stages of randomization?** | **Blinding** | **Randomization procedures** |
| --- | --- | --- | --- | --- | --- | --- |
| **Protocol papers** | | | | | | |
| 1296 | Abuogi LL et al., 2023(1) | Yes | 880 | Yes | Study staff and participants not blinded; principal investigators and analysts blinded | Stratified block randomization. 1:1 first stage allocation ratio. 1:1:1 second stage allocation ratio |
| 1105 | Arean P et al., 2021(2) | Yes | 1000 | Yes | Assessors will be blinded | Stage 1 randomization will be stratified systemic randomization |
| 1247 | August GJ et al., 2016(3) | No | 100 | NA | NA | NA |
| 1072 | Auyeung SF et al., 2009(4) | Yes | 70 | Yes | Double-blind | Stratified randomization |
| 1102 | Bahraini NH et al., 2020(5) | Yes | 140 | Yes | NA | First stage stratified by facility complexity level and performance level at baseline; second stage randomization stratified by facility complexity and performance level; stage 3 randomized stratified by facility complexity. |
| 1290 | Belzer ME et al., 2018(6) | No | 190 | NA | NA | NA |
| 1129 | Berget C et al., 2019(7) | No | 90 | NA | NA | NA |
| 1035 | Buchholz SW et al., 2020(8) | Yes | 312 | Yes | Investigators were blinded to randomization table | Block-randomization at each step |
| 1082 | Carr E et al., 2024(9) | Yes | 117 | Yes | Outcome assessors and data analysts will be blinded to all treatment allocations | A simple equal allocation randomization at the individual level using a computer-generated randomized list will be used |
| 1094 | Comins CA et al., 2019(10) | Yes | 800 | Yes | NA | Randomization is achieved through a blocked design utilizing permuted blocks of random sizes. Randomization is 1:1 at the first and second stages of randomization |
| 1147 | Davis-Ewart L et al., 2023(11) | Yes | 70 | NA | Assessors were blinded | First stage randomization was stratified and used permuted blocks for 1:1 allocation; second stage randomization was not stratified and interventions were allocated 1:1 |
| 1182 | Doorenbos AZ et al., 2023(12) | Yes | 366 | Yes | NA | Both randomizations were stratified with permuted block randomization 1:1:1 |
| 1065 | Drake CL et al., 2022(13) | Yes | 1000 | Yes | Patients are blinded to the active therapy. Therapists are not blinded. Outcomes will be linked to a blinded group variable, which will be unblinded after primary analyses. | Patients are randomized 1:1 at each step using block randomization (step 1 uses 50-person blocks; step 2 uses six-person blocks). Only the study coordinator accesses the allocation sequence and assigns patients to groups. |
| 1044 | Edelman EJ et al., 2021(14) | Yes | 632 | Yes | This is a non-blinded study | In the first stage, participants are randomized in a 1:1 ratio and randomization is stratified based on site and the Heaviness of Smoking Index. In the second stage, re- randomization is stratified based on site and first stage treatment. Permuted block randomization sequences are implemented in REDCap |
| 1036 | Eldridge-Smith ED et al., 2022(15) | Yes | 384 | Yes | All study personnel, except the treatment providers and the team members who employ randomization, are blinded to subject randomization. Study coordinators are unblinded at the end of the follow-up phase | First- and second-stage randomizations use a minimization method, a modified, adaptive randomization procedure that ensures treatment conditions are balanced in regard to pre-treatment stratification variables. The variables used in randomization include age, sex, insomnia severity, pre-PAP AHI, and objective PAP  adherence. Randomization is performed independently at each site by one designated study staff via a computer-based Fortran program |
| 1101 | Fernandez ME et al., 2020(16) | Yes | 6000 | Yes | NA | All randomizations used random permuted blocks with random block size, stratification of clinics by CHC and clinic size, and patient-level stratification by clinic and treatment groups of earlier phases of the SMART. Allocation ratios were 4:1 TM versus CO and 1:1 TM-Cont versus TM+MAPS |
| 1053 | Flynn D et al., 2018(17) | Yes | 280 | Yes | NA | NA |
| 1047 | Fox CK et al., 2024(18) | Yes | 150 | Yes | For the first randomization, all participants will be blinded to their treatment arm for the duration of the study. Because they will know if phentermine is started, study staff collecting primary outcomes will be blinded to participant assignment only up until 12 weeks. For the second randomization, participants and study staff collecting primary outcomes will be blinded to phentermine/placebo | Using permuted blocks of size 2, 4, or 6, each participant will be randomized 1:1 in stage 1 and re-randomized 1:1 in stage 2. Randomization and treatment decision rules will be built into a secure, web-based application (Research Electronic Data Capture; REDCap), and each participant will be sequentially assigned a randomized number. Randomization codes for the second stage randomization will be maintained by Investigational Drug Services Pharmacy. |
| 1073 | Fritz JM et al., 2020(19) | Yes | 1200 | Yes | NA | Randomization will occur using the REDCap randomization module. A study statistician created separate randomization allocation tables for the initial randomization and for re-randomization of non-responders. Blocked randomization with block sizes of four or six will be used. Randomization is stratified by recruitment site, gender, and active-duty status. |
| 1111 | Fu SS et al., 2017(20) | Yes | 1000 | Yes | Data collectors are blinded | Both randomizations used stratified, block randomization schemes. |
| 1019 | Germeroth LJ et al., 2019(21) | Yes | 300 | Yes | NA | At both randomization points, separate randomization schedules are used for women with a pre-pregnancy BMI of 25–29.9 kg/m2 (overweight) and for women with a BMI ≥ 30 kg/m2 (obese). Randomization schedules were generated by a statistician at the start of the study. Women will be randomized with equal probability to one of the two initial interventions stratified by their pre-pregnancy weight status. Since this is a non-restrictive SMART, women are then randomized with equal probability to one of the two postpartum lifestyle interventions, regardless of whether they have met their GWG goals. Procedurally, we created a randomization list of the four treatment sequences, stratified by initial weight status |
| 1176 | Hassett AL et al., 2023(22) | Yes | 400 | Yes | NA | First randomization uses block randomization so that those eligible for all first-stage interventions have a 1:1:1:2 chance of receiving Duloxetine; those who are eligible to receive three of the four treatments have an equal chance of receiving one of the remaining 3 treatments.  Second randomization uses block randomization so that those eligible for Duloxetine (and did not receive in the first stage) are assigned with 1:1:4 weights to Duloxetine; otherwise equally assigned among treatments not received |
| 1159 | Hibbard JC et al., 2018(23) | Yes | 180 | Yes | Investigator taking measurements is blinded | Permuted block randomization with equal probability between all allowed treatment sequences, no stratification |
| 1185 | Inwani I et al., 2017(24) | Yes | 1200 | Yes | NA | Permuted block randomization |
| 1145 | Jain S et al., 2023(25) | Yes | 180 | Yes | NA | NA |
| 1104 | Johnson JE et al., 2018(26) | Yes | 90 | NA | No | The first randomization randomizes clinical in a 3.8:1 ratio of LICF versus EIAU. Re-randomized clinics are randomized 1:1 to LICF versus HICF. Randomization includes balancing trial arms by time (3, 6, 9, 12, or 15 months) and by whether or not the clinic is a FQHC. |
| 1103 | Kilbourne AM et al., 2014(27) | Yes | 80 clinics (N = 1600 patients) | Yes | Outcome evaluators are blinded | First randomization: 1:1 stratified, permuted-block randomization  Second randomization: 1:1, stratified |
| 1170 | Kopelowicz A et al., 2023(28)­ | Yes | 330 | NA | Research assistants collecting outcomes and evaluators are blinded | NA |
| 1084 | Kor PP et al., 2023(29) | Yes | 272 | Yes | The researchers/research assistants who perform the assessment and analysis will be  blinded to the group allocations. | Permuted block randomization will be employed in this study following the allocation concealment mechanism. At all stages, an independent statistician will randomize a list of eligible caregivers via computer-generated random numbers and the caregivers will be informed of their group allocation via a sealed opaque envelope containing information about their group allocation |
| 1179 | Levy R et al., 2019(30) | Yes | 2710 | Yes | Clinical evaluators who conduct follow-up assessments will be blinded to treatment assignment. Participants, IPT/fluoxetine providers and study coordinators will not be blinded | Both randomizations: stratified, block randomization |
| 1071 | Li X et al., 2021(31) | Yes | 720 | Yes | The study was single-blinded, with the evaluators blinded to the treatment assignment | Phase 1 of the trial is a randomized controlled trial (RCT). Phase 2 of the trial uses equipoise-stratified randomization designed to allow patients or their psychiatrists to exclude inappropriate treatment based on previous experience or anticipated risk |
| 1070 | Lion KC et al., 2023(32) | Yes | 55 primary care providers, 648 patients | Yes | Two coders trained to abstract provider-documented diagnosis are blinded to study assignment | Providers will be randomized 1:1, stratified by baseline interpreter use and clinic. Randomization will occur within REDCap, using a sequence generated by the study biostatistician and implemented by a research coordinator. After 9 months, providers in the bottom two tertiles will be re-randomized 1:1 in stage 2. |
| 1083 | Liu H et al., 2021(33) | Yes | 312 | No | Patients, LHSs, nurses who perform recruitment and baseline survey, the statistician who performs randomization, and investigators who perform follow-ups will be blinded to the assignment. | After recruitment and the baseline survey, we will assign participants into group 1 and group 2 by block randomization in R program. At the check point in the SMART trial, we will reassign participants based on their suicide risk by simple randomization in the R program. The allocation ratio in randomization will be 1:1. The randomization will be performed by a statistician in the research team |
| 1131 | Markland AD et al., 2023(34) | Yes | 286 | Yes | Site investigators will be blinded to all outcomes | First stage randomization is stratified by UI symptom severity |
| 1106 | Micheletti RG et al., 2020(35) | Yes | 90 | Yes | Blinded investigators grade the standardized photographs | Both randomizations have equal allocation ratios and both will be stratified |
| 1038 | Nelson B et al., 2018(36) | Yes | 500 | Yes | Research Assistants (RAs) conducting the clinical assessments will be blind to treatment allocation (Step 2: single-blind). Step 3 of the study incorporates a randomized, double-blind, placebo-controlled stage of the study | NA |
| 1199 | O'Keefe VM et al., 2019(37) | Yes | 304 | No | Independent Evaluators (IEs) who administer the assessments and deliver Optimized Case Management are blinded. Interventionists who deliver the brief interventions, New Hope and Elders’ Resilience, are not blinded. | Both randomizations use stratified block randomization with a 1:1 allocation ratio. |
| 1034 | Osilla KC et al., 2023(38) | Yes | 530 | Yes | NA | Permuted block randomization |
| 1018 | Peter SC et al., 2023(39) | Yes | 280 | Yes | Buprenorphine providers and outcomes assessor will be blinded to condition | Randomization for Stage 1 and Stage 2 is a 2-arm, parallel, random assignment with 1:1 allocation ratio using block-randomization (blocks of 4) developed by the study statistician with separate blocks for males and females. Randomization sequence will be stored in a database where the sequence is concealed until participants are enrolled and interventions are assigned by the study coordinators |
| 1079 | Peterson BS et al., 2021(40) | Yes | 404 | Yes | Because PCORI guidelines preclude paying for any component of clinical care, insurance will need to pay for study treatments,  which in turn will preclude blinding patients and clinicians to treatment assignment. Nevertheless, all study assessments have been selected as parent- and youth-reports that require minimal to no interactions with research staff, thereby minimizing or eliminating rater bias from study staff. It is in this sense that we designate this study “single blind”. | In each randomization within trial Stages 1 & 2, eligible and consenting/assenting participants will be randomized in a 1:1 allocation to the 2 treatment regimens. Randomization will be stratified by study site, age group, and baseline symptom severity.  Randomization will be further blocked, with a relatively small block size to ensure balanced randomization over the short term; block size will not be revealed to investigators or trial staff. The study statistician will develop and monitor fidelity to the randomization sequence. The REDCap (Research Electronic Data Capture) randomization module will be used to randomize patients to study treatments; the study statistician will develop the stratified blocked randomization sequences. The randomization sequence will not be viewable. Randomization capability will be limited to the lead research coordinator and study statistician |
| 1045 | Pfammatter AF et al., 2019(41) | Yes | 400 | No | Staff are blinded | Eligible participants are randomized with equal probability to one of two first-line treatments. Randomization will be stratified by gender and baseline BMI. Re-randomization of early non-responders will be stratified by whether the non-responder did or did not lose any weight by the time of re-randomization, relative to baseline |
| 1251 | Quanbeck A et al., 2020(42) | Yes | 256 | No | NA | Both randomizations use stratified block randomization and a 1:1 allocation ratio |
| 1088 | Rabin BA et al., 2023(43) | Yes | 300 | Yes | NA | At enrollment, participants will be stratified by current COVID-19 vaccination status and status of other preventive services vis-à-vis a risk score and then randomized into the first-stage intervention. First stage non-responders will be equally re-randomized in stage 2 |
| 1030 | Sabri B et al., 2021(44) | Yes | 1266 | Yes | Study team members are blinded to first stage randomization but not the second stage | For first stage randomization, women who consent, enroll in the study, and complete the baseline survey are randomized using computer-generated block randomization stratified by length of time in the US. Randomization for the second sequence of the intervention is also computer generated. |
| 1166 | Schlechter CR et al., 2023(45) | No | 200 | NA | NA | All randomization sequences will be based on random permuted blocks with random block size and equal allocation ratios. |
| 1140 | Skolasky RL et al., 2020(46) | Yes | 945 | Yes | Study personnel who are responsible for baseline and follow-up assessments are blinded | For all randomizations, block randomization with random block sizes and a 1:1 allocation ratio will be used. |
| 1063 | Smith SK et al., 2021(47) | Yes | 400 | Yes | NA | A computer-generated random number assignment procedure in REDCap determines approach order. Randomization is stratified by site with equal allocation to each initial intervention |
| 1027 | Sripada RK et al., 2023(48) | Yes | 430 | Yes | Evaluators are blinded | Participants are randomized using a secure centralized interactive web-based randomization application. Randomization is stratified by site, gender, and initial PTSD severity, and is computer-generated using a minimization allocation method. |
| 1146 | Tamura K et al., 2020(49) | Yes | 180 | No | The research team and participants will not be blinded to the intervention or key outcomes | Prior to enrolment, research staff will assign each participant study number to one of the two starting interventions using computer-generated random numbers. The community research coordinator will enroll participants and assign a study number sequentially in order of enrolment. |
| 1076 | van Heerden A et al., 2023(50) | Yes | 900 | Yes | The allocation sequence will be generated by the unblinded biostatistician. Because of the difficulty masking study team and study participants to group allocation, the study is unblinded. However, staff assessing the primary outcome will be masked to the allocation of participants, as will study investigators. | Participants will be initially randomized at a ratio of 1:1 using varying size block randomization. At 6 months into the study, re-randomization of eligible participants will take place at a ratio of 2:1:1 again using varying size block randomization. Due to the intermittent availability of Internet access, envelopes will be used to randomize participants. The allocation sequence will be generated by the unblinded biostatistician. |
| 1286 | Velloza J et al., 2022(51) | Yes | 500 | Yes | Principal investigators are blinded | Both randomizations used variable block randomization in a 1:1 allocation ratio. |
| 1281 | Walton MA et al., 2023(52) | Yes | 700 | Yes | NA | First stage randomization employed block randomization. Second stage randomization was stratified. |
| 1057 | Wan Y et al., 2023(53) | Yes | 358 dyads | Yes | The community family health teams will not be blinded to the participants’ assignment (although investigators conducting the baseline survey and checkpoint evaluations, as well as the statistician performing the randomization, will be blinded, the lack of blinding among health teams could introduce bias). | Patients and their caregivers will be randomly assigned to either intervention group with the 1:1 allocation ratio in randomization. The randomization will be done by statisticians from the WCSPH in Sichuan University, who will not be involved in the study. The SAS 9.4 software will be used to generate random sequences and record all random numbers. In this study, we will employ central randomization. |
| 1167 | Wilroy J et al., 2023(54) | Yes | 40 | Yes | Outcome assessor is blinded | Permuted blocks in a 1:1 allocation ratio |
| 1141 | Windsor L et al., 2022(55) | Yes | 582 | NA | Data analysts are blinded | First stage: equal allocation, randomized permuted blocks with fixed block size  Second stage: stratified by first stage intervention, equal allocation  Third stage: random permuted block with fixed block size |
| 1123 | Yan X et al., 2022(56) | Yes | 150 | Yes | Coaches and participants cannot be masked | All randomizations: 1:1 allocation ratio, block randomization with fixed block size |
| 1278 | Zhao SZ et al., 2022(57) | Yes | 1200 | Yes | Smoking cessation counselors, outcomes assessors, and statistical analysts were blinded to the group allocation | First stage randomization used random permuted blocks and a 1:1 allocation ratio. In the PIM group, non-responders were randomized to OCI or continue PIM using block randomization in a 3:1 ratio. In the RIM group, non-responders were randomized to receive PIM or continue RIM in a 1:3 ratio. |
| 1279 | Zhou G et al., 2020(58) | Yes | 36 clusters, 500 individuals per cluster | No | Field staff conducting active case detections and collecting samples were masked to cluster assignments. Neither investigators nor participants were masked to the treatment allocation of indoor residual spraying. | For the first-stage randomization, clusters were stratified by risk and clusters within strata were allocated to first-stage intervention via block randomization in a 1:1 ratio. Second-stage interventions were allocated in a 1:1 allocation ratio. |
| 1155 | Zullig LL et al., 2021(59) | Yes | 800 | NA | NA | Stratified randomization based on category of PCP |
| **Primary analysis papers** | | | | | | |
| 1099 | Butzer JF et al., 2023(60) | No | NA | NA | Certified Therapeutic Recreation Specialists (CTRS) were not blinded to the exercise groups. | NA |
| 1282 | Czyz EK et al., 2021(61) | No | NA | NA | Interviewers were masked to treatment assignment | Both randomizations were stratified and treatments allocated 1:1 |
| 1271 | Fatori D et al., 2018(62) | No | NA | NA | Blinded evaluators assessed the study outcomes | Both randomizations used a sequential allocation method developed to minimize the possibility of differences between groups by balancing allocation via a computer algorithm using the Aitchison’s compositional distance to calculate the smallest difference between treatment groups, based on prognostic factors |
| 1081 | Gao K et al., 2020(63) | No | NA | NA | NA | NA |
| 1273 | Geng EH et al., 2023(64) | Yes | 1900 | Yes | Treatment assignments were only revealed to participants and study staff when randomization/re-randomization criteria met | Treatment assignments were randomized 1:1 |
| 1124 | Kruse GR et al., 2023(65) | Yes | 35 patients | Yes | Research coordinator was not blinded | One-to-one randomization |
| 1269 | Lambert SD et al., 2022(66) | Yes | 55 | Yes | Participants were not blinded to group allocation but were blinded to study objectives | Both randomizations used stratified blocks and allocated treatments 1:1. |
| 1203 | McKay JR et al., 2015(67) | No | 500 | NA | NA | Both randomizations used within-site block randomization |
| 1200 | Morgenstern J et al., 2021(68) | No | NA | NA | NA | Both randomizations used urn randomization |
| 1078 | Naar-King S et al., 2016(69) | Yes | 180 | No | Data collectors were kept blind to participants’ group assignments. | Randomization was stratified based on the presence of  adolescent comorbidities and adolescent percent overweight. |
| 1020 | Pelham Jr WE et al., 2016(70) | Yes | 150 | No | NA | NA |
| 1219 | Pistorello J et al., 2017(71) | Yes | NA | NA | NA | First-stage randomization used an adaptive-biased coin design to ensure treatments were balanced with respect to gender. Second-stage randomization was balanced for stage 1 assignment and gender. |
| 1062 | Sauer-Zavala S et al., 2022(72) | No | NA | NA | Patients and study therapists were masked to condition until the second-stage randomization. | NA |
| 1005 | Schlam TR et al., 2024(73) | Yes | 600 | Yes | NA | NA |
| 1075 | Sikorskii A et al., 2023(74) | Yes | 430 | Yes | Trained bilingual and bicultural interviewers were blinded to group assignment. | High need survivors were randomly assigned initially to either: 1) SMSH alone or 2) SMSH+TIPC (Fig. 1) in a 3:1 ratio using a minimization procedure that balanced trial arms with respect to recruitment location, site of cancer (breast, colon, lung, other), and last treatment received (chemotherapy/targeted therapy or radiation). |
| 1002 | Stanger C et al., 2020(75) | No | NA | NA | Research assistants were not blinded to condition (due to staffing constraints). | NA |
| **Protocol + primary analysis papers** | | | | | | |
| Primary: 1232  Protocol: 1032 | Primary: Fortney JC et al., 2021(76)  Protocol: Fortney JC et al., 2020(77) | Yes | 1000 | Yes | Treatment group assignment was masked to telephone interviewers who administered surveys | Both randomizations used blocking and stratification |
| Primary: 1014  Protocol: 1054 | Primary: Gonze BB et al., 2020(78)  Protocol: Morais Pereira Simões et al., 2019(79) | Yes | 24-50 | No | All researchers who did the assessments were blinded. The researchers who did the group allocation, sent messages, and participated in gamification were not blinded. | Randomized participants in blocks of six (two to each group). Randomization was concealed using opaque envelopes and the sequence of group allocation was computer generated. |
| Primary: 1223  Protocol: 1122 | Primary: Gunlicks-Stoessel M et al., 2019(80)  Protocol: Gunlicks-Stoessel M et al., 2016(81) | No | NA | NA | Evaluators conducting outcome assessments and interviews were blinded. | Both randomizations allocated treatments 1:1 |
| Primary: 1008  Protocol: 1297 | Primary: Igudesman D et al., 2023(82)  Protocol: Corbin KD et al., 2022(83) | Yes | 72 | No | Patients wore a blinded continuous glucose monitor at baseline and each subsequent measurement visit. | Study statisticians generated permuted block randomization stratified by site and confidentially. |
| Primary: 1177  Protocol 1239: | Primary: Karp JF et al., 2019(84)  Protocol: Karp JF et al., 2016(85) | No | 135 | NA | Blinding was maintained by the data manager, and independent evaluators and the biostatisticians were masked to random assignment until outcomes analysis was complete. | The first randomization employed a permuted block scheme to allocate treatments in a 2:1:2 ratio; the second randomization allocated treatments with equal probability. |
| Primary: 1216  Protocol: 1299 | Primary: Morin CM et al., 2020(86)  Protocol: Morin CM et al., 2016(87) | Yes | 224 | Yes (in the supplement) | Single-blinded (blinded raters) | First-stage randomization was stratified and interventions were allocated 1:1. Second-stage randomization was stratified. |
| Primary: 1298   Protocol: 1193 | Primary: Mustanski B et al., 2023(88)  Protocol: Mustanski B et al., 2020(89) | Yes | 1878 | No | NA | Initial randomization to 1 of 4 embedded regimes used stratified block randomization. They randomized within 8 strata comprising all combinations of language preference (English or Spanish), rurality (living in an urban or rural zip code), and lifetime anal sex experience (any or none). Within each stratum, embedded regimes were assigned using a permuted block design with blocks of size 4. |
| Primary: 1154  Protocol: 1080 | Primary: Patrick ME et al., 2021(90)  Protocol: Patrick ME et al., 2020(91) | Yes | 675 | Yes | NA | Used computer-generated, blocked randomization schemes for eah of the two potential randomizations. First randomization allocated treatments in a 1:1:1 ratio; second randomization in a 1:1 ratio |
| Primary: 1228  Protocol: 1077 | Primary: Schmitz JM et al., 2024(92)  Protocol: Schmitz JM et al., 2018(93) | Yes | 160 | Yes | Placebo-controlled pharmacotherapy augmentation | First-stage randomization used urn randomization |
| Primary: 1121  Protocol: 1125 | Primary: Schoenfelder EN et al., 2019(94)   Protocol: Chronis-Tuscano A et al., 2016(95) | No | NA | NA | Masked assessments | Randomized 23 dyads to initial MSM and 23 dyads to initial BPT. All participants were randomized twice. |
| Primary: 1246  Protocol 1245: | Primary: Sherwood NE et al., 2022(102)  Protocol: Sherwood NE et al., 2016(103) | Yes (post-hoc) | NA | Yes (post-hoc) | Weight loss coaches and participants were blinded to the first randomization but not to the results of the second randomization. Study team members were blinded. | Both randomizations used stratified block randomization. |
| Primary: 1112  Protocol: 1274 | Primary: Smith SN et al., 2022(96)  Protocol: Kilbourne AM et al., 2018(97) | Yes | 94 high schools (169 school professionals) | Yes | NA | Both randomizations allocated treatments with equal probability. These randomizations were stratified. |
| Primary: 1250  Protocol: 1137 | Primary: Somers TJ et al., 2023(98)  Protocol: Kelleher SA et al., 2017(99) | Yes | 284 | No | Participants were not blinded to condition. Study therapists were blinded to participant’s assessment responses and analyses were conducted by team members who did not interact with participants. | First stage randomization allocated treatments 1:1. |
| Primary: 1097  Protocol: 1012 | Primary: Wyatt G et al., 2021(100)  Protocol: Sikorskii et al., 2017(101) | Yes | 331 | Yes | Interviewers were blinded to dyad's group assignments. | Randomization was performed using a computerized minimization procedure from the central study office to ensure allocation concealment and blinding of data collectors. By design, the odds of allocation to reflexology or meditative practices were the same, but the odds of allocation to the control group was three times smaller. The balancing factors were recruitment location, site of cancer (breast, lung, colon, prostate, other), stage of cancer (early, late), and treatment type (hormonal therapy alone or chemotherapy and/or targeted therapy). Dyads with nonresponding patients were rerandomized in 1:1 ratio. The technique for the second randomization was the same as for the first, with the same balancing factors. |

**TABLE S5. Estimation and inference for embedded regimes or deeply tailored regimes in primary analysis papers**

| **SR_number** | **First author** | **Were any embedded regimes analyzed?** | **What was the objective of the analysis of the embedded regimes?** | **What methods were used to analyze the embedded regimes?** | **Were any deeply tailored regimes analyzed?** | **What was the objective of the analysis of the deeply tailored regimes?** | **What methods were used to analyze the deeply tailored regimes?** |
| --- | --- | --- | --- | --- | --- | --- | --- |
| 1099 | Butzer JF et al., 2023(60) | No | NA | NA | No | NA | NA |
| 1282 | Czyz EK et al., 2021(61) | No | NA | NA | No | NA | NA |
| 1271 | Fatori D et al., 2018(62) | Yes | Compare outcomes (OCD symptom reduction measured by Y-BOCS scores) among four adaptive treatment strategies defined by the initial treatment and treatment response over time | Cubic polynomial modeling and Mann-Whitney and Wilcoxon tests | No | NA | NA |
| 1232 | Fortney JC et al., 2021(76) | No | NA | NA | No | NA | NA |
| 1081 | Gao K et al., 2020(63) | No | NA | NA | No | NA | NA |
| 1273 | Geng EH et al., 2023(64) | Yes | Provide effectiveness estimates for a range of possible sequenced strategies and contrast the standard-of-care strategy (SOC, with SOC-Outreach if a lapse occurred) with each ‘fully active’ adpative strategy | Longitudinal TMLE | No | NA | NA |
| 1014 | Gonze BB et al., 2020(78) | No | NA | NA | No | NA | NA |
| 1223 | Gunlicks-Stoessel M et al., 2019(80) | Yes | Examine the clinical and psychosocial outcomes of each of the four IPT-A algorithms | Weighting and replication (Nahum-Shani et al., 2012) | No | NA | NA |
| 1008 | Igudesman D et al., 2023(82) | No | NA | NA | No | NA | NA |
| 1177 | Karp JF et al., 2019(84) | Yes | To compare CBT-CBT, CBT-PT, PT-PT, PT-CBT, and EUC based on Stage 2 response | G-computation (Bembom and van der laan), linear mixed effects model (Miyahara and Wahed) | No | NA | NA |
| 1124 | Kruse GR et al., 2023(65) | No | NA | NA | No | NA | NA |
| 1269 | Lambert SD et al., 2022(66) | No | NA | NA | No | NA | NA |
| 1203 | McKay JR et al., 2015(67) | No | NA | NA | No | NA | NA |
| 1200 | Morgenstern J et al., 2021(68) | Yes | To test the adaptive treatment strategies (i.e., combinations of treatment) on week 13 outcomes | Weighting and replication (Nahum-Shani et al., 2012) | No | NA | NA |
| 1216 | Morin CM et al., 2020(86) | Yes | Evaluate the comparative efficacy of 4 treatment sequences involving psychological and medication therapies for insomnia and examine the moderating effect of psychiatric disorders on insomnia outcomes | Weighting and replication (Nahum-Shani et al., 2012) | No | NA | NA |
| 1298 | Mustanski B et al., 2023(88) | No | NA | NA | No | NA | NA |
| 1078 | Naar-King S et al., 2016(69) | No | NA | NA | No | NA | NA |
| 1154 | Patrick ME et al., 2021(90) | Yes | To compare outcomes of the APIs (i.e., intervention sequences) embedded in the study design for the intervention group | Weighted and replicated GEE (Lu et al 2016, Nahum-Shani) | No | NA | NA |
| 1020 | Pelham Jr WE et al., 2016(70) | Yes | What is the most effective treatment protocol, or pattern of initial treatment and conditional secondary/adaptive treatment among the four that we employed (BM, Behavioral-Behavioral [BB], Medication-Behavioral [MB], and Medication-Medication [MM])? | Weighting and replication (Nahum-Shani et al., 2012) | No | NA | NA |
| 1219 | Pistorello J et al., 2017(71) | No | NA | NA | No | NA | NA |
| 1062 | Sauer-Zavala S et al., 2022(72) | No | NA | NA | No | NA | NA |
| 1005 | Schlam TR et al., 2024(73) | No | NA | NA | No | NA | NA |
| 1228 | Schmitz JM et al., 2024(92) | No | NA | NA | No | NA | NA |
| 1121 | Schoenfelder EN et al., 2019(94) | No | NA | NA | No | NA | NA |
| 1246 | Sherwood NE et al., 2022(102) | Yes | To compare 6m and 18m weight loss across the 4 AIs, overall (Early PCM, Early ABT, Late PCM, Late ABT) and by participant subgroups (sex, BMI category, binge eating disorder status) | Estimated GEE predicted weight change (Nahum Shani 2019) | No | NA | NA |
| 1075 | Sikorskii A et al., 2023(74) | Yes | To test the relative effectiveness of the three dynamic treatment regimes (DTRs) embedded within the SMART | Weighting and replication (Nahum-Shani et al., 2012) | No | NA | NA |
| 1112 | Smith SN et al., 2022(96) | Yes | To evaluate the effectiveness of an adaptive implementation strategy versus providing REP alone | Marginal, weighted least squares (NeCamp et al 2017) | No | NA | NA |
| 1250 | Somers TJ et al., 2023(98) | Yes | Compare the eight intervention sequences embedded in the SMART study design | Modification of Weighting and replication (Nahum-Shani et al., 2012) | No | NA | NA |
| 1002 | Stanger C et al., 2020(75) | Yes | Exploratory analyses compared the four treatment strategies embedded in the trial" to evaluate the effects of different adaptive treatment strategies on outcomes (weeks of continuous abstinence) | Weighting and replication (Nahum-Shani et al., 2012) | No | NA | NA |
| 1097 | Wyatt G et al., 2021(100) | Yes | To compare four decision rules (DRs) with respect to the primary outcome of severity of fatigue, and secondary outcomes of summed index of severity of other symptoms, depression, and anxiety | Weighting and replication (Nahum-Shani et al., 2012) | No | NA | NA |
